# Genetic regulation of cell type–specific chromatin accessibility shapes immune function and disease risk

**DOI:** 10.1101/2025.08.27.25334533

**Authors:** Angli Xue, Jianan Fan, Oscar A. Dong, Hao Lawrence Huang, Ling Chen, Peter C. Allen, Eleanor Spenceley, Eszter Sagi-Zsigmond, Blake Bowen, Albert Henry, Anna S.E. Cuomo, Hope A. Tanudisastro, Zhen Qiao, Elizabeth Dorans, Eyal Ben-David, Kyle Kai-How Farh, Linfeng Hu, Yijia Christiana Liu, Drew Neavin, Arthur S. Lee, Anne Senabouth, Caitlin Bartie, Rachael A. McCloy, Venessa Chin, Wei Zhou, Alkes L. Price, Katrina M. de Lange, Gemma A. Figtree, Alex W. Hewitt, Daniel G. MacArthur, Joseph E. Powell

**Author notes:** Corresponding authors: Joseph E. Powell, Angli Xue. J.F., O.A.D., H.L.H, L.C., P.C.A., E.S. contributed equally to this work and should be regarded as joint second authors.

## Abstract

Understanding how genetic variation influences gene regulation at the single-cell level is crucial for elucidating the mechanisms underlying complex diseases. However, limited large-scale single-cell multi-omics data have constrained our understanding of the regulatory pathways linking variants to cell type-specific gene expression. Here we present chromatin accessibility profiles from 3.5 million peripheral blood mononuclear cells (PBMCs) across 1,042 donors, generated using single-cell ATAC-seq, including ∼100,000 PBMCs from 90 donors using multiome (RNA+ATAC) sequencing, with matched whole-genome sequencing. We identified 440,996 chromatin peaks across 28 immune cell types and mapped 243,225 chromatin accessibility quantitative trait loci (caQTLs), of which 60% were cell type-specific. Rare variant analysis revealed associations with 27,927 peaks, recapitulating the same cell type-specific regulatory architecture. Colocalization with the eQTLs from scRNA-seq data (5.4 million PBMCs) identified 31,688 candidate *cis*-regulatory elements; around half (44.40%) show evidence of putative mediating effects via chromatin accessibility. Integrating caQTLs with GWAS summary statistics for 16 diseases and 44 blood traits uncovered 4.5% - 22.6% more colocalized signals compared with using eQTLs alone, many of which have not been reported in prior studies. We show that the limited concordance between GWAS and eQTL signals can be substantially improved by incorporating cell type– and tissue-specific context, distal regulatory effects, models allowing for multiple causal variants, and promoter/enhancer priming, together increasing overlap from ∼20% to ∼60%. In addition, using a graph neural network, we inferred peak-to-gene relationships from unpaired multiome data by integrating caQTL and eQTL signals, achieving up to 80% higher accuracy than with paired multiome data lacking QTL information. This gain translated into improved gene regulatory network inference, enabling the identification of 128 additional transcription factors (TF)–target gene pairs (a 22% increase). Together, these results provide a comprehensive single-cell map of chromatin accessibility and genetic variation in human circulating immune cells, establishing a powerful resource for dissecting cell type-specific regulation and advancing our understanding of genetic risk for complex diseases.

## Introduction

Understanding how genetic variation influences complex traits and disease risk remains a major challenge in human genomics. Although genome-wide association studies (GWAS) have identified tens of thousands of risk loci, most lie in non-coding regions^1^, complicating interpretation. One approach to bridge this gap is to map molecular quantitative trait loci (molQTLs), identifying genetic variants that affect intermediate molecular phenotypes such as gene expression or chromatin accessibility, and test for colocalization with GWAS loci to infer potential causal mechanisms^2^.

Most molQTL studies have focused on expression QTLs (eQTLs), derived from bulk RNA-sequencing, which have low sensitivity for cell type specific effects^3,4^. More recently, increasingly well-powered studies have utilized single-cell RNA sequencing (scRNA-seq) to identify how genetic regulation of gene expression varies across cell types and cell states^5–9^. However, many GWAS signals still lack corresponding regulatory effects, leaving their underlying molecular signatures unresolved. This gap is often referred to as the “missing regulation” problem^10–13^.

Chromatin accessibility QTLs (caQTLs) offer a complementary modality that directly reflects the physical state of regulatory elements, capturing non-coding variant effects on enhancer and promoter activity. Recent studies suggest that caQTLs colocalized more GWAS loci than eQTLs^10,11,14,15^, particularly at distal elements, but current caQTL maps remain limited in scale, resolution, and tissue diversity. Moreover, many caQTLs lack corresponding eQTLs, potentially due to less power to detect eQTL than caQTL, different test window sizes, or eQTL being more cell type-specific than caQTL^11^.

In immune cells, key mediators of many complex diseases, caQTL mapping has been restricted by small sample sizes^16^, low single-cell resolution, or reliance on genotypes imputed from the chromatin data. Moreover, dynamic, cell state-dependent regulatory effects, which may be critical for immune traits, are often overlooked. Integrating single-cell chromatin and expression profiles with genotype data has the potential to uncover these effects and refine cell type-specific gene regulatory networks (GRNs)^17,18^.

Here, as part of the TenK10K study^19^, we generated a large-scale atlas of genetic effects on single-cell chromatin accessibility in peripheral blood mononuclear cells (PBMCs). We profiled over 4.6 million single-cell ATAC-seq and 95,728 multiome (RNA+ATAC) cells across 1,042 individuals. After stringent quality control, we analysed 3.5 million high-quality nuclei from 922 donors for discovery and 60 for replication, with matched whole genome sequence (WGS) data. We identified 243,225 caQTLs across 28 immune cell types, with more than half showing cell type-specific effects. Integration with eQTL data revealed over 70,000 colocalized signals, including 25,280 candidate *cis*-regulatory elements supported by Mendelian randomisation. We detected thousands of caQTLs with cell state-dependent effects, and integrating caQTLs uncovered thousands of GWAS colocalizations missed by eQTLs alone. Finally, combining caQTL and eQTL information improved GRN inference, outperforming conventional multiome-based methods.

Together with accompanying manuscripts exploring the broader impacts of common and rare genetic variation^19^ and tandem repeat variants^20^ on gene expression, this work provides a comprehensive single-cell atlas of genetic regulation of chromatin accessibility for human circulating immune cells. By filling gaps left by eQTL-only approaches, it sheds light on how non-coding variation influences immune cell function and disease risk through chromatin-mediated pathways.

## Results

### The TenK10K multiome cohort

We generated the TenK10K phase 1 multiome dataset by profiling chromatin accessibility (scATAC-seq) in 4.5 million nuclei from 952 donors, as well as multiome (RNA+ATAC) from 90 donors (**Methods** and **Supplementary Tables 1-6**). The 952 donors were obtained from the TenK10K pilot cohort, known as OneK1K or Tasmanian Ophthalmic Biobank (TOB)^5^ (**Methods**). These data were complemented by 5.4 million PBMC scRNA-seq profiles from 1,925 donors^19^, including 932 overlapped with matched scATAC-seq profiles (**Methods; Fig. 1A**). Cell types were annotated via label transfer using a CITE-seq reference^21^ and a bridging multiome dataset (**Methods**), yielding 28 immune cell types consistent with those in the scRNA-seq dataset (**Fig. 1B–C** and **Supplementary Tables 4-6**). After quality controls, 3,472,552 scATAC-seq nuclei from 922 donors were retained for discovery analyses (excluding 10 donors excluded during demultiplexing), with additional 60 donors used for replication (**Fig. 1D**). On average, each donor contributed 3,766 nuclei; CD14+ monocytes (CD14_Mono_) were the most abundant cell type, while proliferating CD8+ T cells (CD8_Proliferating_) were the rarest (**Fig. 1E**). The workflow and key metrics during the quality control was described in **Supplementary Fig. 1**.

**Figure 1.**
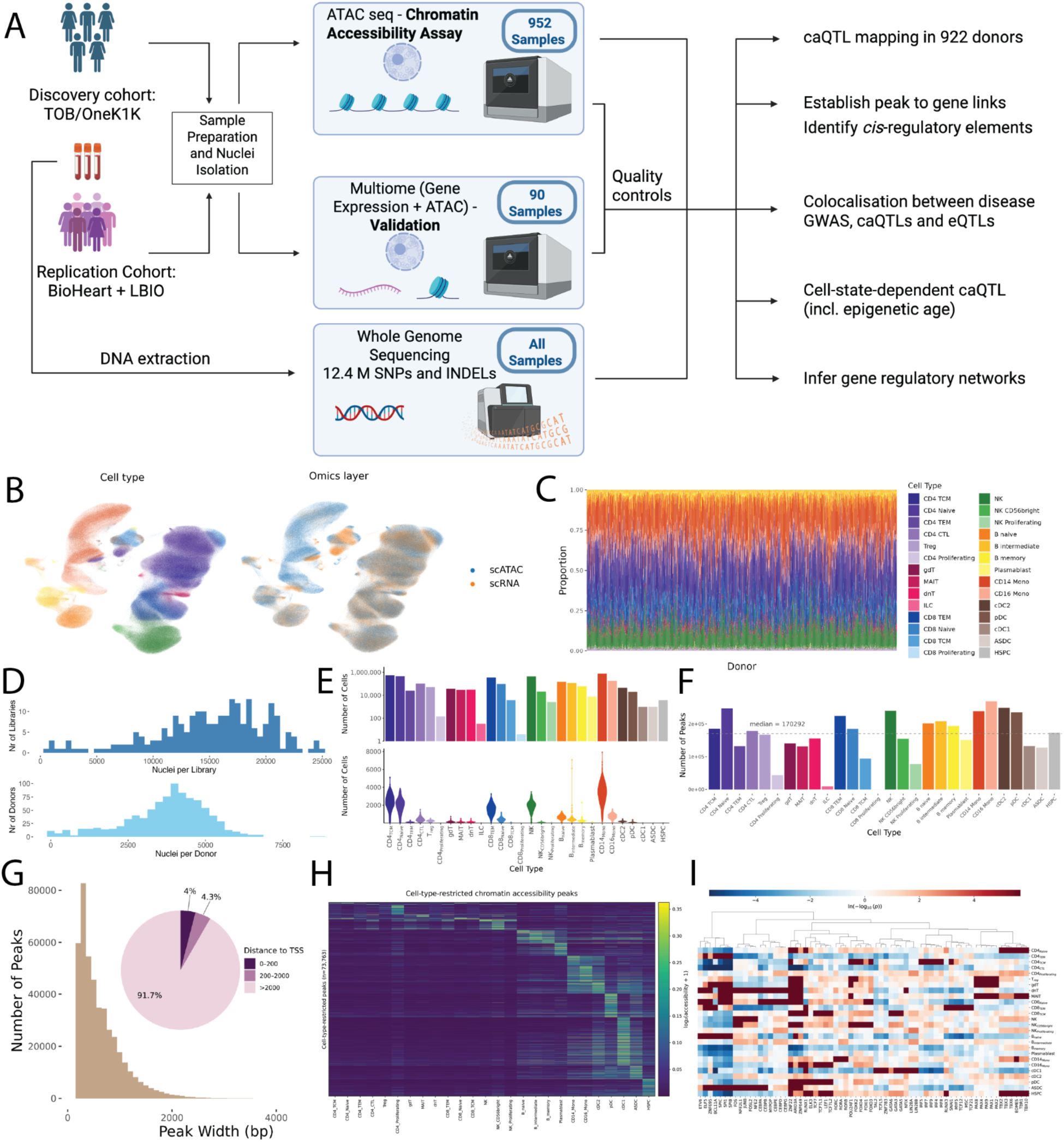
Population-scale single-cell chromatin accessibility and multi-omics profiling across 922 donors identifies 28 immune cell types and 12 million genetic variants. (**A**) Overview of the TenK10K cohort, sequencing strategy and analysis workflow (**B**) UMAP projection of scATAC-seq profiles annotated via label transfer from scRNA-seq, showing 28 immune cell types. The non-overlapping clusters are driven by differences in CD14 monocyte proportions between the scATAC and scRNA datasets. A more detailed explanation can be found in the **Methods** section. (**C**) Cell type composition across all donors. (**D**) Distribution of the number of nuclei per library and donor. (**E**) Nuclei counts per cell type after quality control. Top: total number of nuclei per cell type; Bottom: violin plots showing per donor distributions. (**F**) Number of high-quality chromatin accessibility peaks identified per cell type. (**G**) Peak width distribution across all peaks. Inset: pie chart showing the proportion of peaks by distance to the nearest transcription start site (TSS) (**H**) Heatmap of cell type-specific marker peaks across 26 immune cell types (two rare types excluded). Color scale reflects log_2_(RPKM+1) chromatin accessibility. (**I**) Transcription factor motif enrichment in marker peaks across immune 26 cell types (two rare types excluded). Color scale reflects –log_10_(*p*-value) of enrichment. The dendrogram tree of the heatmap indicates the subclusters of transcription factors and cell types based on the hierarchical clustering.

Chromatin peaks were called independently in each cell type using MACS3^22^ (via SnapATAC2^23^) and merged to generate a unified list of 440,996 peaks (**Fig. 1F**). Peak widths ranged from 201 to 5,298 bp (median 626 bp), and the majority of peaks were distal to promoter regions: only 4.0% were within 200 bp of a transcription start site (TSS), and most were > 2 kb from the nearest TSS (**Fig. 1G**). Compared with three datasets (ENCODE v4, Zhang et al.^24^ and 10X PBMC ATAC v2), only 44,015 peaks (9.98%) lacked overlap with known chromatin peaks or candidate *cis*-regulatory elements (cCREs) (**Methods** and **Supplementary Table 7**).

To assess the biological specificity of these peaks, we clustered cell type-enriched features and observed grouping by hematopoietic lineage (**Fig. 1H** and **Supplementary Figure 2**). Transcription factor (TF) motif enrichment analyses supported known lineage functions: for example, monocyte-specific peaks were enriched for *CEBP* family motifs (*CEBPB*, *CEBPD*), consistent with their established roles in monocyte differentiation and activation (**Fig. 1I**).

Together, these results confirm that chromatin accessibility profiles capture meaningful cell type-specific regulatory signals and reveal novel candidate elements across the immune landscape.

### Cell type–specific genetic regulation of chromatin accessibility

To investigate how genetic variation influences chromatin accessibility, we mapped chromatin accessibility QTLs (caQTLs) across 28 immune cell types using data from 922 donors. We tested 440,996 peaks against all common variants (MAF ≥ 5%) within 1 Mb of the peak center, encompassing ∼9 million SNPs and 3.4 million indels (**Methods**). Using permutation-based correction and a *q*-value < 0.05 significance threshold, we identified 243,225 significant caQTLs (unique variant–peak–cell type triplets), corresponding to 100,486 unique peaks (termed caPeaks) showing at least one significant association (**Fig. 2A**). caQTL discovery scaled with sample size per cell type (**Fig. 2B**), suggesting that further sampling, especially for rare cell populations, would increase detection power. Wider peaks also tend to be caPeaks than those narrower ones (**Supplementary Fig. 3**).

**Figure 2.**
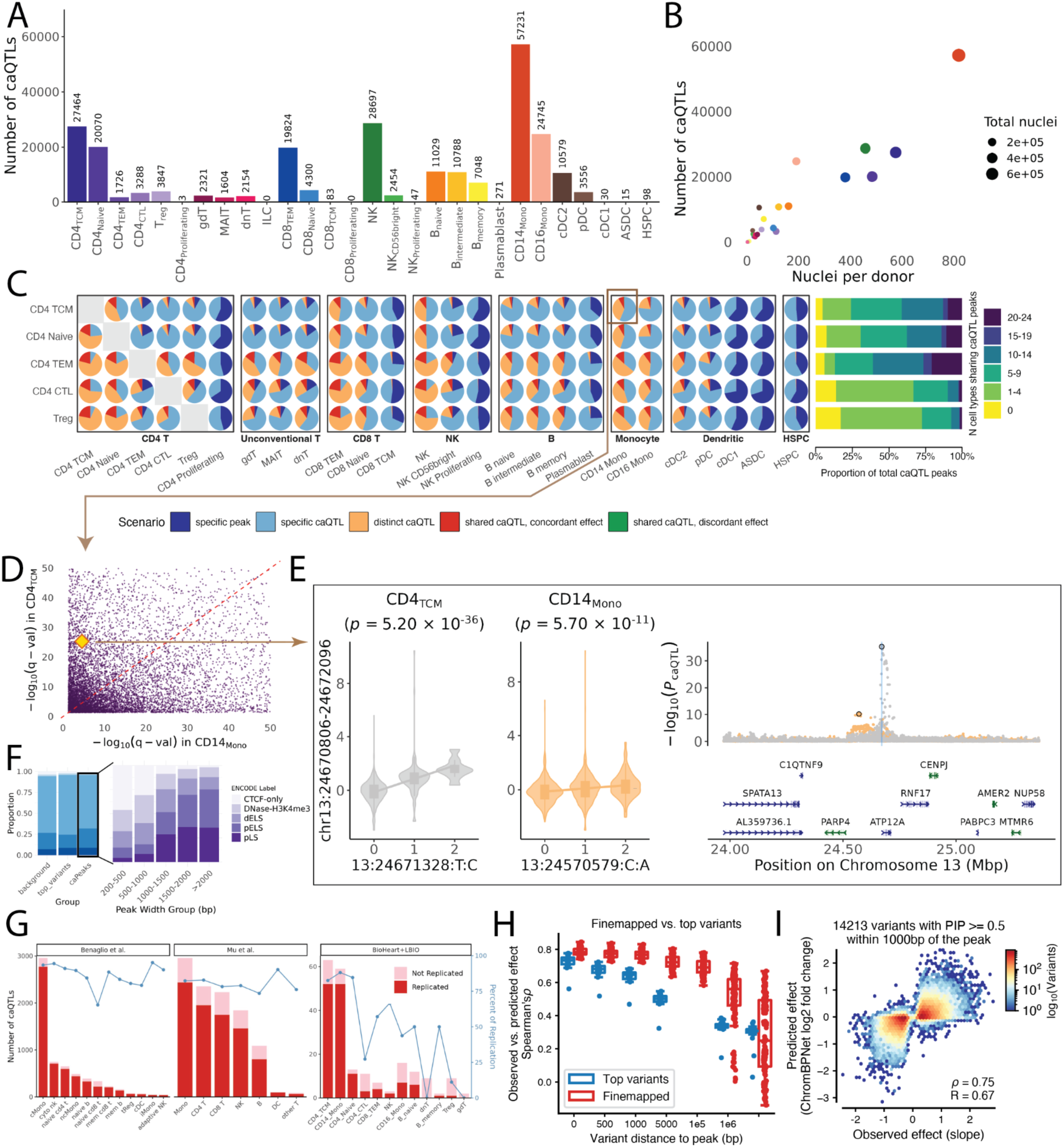
Common chromatin accessibility QTLs (caQTLs) show cell type-specific effects. (**A**) Number of significant caQTLs across 28 immune cell types. The CD8_Proliferating_ and innate lymphoid cells (ILCs) were not tested due to the limited donor size and annotated as 0. (**B**) Relationship between the number of caQTL signals and nuclei per donor across cell types (**C**) Cell type-specificity of CD4^+^ T cell caQTLs, categorized into five different scenarios (pie chart). Bar plot (right) shows the distribution of caPeaks by the number of cell types in which the effects are shared (scenarios 3-5) with. The pie chart highlighted by a brown framework represents the scenario of CD4_TCM_ caQTLs in CD14_Mono_, which will be further compared in panel D. (**D**) Comparison of caQTL significance (-log_10_(*q*-value)) between CD4_TCM_ and CD14_Mono_. The yellow star highlights the caPeak that will be demonstrated in panel E. (**E**) Left: Combined violin and boxplot of the caQTL (chr13:24670806-24672096) for its top variants in CD4_TCM_ and CD14_Mono_. The *y*-axis denotes the *z*-score normalized pseudobulk values. Right: Locus plot of the caQTL. The upper panel depicts the significant level of all the associated variants. The x-axis indicates the genomic location and the y-axis indicates -log_10_(nominal *p*-values). The top variants in CD4_TCM_ and CD14_Mono_ are highlighted with a black circle and the peak is represented as a blue vertical line. The color of the locus plot is associated with the cell type, the same as the violin plot on the left. The bottom panel shows the gene location track. (**F**) Functional annotation of regulatory elements using ENCODE v3 cCRE categories. Left: distribution across all peaks, top caQTL variants, and caPeaks. Right: annotation of caPeaks stratified by peak width. (**G**) Replication of caQTLs in two published PBMC datasets, showing significance of overlap and effect direction concordance (**H**) Spearman’s correlation between observed and ChromBPNet-predicted effect sizes, grouped by top or fine-mapped variants and stratified by variant distance to the peak in x-axis. Each dot is a cellular context. (**I**) The observed effect size (slope) versus predicted log_2_ fold-change for fine-mapped variants within tested peaks.

The top variants of caQTLs were typically proximal to the regulated peak (median distance = 7.7 kb; 88.1% within ±100 kb), but 2.7% were >500 kb away (**Supplementary Fig. 4**). Expanding the testing window from ±10 kb to ±1 Mb in CD14_Mono_ identified 4,291 additional significant peaks, with ∼85% showing >10-fold increased statistical support (**Supplementary Fig. 5**). These results underscore the importance of considering distal regulatory effects in caQTL mapping^25^.

To quantify cell type specificity, we used a five-category model comparing caQTL signals across cell type pairs (**Methods**, **Fig. 2C**, and **Supplementary Fig. 6-7**). Existing methods for assessing cell type-specificity across conditions, such as mashR^26^, are restricted to features detectable in all conditions (here, peaks called across cell types). Consequently, only a subset of peaks can be tested - for example, just 25,892 (5.87%) are accessible across all 26 cell types (**Supplementary Fig. 8**). This limitation motivated our use of this approach to evaluate cell type-specificity, which has been applied in the TenK10K flagship project for eQTLs^27^. The categories ranged from the chromatin being open only in one cell type (Scenario 1), chromatin being open in both cell types but caQTL being only detected in one cell type (Scenario 2), to distinct caQTL variants for the same peak in two cell types (Scenario 3), and shared caQTLs with concordant or discordant allelic effects (Scenarios 4–5 respectively). For example, a subset of caPeaks are both significant in CD14_Mono_ and CD4+ central memory T cells (CD4_TCM_) (**Fig. 2D**). However, some shared caPeaks had distinct (i.e., independent) top variants in two cell types, suggesting distinct regulatory mechanisms for the same peak in different cell types. For instance, chromatin peak chr13:24670806-24672096 is a caPeak in CD4_TCM_ (*q*-value = 5.03 × 10^-26^) and CD14_Mono_ (*q*-value = 2.77 × 10^-5^) and their top variants (13:24671328:T:C in CD4_TCM_ and 13:24570579:C:A in CD14_Mono_) are independent of each other (LD *r*^2^ = 4.5 x 10^-7^) (**Fig. 2E**). We performed conditional analysis of this peak in CD14_Mono_, fitting the genotype of the top variant 13:24570579:C:A in the association model, and identified a significant secondary signal, 13:24675992:C:G, which is ∼4.6kb away and in nearly perfect LD (LD *r*^2^ = 0.94, D’ = 1) with the top variant (13:24671328:T:C) in CD4_TCM_. These results revealed genetic effects on chromatin accessibility in a cell-type-specific manner through independent variants at the same locus.

Functional annotation of caPeaks using ENCODE v3 revealed that 64.2% overlap distal enhancers and 22.8% overlap proximal enhancers (**Fig. 2F**). caPeaks were enriched for promoter and enhancer elements relative to the background peak set (Chi-squared test, *p*-value < 2.2 × 10^-16^). Peak width correlated with annotation class: broader peaks were more likely to overlap promoters, while narrow peaks tended to map to CTCF binding sites. We also identified 11,585 caPeaks in gene deserts (> 500 kb without annotated genes), many of which were enriched for distal enhancers (72.1% vs. 64.2% genome-wide, *p*-value < 2.2 × 10^−16^), suggesting potential long-range regulatory roles of these peaks. To improve the functional annotation, we performed caQTL fine-mapping using SuSiE^28^. In total, we identified 242,838 credible sets in 213,264 caPeaks spanning all cell types; notably, most (88.56%) caPeaks harboured a single credible set with cumulative posterior inclusion probability (PIP) > 0.9 (**Supplementary Fig. 9**). Compared to the top variants, fine-mapped variants are on average slightly closer to the peaks (**Supplementary Fig. 4A-B**). Integrating these fine-mapped results helps explain cell type-specific signals (Scenario 3; **Fig 2E**). For example, we observed one credible set in CD4_TCM_ but two in CD14_Mono_ (**Supplementary Fig. 10**): a shared set proximal to the caPeak present in both cell types, and a second, intergenic set detected only in CD14 monocytes.

We assessed replication in three ways. First, we compared our results to two published studies on caQTL mapping in PBMCs^16,29^ (*N* = 13 and 48), observing high replication (63–95%, assessed based on whether our caQTLs were reported as significant in either of two published studies) and effect direction concordance (78–90%) despite differences in cell annotation, model, and donor health status (**Fig. 2G** and **Methods**). In total, the two published studies reported 10,884 unique caPeaks, of which 10,011 were also identified in our study, highlighting our substantially greater power to detect an additional 90,475 caPeaks in PBMCs. Second, we performed internal replication using multiome data from 60 donors with harmonized cell type annotations. For the two most abundant cell types (CD4_TCM_ and CD14_Mono_), we achieved replication rates of 82.5% and 88.1%, respectively (**Fig. 2G**). For other cell types, replication rates are lower, likely due to fewer nuclei per donor (7,384 and 5,883 in two published studies versus 738 in ours), and/or our pseudobulk mapping and linear regression approaches^30^ compared to joint modeling of population-level and allele-specific read counts in a likelihood framework^31^. To complement these analyses and more directly assess allelic effects while mitigating mapping bias, we additionally performed WASP analysis^32^ (**Methods**), which estimates allelic-specific effects on chromatin accessibility while correcting for mapping bias. This approach yielded consistent replication rates (e.g., Spearman’s *r* = 0.62 for CD14_Mono_) despite the modest sample size (n = 16 donors; **Supplementary Fig. 11**). Our results thus demonstrate strong reproducibility across cohorts, methods, and biological contexts.

Finally, we further evaluated the caQTL effect sizes using ChromBPNet^33^, a deep learning model that trains on scATAC-seq data to predict regulatory variant effects (**Methods**). Predicted and observed effect sizes were strongly correlated for fine-mapped variants (PIP > 0.5) within peak regions (Spearman’s *ρ* = 0.75), with accuracy decreasing as a function of distance (**Fig. 2H–I** and **Supplementary Fig. 12-13**). Notably, prediction performance remained high for fine-mapped variants located 1-5kb from peaks (Spearman’s *r*∼0.7), compared to top variants (*r*∼0.4), suggesting that distal top variants are more likely to represent tagging SNPs lacking direct regulatory function, whereas fine-mapped variants at similar distances remain enriched for functional effects. In contrast, novel peaks exhibited consistently lower ChromBPNet prediction accuracy and greater donor-level sparsity relative to known peaks (**Supplementary Fig. 14–15**). These findings highlight the ability of chromatin accessibility data to predict the regulatory impact of genetic variants, particularly those within the functional regions.

### Mapping the effects of rare variation on single-cell chromatin accessibility

While several studies have identified common variants associated with chromatin accessibility^16,29^, the contribution of rare variants remains largely underexplored. Leveraging the large sample size of this cohort, we performed association testing for rare variants (MAF < 5%) within a *cis*-window (±100kb) around the peak center using SAIGE-QTL^34^, and applied a Cauchy combination of burden tests and SKAT to define significant signals^35^ (**Methods** and **Supplementary Figure 16**). In total, we identified 216,159 rare caQTLs across 26 cell types (**Fig. 3A**). To assess whether these rare variant signals were driven by linkage with common variants near the 5% MAF threshold, we performed an iterative conditional analysis incorporating the genotypes of significant common variants into the rare caQTL model. After conditioning analysis, we identified 62,866 rare caQTLs corresponding to 38,161 unique peaks (**Fig. 3A**), of which 24.1% (15,154/62,866) overlap with common variant caPeaks in the same cell type. Notably, only 24.6% (53,195 / 216,159) of rare caQTLs remained significant after conditioning on common caQTLs (**Fig. 3A** and **Supplementary Table 8**).

**Figure 3.**
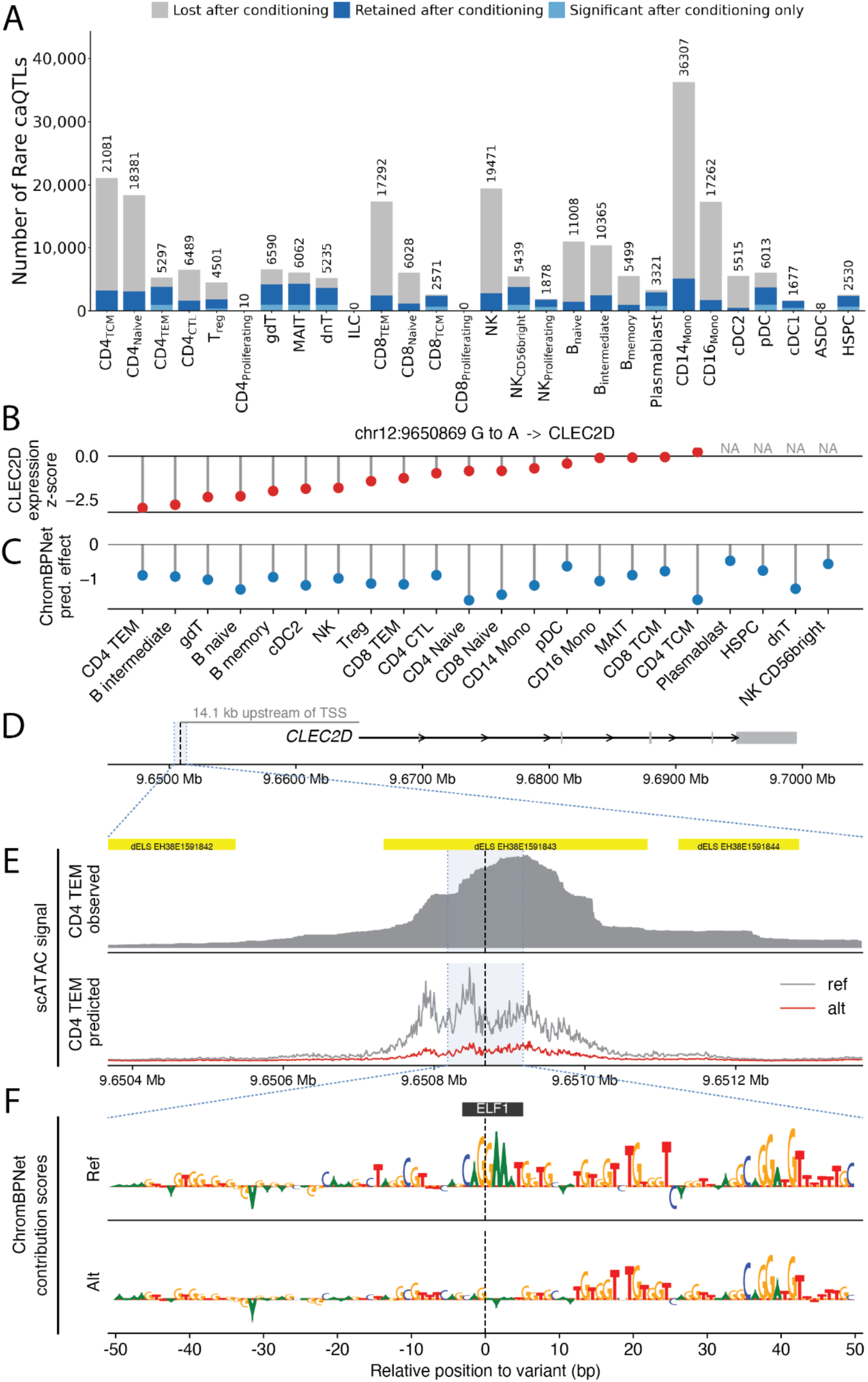
Rare caQTLs are enriched in regulatory elements and affect gene expression by disrupting transcription factor motifs. (**A**) Number of significant rare variant caQTLs across 28 immune cell types. The number above the stacked bar represents the number of rare caQTLs before adjusting the nearby top common variant. Grey bar indicates the caQTL that are not significant after conditioning. The dark blue bar indicates those remain significant after conditioning. The light blue bar indicates those only significant after conditioning. (**B**) Red lollipop plot showing target *CLEC2D* gene expression *z*-scores, ordered from the most to least affected. The *z*-scores represent the difference in expression between carriers of the alternative allele versus the reference allele. (**C**) Blue lollipops indicate the corresponding difference in ChromBPNet-predicted chromatin accessibility of peak chr12:9650649–9651305. (**D**) Genomic context of the variant relative to the target gene. The black dashed line marks the variant position. (**E**) Observed and predicted cell-type–specific scATAC-seq signal in the cell type with the largest ChromBPNet-predicted log-count difference between alleles, shown over a ±500 bp window centered on the variant. The black dashed line denotes the variant position. ENCODE SCREEN distal enhancer-like sequence (dELS) are shown as yellow bars. (**F**) ChromBPNet contribution scores for the reference and alternative alleles across a ±50 bp window centered on the variant. The highest-scoring transcription factor motif overlapping the variant, derived from the reference sequence, is indicated. The black dashed line indicates the variant position.

To further functionally annotate the identified rare variant caQTLs, we used ChromBPNet to predict the allelic-specific chromatin accessibility and assess whether rare variant associations could be explained by disruption of TF binding motif (**Methods**). We identified a rare variant, chr12:9650869:G:A, located 14.1kb upstream of *CLEC2D* within a distal enhancer annotated in the ENCODE cCRE database and marked by H3K27Ac signal across multiple cell lines (**Fig. 3C-D**). The scRNA-seq data from TenK10K show that this variant is associated with reduced *CLEC2D* expression across multiple cell types, with the strongest effect observed in CD4 TEM (**Fig. 3C**). *CLEC2D* encodes LLT1, a ligand for the NK cell inhibitory receptor CD161 that suppresses NK cell cytotoxicity^36,37^ and stimulates IFN-γ release^38^. I*n silico* predictions using ChromBPNet support this functional interpretation: the reference allele (G) is predicted to generate a strong, localized chromatin accessibility signal for peak chr12:9650649–9651305, whereas the alternative allele (A) markedly attenuates this signal (**Fig. 3E**). This peak does not have significant common caQTLs in any cell type. Contribution score analysis further indicates that the reference sequence contains a canonical *ELF1* binding motif (MA0473.4) centred on the variant (**Supplementary Table 9**), which is disrupted by the alternative allele, abolishing positive nucleotide contributions at this site (**Fig. 3F** and **Methods**). These results suggest that this rare G>A variant reduces chromatin accessibility at a distal regulatory element by disrupting an *ELF1* binding motif, leading to decreased *CLEC2D* expression.

Another example of a rare caQTL is chr19:16289808 (C>T) (**Supplementary Fig. 17**). The alternative (T; allele count = 1) located 34.8kb upstream of *KLF2* and overlaps with distal enhancer annotation in the ENCODE cCRE catalogue. The T allele is associated with outlier under-expression of *KLF2* across multiple cell types and is predicted by ChromBPNet to reduce chromatin accessibility at the corresponding peak (chr19:16289593–16290014) by weakening a *CREB1* binding motif (**Supplementary Fig. 17** and **Supplementary Table 9**). *CREB1* is a stimulus-responsive transcriptional activator that recruits CBP/p300 coactivators downstream of TCR and cAMP/PKA signalling, providing a plausible direct mechanism by which loss of *CREB1* binding at the T allele reduces enhancer activity and *KLF2* expression. A rare caQTL chr21:44932815 (C>T) associated with peak chr21:44932408–44933489. This rare variant is 735bp upstream of the transcription starting site of *ITGB2*, and the T allele is associated with outlier under-expression of *ITGB2* in many cell types, with the strongest reduction in NK cells (**Supplementary Fig. 18**). The T allele is also predicted to reduce the chromatin accessibility of the corresponding peak (**Supplementary Fig. 18**).

Together, these examples demonstrate that rare single-nucleotide variants can exert precise, mechanistically interpretable effects on chromatin accessibility through transcription factor motif disruption, with downstream consequences for the cell type–specific regulation of genes underpinning immune function and effects that are largely invisible to analyses restricted to common variation.

### Identification and characterization of cis-regulatory elements across cell types

To link chromatin peaks to genes and characterize their regulatory roles, we leveraged summary statistics from TenK10K caQTLs and eQTLs to perform colocalization across ∼3.7 million peak–gene-cell type pairs within ±1 Mb windows (**Methods**). Using *coloc*^39^, we identified 86,364 significant colocalization events by selecting those with a posterior probability of sharing a single causal variant (denoted as PP.H4) greater than 0.8, involving 31,688 peaks and 11,665 genes (**Fig. 4A** and **Supplementary Table 10**). These peaks are defined as candidate *cis*-regulatory elements (cCREs), enabling peak-gene linkage without requiring paired multiome data.

**Figure 4.**
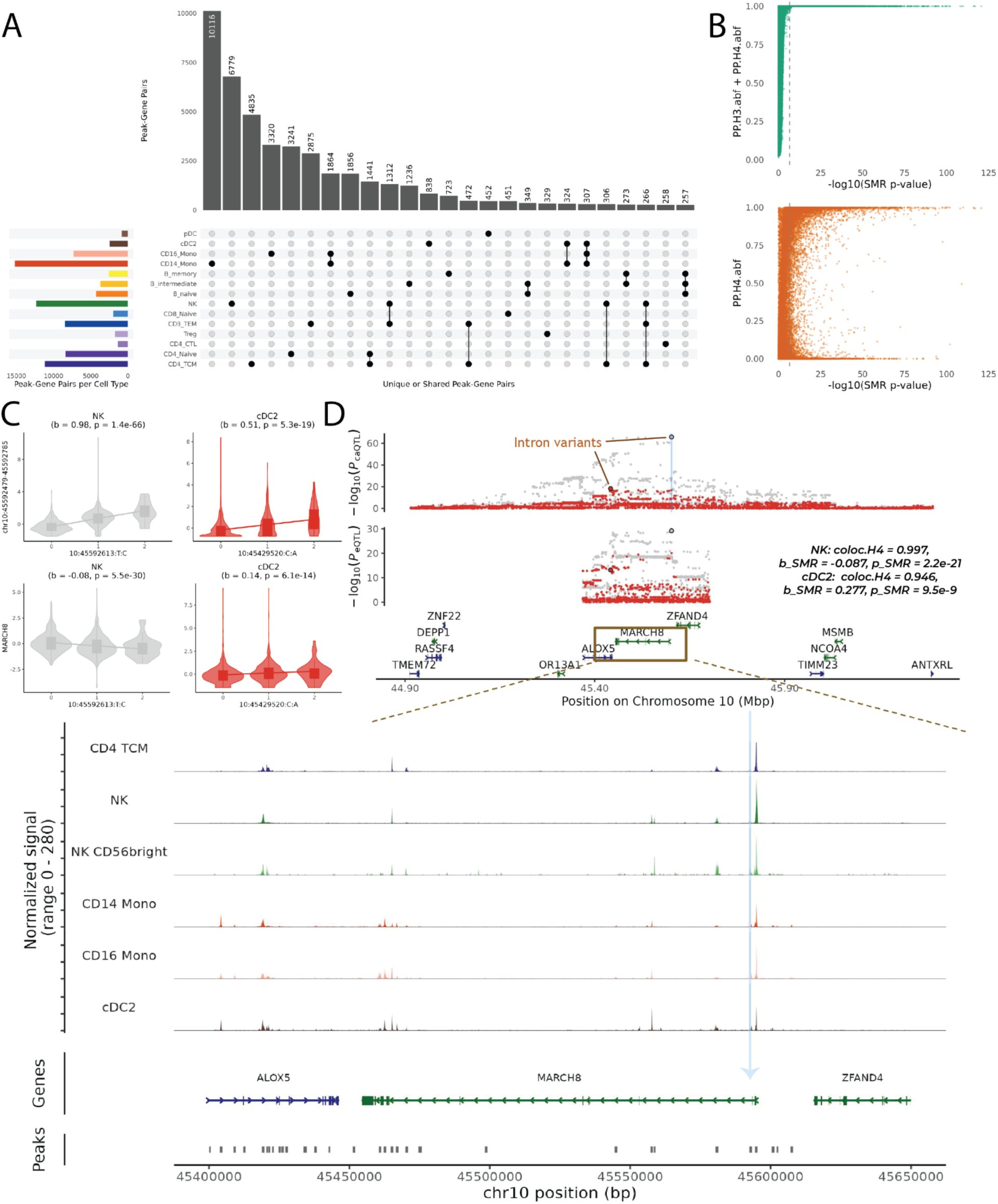
Characterization of candidate cis-regulatory elements (cCREs) across immune cell types. (**A**) Cell type-specific and shared cCREs identified by colocalizing caQTLs and eQTLs. The main panel shows the number of peak-gene pairs that are colocalized in or shared by multiple cell types. The left-side bar plot shows the number of colocalized peak-gene pairs (PP.H4 > 0.8) per cell type, grouped by similar lineage and in the same color scheme. (**B**) Comparison between *coloc* and SMR results for peak–gene pairs. Top: posterior probability of shared (PP.H4) or independent (PP.H3) causal variants. Bottom: concordance between coloc posterior probability (PP.H4) and SMR *p*-values, showing strong agreement between the two approaches. (**C**) Combined violin and box plots of the top variants for caQTL (chr10:45592479-45592785) and eQTL (*MARCH8*) in NK cells and cDC2. The x-axis indicates the three genotype groups of the top variant, and the y-axis represents the standardized pseudobulked values of chromatin accessibility and gene expression, respectively. The slope estimates and nominal *p*-value for the association are annotated. (**D**) Locus plot of the caQTL. The upper panel depicts the significance level of all the associated variants. The x-axis indicates the genomic location and the y-axis indicates -log_10_ of the nominal *p*-values. Two top variants are highlighted in black circles and the peak is represented as a blue vertical line in its location. The middle panel shows the gene location track, with genes on the forward strand as blue and reverse strand as green. The bottom panel shows the zoomed-in region of *MARCH8* locus and chromatin peak map in five different cell types. A zoom-in gene track and peak track are also included. In the peak track, each dash indicates an open chromatin peak. The skyblue vertical line indicates the peak, connecting to the caPeak (chr10:45592479-45592785) in the peak track.

The number of colocalized events (i.e., significant peak-gene pairs) varied by cell type, with the most abundant cell type CD14_Mono_ showing the highest (15,243), including 10,115 pairs unique to CD14_Mono_ and 1,864 shared with CD16+ monocytes (CD16_Mono_) (**Fig. 4A**). Cell type-specificity was evident: for instance, the inflammatory bowel disease (IBD) risk gene *IRGM*, which encodes an interferon-inducible autophagy protein, showed a regulatory effect via promoter chromatin accessibility (chr5:150846177-150847636) in CD8+ effector memory T cells (CD8_TEM_) but not in CD8+ naïve T cells (CD8_Naive_) (**Supplementary Fig. 19**). This suggests that *IRGM* promoter activity is dependent on memory T cell state, refining our understanding of the cell state–specific regulatory context of this established IBD risk locus. Together, these results demonstrate that integrating eQTL and caQTL data allows for precise identification of regulatory elements and characterization of their context-specific activity.

While *coloc* infers whether the two phenotypes share a common causal genetic variant, it does not estimate the direction of effect or the magnitude of the causal effect. To address this, we applied summary-based Mendelian randomisation (SMR)^40^, treating chromatin accessibility as the exposure and gene expression as the outcome (**Methods**). We tested 3,667,924 peak–gene–cell type combinations (limited to the same peak-gene-cell type paired tested in *coloc*), identifying 81,550 significant associations at *p*_SMR_ < 1.363 × 10^-8^ (i.e., 0.05 / 3,667,924) (**Supplementary Table 11**). The significance of SMR results was strongly correlated with *coloc* statistics: for example, in CD14_Mono_, nearly all (11,613/12,006) SMR-significant peak–gene pairs had a combined posterior probability (PP.H3 + PP.H4) > 0.8, suggesting independent or shared causal variants between chromatin accessibility and gene expression. Moreover, pairs with strong evidence for colocalization were far more likely to yield significant SMR results. The median PP.H4 for colocalized pairs with significant SMR results was 0.794, compared to just 0.023 for those without significant SMR results (**Fig. 4B** and **Supplementary Fig. 20A–B**). Similarly, the built-in HEIDI (heterogeneity in dependent instruments) test results aligned with *coloc*’s PP.H3, supporting the presence of distinct causal variants when applicable (**Supplementary Fig. 20C**). Of the 81,550 significant peak-gene pairs, 29,517 exhibited very low colocalisation support (PP.H4 < 0.01) but stong evidence for distinct causal variants (mean PP.H3 = 0.98). Amongst these, 25,433 (86.16%) also showed significant heterogeneity (*p*_HEIDI_ < 0.05).

Of 86,325 colocalization events that were also tested in SMR analysis, 38,329 (44.40%) were also supported by significant SMR tests, strengthening confidence in their causal regulatory role. These include 16,111 cCREs and 6,129 target genes in total. A subset of 753 events (467 unique peak–gene pairs) achieved PP.H4 = 1, which indicates the chromatin accessibility and gene expression share the same causal variant, were frequently observed across multiple related cell types (**Supplementary Fig. 21**). To better characterize the functional roles of the cCREs with causal inference support, we identified bi-directional cCREs, including: (A) 2,425 peaks with opposing regulatory effects on multiple genes in the same cell type, (B) 533 peaks influencing the same gene opposite effect direction in different cell types, and (C) 2,907 peaks influencing multiple genes with opposite effect directions in different cell types. These context-specific directionality patterns further underscore the complex logic of genetic regulation across diverse genes and cell types in the human immune system, such as bivalent chromatin^41^.

We highlighted a locus, where a chromatin peak chr10:45592479-45592785 shows a negative effect (*b*_SMR_ = -0.087, *p*_SMR_ = 2.2 × 10^-21^) on gene *MARCH8* in NK cells but a positive effect (*b*_SMR_ = 0.277, *p*_SMR_ = 9.5 x 10^-9^) in Conventional Dendritic Cell 2 (cDC2) (**Figure 4C-D**). In NK cells, the top caQTL variant (10:45592613: T:C) is an intron variant of *MARCH8* and located within the peak. Its minor allele shows an increasing effect on the chromatin accessibility (**Figure 4C**). On the contrary, the top caQTL variant of this peak in cDC2 is 10:45429520:C:A, an intron variant of an upstream gene, *ALOX5*. The two top variants are in high LD (*r*^2^ = 0.73), show concordant effects on chromatin accessibility, but opposite directions on gene expression, highlighting distinct regulatory mechanisms of the same cCRE in different cell types. This peak-to-gene pair also shows very strong *coloc* PP.H4 values (0.997 in NK cells and 0.946 in cDC2), suggesting that the chromatin accessibility and gene expression share the same underlying causal variant (**Figure 4D** and **Supplementary Fig. 22**). These results demonstrate the feasibility of combining summary statistics from caQTL and eQTL analyses to not only identify *cis*-regulatory elements, but also to estimate effect size and direction under different contexts.

### Colocalization of caQTLs and GWAS reveals regulatory effects missed by eQTL analyses

GWAS have identified tens of thousands of loci associated with complex traits, yet the regulatory mechanisms through which many of these variants exert their effects remain unknown. We hypothesized that a substantial subset of GWAS loci may colocalize with caQTLs, rather than eQTLs, particularly where the regulatory variant influences chromatin accessibility at a distance (i.e., >100kb away) from the promoter or enhancer of the target gene. To evaluate this, we performed colocalization analysis between caQTLs and GWAS loci, and separately between eQTLs and GWAS loci across 16 diseases and 44 blood traits (**Methods**). On average, 9.8% (from 4.5% to 22.6%) more GWAS loci colocalized (PP.H4 > 0.8) with caQTLs than with eQTLs alone (**Fig. 5A** and **Supplementary Table 12**). This pattern was consistent when applying SMR to identify putative causal genes or peaks, reinforcing the causal relevance of caQTLs (**Fig. 5A** and **Methods**). We divided the colocalization results (for both *coloc* and SMR) into four groups, whether the GWAS loci colocalize with both caQTL and eQTL, with eQTL only, with caQTL only, or with neither (**Fig. 5B**). Incorporating bulk eQTL datasets from GTEx^42^ and eQTLGen^4^ further increased the colocalization rates by 2.2%-21.4% (**Methods** and **Supplementary Fig. 23**). Using eQTLGen as the baseline, the addition of TenK10K eQTL and caQTL nearly doubled the colocalization rates for most diseases and blood traits (**Supplementary Fig. 24**), indicating that a substantial proportion of cell type-specific genetic regulation underlying GWAS associations is missed by bulk eQTL analyses. Notably, many of these colocalization signals are highly cell type-specific. Across diseases, an average of 27.7% of colocalizing loci were detected in only a single cell type and would have been missed by bulk or single cell type analyses. For example, in IBD, 14 of 72 (19.4%) caQTL colocalization events were unique to a single cell type, with detection strongly dependent on the cell type abundance (**Supplementary Fig. 25**). Together, these findings underscore the importance of resolving genetic effects at the level of individual cell types. To further investigate why some GWAS loci colocalise with caQTLs but not eQTLs, and to interrogate the remaining “missing regulation”, we performed a series of analyses testing alternative mechanistic hypotheses.

**Figure 5.**
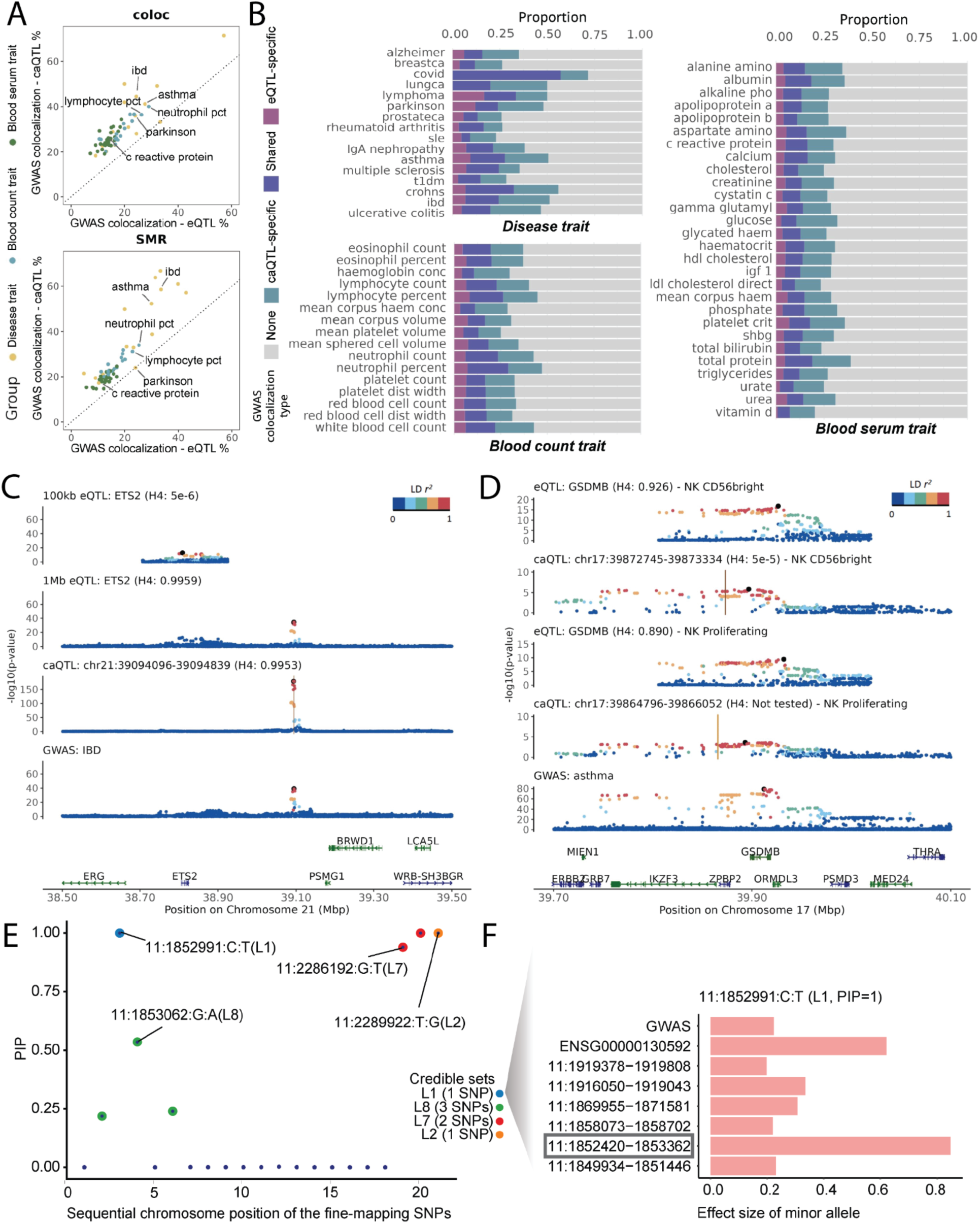
caQTLs explain additional GWAS signals beyond eQTLs by capturing distal regulatory effects. (**A**) Proportion of GWAS loci colocalising with caQTLs or eQTLs across 16 diseases and 44 blood traits. Trait categories are color-coded: complex diseases, blood serum markers, and blood cell counts (**B**) Distribution of GWAS loci by colocalization pattern. Stacked bars indicate the proportion of loci colocalising with caQTLs only, eQTLs only, both, or neither. (**C**) Locus plot at the *ETS2* locus illustrating colocalization results using eQTL (±100kb and ±1Mb window) and caQTL (±1Mb window) in CD14_Mono_. Dot color represents linkage disequilibrium (estimated by LD *r^2^*) with the lead GWAS SNP (21:39094644:G:A). Gene annotations (bottom) show genomic context and transcriptional orientation (blue: forward strand; green: reverse strand). (**D**) Locus plot at the *GSDMB* locus illustrating a GWAS signal that colocalizes with an eQTL, but not with a caQTL, in NK_CD56bright_ and NK_Proliferating cells_. (**E**) An example fine-mapping results using *mvSuSiE* in CD14_Mono_, identifying a credible set (L1) that affects IBD risk, gene expression and chromatin accessibility with posterior inclusion probability (PIP) = 1 for the causal SNP. The SNPs are ordered in sequential chromosome position rather than actual position for better visualization. (**F**) The zoom-in session shows the estimated effect sizes of the minor allele from the causal variant on chromatin accessibility, gene expression and IBD risk. The peak containing the L1 SNP is highlighted with a frame.

First, we examined whether chromatin accessibility captures more distal regulatory effects. Different eQTL studies chose various *cis*- window sizes (from ±100 kb to 1Mb) around the gene body, which may exclude distal enhancers harboring causal variants. For example, the *ETS2* locus is associated with IBD, and the GWAS lead SNP (21:39094644:G:A) lies ∼269 kb downstream of the *ETS2* gene in a gene desert annotated as a distal enhancer^43^. Colocalization between IBD GWAS and *ETS2* eQTLs was not detected when ±100 kb window is used because the GWAS lead SNP was not detected as an eQTL due to the distance > 100kb and therefore could not be assessed for colocalization. However, the same locus showed strong colocalization with a caQTL (chr21:39094096-39094839) in CD14_Mono_ (*q* = 2.1 × 10^-1^^55^) with PP.H4 = 0.995, as well as in CD16_Mono_ (*q* = 1.3 × 10^-57^) and cDC2 cells (*q* = 1.7 × 10^-10^) (**Fig. 5C**). Notably, this GWAS lead SNP resides within a caPeak in these cell types and the A allele is associated with decreased accessibility, consistent with prior literature^43^.

The GWAS SNP is in perfect linkage (*r^2^* = 1.0) with the top variant (21:39092834:CCTGTAATCCCA:C) in CD14_Mono_ and CD16_Mono_, and in high LD (*r^2^* = 0.87) with the top variant (21:39096912:G:A) in conventional dendritic cell 2 (cDC2). The eQTL analysis with a ±1 Mb window revealed significant colocalization with IBD GWAS (PP.H4 = 0.996) and strong SMR support (*p*_SMR_ = 4.2 × 10^-19^), where no association was detected under a 100 kb window (PP.H4 ≈ 0, *p*_SMR_ = 0.84). In total, we identified 35,382 such cases across genes, cell types, and traits, when expanding the window size from 100kb to 1Mb. This example highlights how the conventional ±1Mb eQTL window can capture distal regulatory effects that would be missed under a narrower window definition. Notably, even using eQTLs with a ±1 Mb window, caQTLs can still account for more GWAS loci by showing on average of 15.9% greater colocalization with them (vs 16.7% when using ±100kb window eQTLs) (**Supplementary Tables 12-13**). In addition, caQTLs that colocalize with eQTLs do not exhibit larger effect sizes than those that do not (**Supplementary Fig. 26**), arguing against a simple power-related explanation. These results suggest that window size may not be the primary factor in the “missing regulation”; instead, biological mechanisms such as multiple underlying causal variants or context-specific transcription factor activity may play a more important role. We further evaluated those caQTL-specific colocalization signals when using ±1 Mb eQTL window and observed 35.56% of loci with high PP.H3 values (> 0.8), indicating a substantial fraction of remaining caQTL-specific colocalization signals are potentially driven by multiple causal variants.

Second, we evaluated the impact of multi-causal-variant modelling on colocalization by comparing the conventional single-variant approach (coloc.abf) with SuSiE-based colocalization^44^ (coloc.susie) for IBD GWAS signals against TenK10K eQTL and caQTL data. For each cell type, we quantified genes and chromatin peaks showing strong evidence of colocalization (PP.H4 > 0.8) under each method, both across all tested features and within the subset jointly tested by both approaches. By modelling multiple independent signals per locus via credible sets, the SuSiE-based framework increased the proportion of colocalizing genes or peaks from 47.2% (77/164) to 53.0% (87/164) relative single-causal-variant model, consistent with the expectation that conventional coloc can miss signals at loci harbouring multiple causal variants (**Supplementary Table 14**). These results highlight the improved sensitivity of multi-signal colocalization approaches for resolving the cell type-specific regulatory architecture underlying complex disease associations.

Third, prior work^45^ has suggested that some caQTLs lacking corresponding eQTLs reflect enhancer priming. To test this, we leveraged the cellular diversity of our dataset by using naive CD4 T cell (CD4_Naive_) as a proxy for an unstimulated cell state. We first identified caQTLs in CD4_Naive_ cells that (i) overlapped with ENCODE v3 enhancer annotations and (ii) lacked a corresponding eQTL signal in the same cell type. We then assessed whether these caQTLs were also significant in effector CD4 T cell states (CD4_TCM_, CD4_TEM_, CD4_CTL_, and Treg), reasoning that the emergence of an eQTL in the effector state would be consistent with enhancer priming. To ensure comparability, analyses were restricted to variants that were significant caQTLs in both the naive and each respective effector state. Across the four effector cell types, the proportion of caQTL-only signals consistent with enhancer priming ranged from ∼5% to 15% (mean ∼10%; **Supplementary Fig. 27**). Extending this analysis to promoter regions yielded a slightly higher proportion (∼16%), consistent with promoter priming effects (**Supplementary Fig. 27**). While these results provide evidence supporting enhancer and promoter priming as a contributing mechanism, they account for only a minority of cases. Additional regulatory processes, such as post-transcriptional buffering or broader cell state-dependent regulation, are therefore likely to explain a higher fraction of caQTL-only signals.

On the other hand, some GWAS loci (∼5.16% on average) colocalized with eQTLs but not with caQTLs, which could be due to limited power for detecting caQTLs, as our eQTL dataset is twice as large as the caQTL dataset (1,925 vs 922) and ten times more features were tested for peaks than genes (440,996 vs 21,404). For instance, the asthma GWAS^46^ has 170 independent loci (**Supplementary Table 12**) and 16 of them only colocalized with eQTL results. One such locus is *GSDMB*, which regulates interferon-stimulated gene expression and airway inflammation upon respiratory virus infection^47^. While strong colocalization signals were observed between the *GSDMB* eQTL and the asthma GWAS in two relatively rare NK cell subtypes (NK_CD56bright_ and NK_Proliferating_), with PP.H4 = 0.93 and 0.89, no such colocalization was detected for caQTLs (**Fig. 5D**). Nevertheless, the overall association pattern and LD structure with the top variants appears consistent between the eQTL and caQTL results. The lack of caQTL colocalization is likely due to reduced statistical power in these rare cell types, resulting in weaker or non-significant caQTL signals compared to eQTLs. None of caQTL variants passed the genome-wide significant level so this peak was not tested in the SMR analysis between caQTL and asthma GWAS. We then used a relaxed significance threshold and the SMR analysis revealed suggestive associations at this locus with asthma in NK_CD56bright_ cells (*p*_SMR_ = 4.8 × 10^-6^) and NK_Proliferating_ cells (*p*_SMR_ = 3.5 × 10^-4^). Furthermore, based on the caQTL-eQTL SMR results in these respective cell types, it was confirmed that these peaks showed the strongest association with the *GSDMB* gene compared to nearby genes (e.g., in NK_CD56bright_ cells, *GSDMB p*_SMR_ = 3.5 × 10^-5^; all nearby genes had *p*_SMR_ > 0.05). We further show that caQTL signals at GWAS loci that colocalize only with eQTLs are substantially weaker than those that colocalize with both eQTLs and caQTL (**Supplementary Fig. 28A**), and that eQTL-only colocalization events are enriched in cell types with lower caQTL power (**Supplementary Fig. 28B**). These findings further support the possibility of a causal relationship between these regulatory peaks and the disease trait, which may be obscured by limited power in caQTL mapping.

### Multiomic fine-mapping resolves causal variants at immune disease loci

To pinpoint causal variants that simultaneously affect complex disease risk, chromatin accessibility, and gene expression, we applied a multi-trait fine-mapping framework^48^ based on summary statistics from univariate analyses (GWAS, caQTL, and eQTL) (**Methods**). We chose focal genes that are colocalised with GWAS (PP.H4 > 0.8) followed by caPeak selection for each focal gene, based on colocalization with eQTL (PP.H4 > 0.8). The focal region for each selected gene was set to ±1Mb of the gene body. We applied this approach to IBD GWAS data across all immune cell types, as it is one of the best-powered autoimmune diseases.

The method outputs credible sets designed to capture at least one causal variant per locus. Spanning 21 immune cell types, we identified 936 credible sets-phenotype pairs, of which 671 are single-variant credible sets, out of 916 that fit of focal gene-cell type pairs (**Supplementary Fig. 29** and **Supplementary Table 15**). As an illustrative example, the genetic variant 11:1852991:C:T, located within the 2kb upstream region of *LSP1*, an IBD-associated gene^49^, was identified as a causal variant for IBD risk. This SNP is strongly associated with the expression level of *LSP1* (nominal *p*_eQTL_ = 2.21 × 10^-1^^31^), and chromatin accessibility of peak chr11:1852420-1853362 (nominal *p*_caQTL_ = 9.77 × 10^-47^) in CD14_Mono_ cells (**Fig. 5E**). The colocalization results between caQTL and eQTL (PP.H4 = 1) also supported that the chromatin accessibility and gene expression shared one underlying causal variant. The minor allele effect of SNP shows positive effects for both chromatin accessibility and gene expression, which indicates the peak has a positive regulatory effect on the gene. This is also consistent with the significantly positive estimate in peak-gene SMR analysis (*b*_SMR_ = 0.47, *p*_SMR_ = 3.71 × 10^-35^) in CD14_Mono_. In addition, the top eQTL of *ETS2* (21:39094644:G:A) was also observed in a single-variant credible set, with a posterior in probability of 1 in CD14_Mono_ (**Supplementary Fig. 30**), consistent with the previous functionally prioritized results^43^, which could not be identified using GWAS only fine-mapping results due to high LD between candidate SNPs. This multi-layered regulatory effect highlights the power of joint fine-mapping to resolve the mechanistic basis of disease loci. By integrating eQTL and caQTL information, this approach provides a high-resolution view of how genetic variation shapes disease risk through coordinated regulatory effects on chromatin and transcription.

These findings underscore the value of caQTLs in resolving GWAS loci with previously uncharacterized regulatory mechanisms and in prioritizing candidate causal variants. While some GWAS signals colocalize with eQTLs but not caQTLs, on average ∼16.7% of loci colocalize exclusively with caQTLs. Incorporating caQTL, therefore, substantially expands the catalogue of regulatory variants, particularly those acting through multiple causal elements or context-dependent chromatin priming. Importantly, after accounting for all these factors, we achieve ∼60% colocalization GWAS loci (using IBD as an example), compared to ∼25% using TenK10K eQTL alone, the largest available single cell eQTL resource. This indicates that the ’missing regulation’ gap can be substantially reduced, although fully resolved. Further progress will require integrating multiple layers simultaneously, including multi-variant statistical modelling, expanded regulatory windows, chromatin-based detection of primed or context-specific regulatory states not captured by transcriptomic data alone, and the inclusion of disease-relevant tissues and patient-derived samples^50^.

### Genetic regulation of chromatin accessibility reveals cell state-dependent patterns along epigenetic age

To investigate whether genetic effects on chromatin accessibility vary with cell state, we applied EpiTrace^51^ to assign each cell a relative ‘epigenetic age’ based on chromatin accessibility across ∼126,000 age-associated genomic regions. When cells were ordered along this axis, we observed trajectories consistent with known hematopoietic differentiation pathways – for example, naive B (B_naive_) cells were classified as epigenetically younger than terminally differentiated plasmablasts (**Fig. 6A**). The inferred epigenetic age is not correlated with donors’ chronological age (**Supplementary Fig. 31**). These results confirm that epigenetic age serves as a biologically meaningful proxy for cell state and maturation. Using this metric, we tested for genotype-by-epigenetic-age interactions at previously identified caQTLs to identify caQTLs whose effects are modulated by epigenetic age cell-state dynamics. For each cell type, chromatin accessibility was modeled as a function of genotype, median epigenetic age, and their interaction, adjusting for relevant covariates (**Methods**).

**Figure 6.**
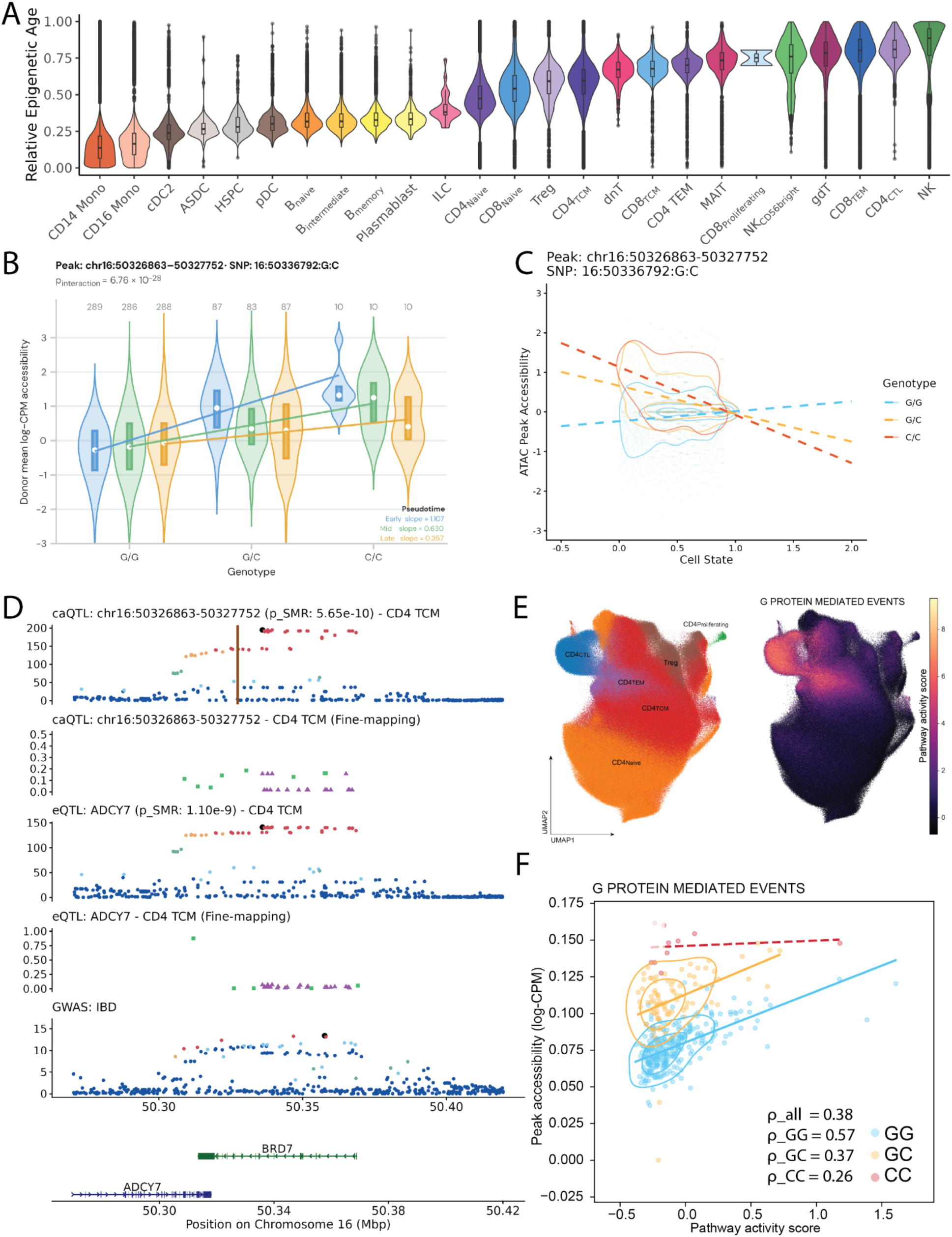
Genetic effects on chromatin accessibility vary with epigenetic age, revealing context-dependent regulatory patterns. (**A**) Distribution of relative epigenetic age across individual cells, grouped by cell type. Epigenetic age is derived from chromatin accessibility at ∼126,000 age-associated regions. (**B**) Example of a cell state-dependent caQTL (chr16:50326863-50327752) in CD4_TCM_ cells. Boxplot shows chromatin accessibility at a peak in the *ADCY7* locus (stratified by 16:50336792:G:C genotype and pseudotime group), illustrating decreased effect size in cells of the later pseudotime developmental group. The number above each violin plot indicates the number of donors in each group. (**C**) Density plot of the inferred pseudotime within the CD4+ T cell population (N = 493,520). The *x*-axis represents pseudotime, and the *y*-axis shows normalized chromatin accessibility peak chr16:50326863-50327752. Points are coloured by genotype group. (**D**) Locus plot integrating the eQTL for *ADCY7,* the caQTL for chr16:50326863-50327752 in CD4_TCM_, and IBD GWAS signals. The first four panels show caQTL and eQTL associations along with their fine-mapped results in CD4_TCM,_ as indicated in the subtitle. The vertical brown line marks the caPeak position. Point color denotes linkage disequilibrium (LD) with the lead variant (highlighted in purple). Two credible sets are indicated by a green square and a purple triangle. The fifth panel shows the SMR association with IBD GWAS (*p*_SMR_). In fine-mapping panels, the *y*-axis represents posterior inclusion probability (PIP) estimated by SuSiE, while in QTL and GWAS panels, the *y*-axis shows -log_10_ nominal *p*-values. The bottom panel displays gene annotations. (**E**) UMAP of CD4+ T cell population and pathway activity scores for G-protein mediated events. (**F**) Correlation between peak accessibility and pathway activity score for G-protein-mediated events at the pseudobulk level. Pearson’s correlation coefficient is estimated for all donors or the three genotype groups separately.

Across all tested lineages, we identified 3,080 significant caQTL-age interactions (FDR < 0.05; **Supplementary Fig. 32A**). The EpiTrace model includes 126k peaks, of which only 326 overlap with our significant csPeaks, indicating minimal overlap and suggesting that our results are unlikely to be biased by the model. These interactions reflect cell state–dependent penetrance of regulatory variants, with genetic effects that vary along the trajectory of cellular differentiation or activation. However, we acknowledge that differences in statistical power across cell types cannot be ruled out as a contributing factor to the observed cell-type specificity of these interaction effects. To further characterize these interactions, we first examined the direction of allelic effects. A significant bias was observed towards positive interactions, where the minor allele was associated with increased chromatin accessibility in epigenetically ‘older’ cells. Specifically, 54% of the significant interactions had a positive coefficient (*p* = 1.8 x 10^-6^, binomial test), suggesting epigenetic age additively influences the genetic effects on chromatin accessibility of those peaks.

A representative example is observed in NK cells at a caPeak (chr6:31243369-31243947) within the *HLA-C* locus (interaction effect = 1.75, adjusted *p*-value = 1 × 10^-5^). In epigenetically young cells, the top SNP (6:31245967:T:A) shows no association with chromatin accessibility, whereas in epigenetically older cells the alternative allele is associated with a marked increase in accessibility near *HLA-C* (**Supplementary Fig. 32B-E**). This suggests that the regulatory influence of this variant emerges or strengthens during NK cell maturation. This caPeak also shows a very high PP.H3 estimate (=1) in the *coloc* with rheumatoid arthritis GWAS as well as causal inference (*p*_SMR_ = 4.8 x 10^-9^ and *p*_HEIDI_ = 2.79 × 10^-28^). The extremely high PP.H3 value in *coloc* and the significant heterogeneity test in SMR suggest more than one causal variant at this locus, which is consistent with the significant conditional analysis and the presence of a secondary signal (6:31152239:G:A) in moderate LD (*r*² = 0.54) with the top caQTL SNP.

To resolve cell state-dependent genetic regulation of chromatin accessibility at higher resolution, we inferred pseudotime using Palantir^52^ and performed trajectory analysis within the CD4+ T cell compartment (**Methods**). The inferred pseudotime showed a modest correlation with epigenetic age (mean Pearson’s *r* = 0.45; **Supplementary Fig. 33**), indicating partial concordance in capturing shared aspects of cellular state while also reflecting distinct biological signals. We then applied the SAIGE-Dynamic model^34^ to test for caQTL-pseudotime interactions at single-cell resolution, rather than using median pseudotime in a pseudobulk framework (**Methods**). This analysis identified 209,189 significant interactions (FDR < 0.05), of which 57,308 (27.3%) lacked a detectable main-effect caQTL effects, highlighting the additional power of interaction-based models to uncover context-specific regulatory variation (**Fig. 6B**).

Notably, the number of significant interactions detected using pseudotime substantially exceeded those identified using epigenetic age, underscoring the single-cell approach’s greater sensitivity and resolution relative to pseudobulk analyses. This framework also revealed more pronounced interaction patterns. For example, in CD4⁺ T cells, the peak at chr16:50,326,863–50,327,752 shows a significant interaction with pseudotime, with marked differences in caQTL effect sizes (β) of SNP 16:50336792:G:C across cellular states (**Fig. 6B**). The effect is genotype-dependent: individuals with the G/G genotype exhibit a positive association between pseudotime and chromatin accessibility, whereas alternative genotypes show the opposite trend (**Fig. 6C**), indicating allele-specific modulation of chromatin accessibility along the trajectory (**Fig. 6D**). The caQTL at this peak shows significant positive SMR association with the eQTL of *ADCY7* in CD4_TCM_ (*b*_SMR_= 0.218, *p*_SMR_ = 2.88×10^-101^), which encodes a membrane-bound adenylate cyclase that catalyses cAMP synthesis in immune cells. *ADCY7* is a known ulcerative colitis risk gene^53^, and a recent functional study showed that *ADCY7* in primary human CD4⁺ T cells regulates the Th1/Th2 cytokine balance, with its loss-of-function shifting cytokine production toward a Th2 phenotype^54^. Our results suggest that the cell state-dependent, allele-specific accessibility at this peak may modulate *ADCY7* expression along the CD4⁺ T cell differentiation trajectory, potentially contributing to disease-relevant variation in CD4⁺ T cell function. Further Reactome pathway enrichment analysis using scDeepID [**URLs**] revealed four pathways, including G-protein-mediated events (**Fig. 6E**). Their pathway activity scores are positively correlated with peak accessibility, and the correlation strength varies across the genotype groups (**Fig. 6F**).

Taken together, these results highlight regulatory effects that emerge only within dynamic cellular contexts and help explain additional layers of genetic regulation not captured by conventional main-effect analyses.”

### Integrating multiome data and QTL priors identifies novel pathways in gene regulatory networks

Dissecting the molecular basis of complex traits requires understanding how genetic variants affect gene function within regulatory networks. A key challenge is the robust inference of gene regulatory networks (GRNs) from multiome data. While many methods rely on co-expression or co-accessibility patterns^55,56^, these approaches do not typically incorporate genetic variation, which can provide prior knowledge or causal anchors for network inference. To address this, we integrated caQTL and eQTL summary statistics with single-cell multiome profiles to guide peak-to-gene and GRN inference. We hypothesized that incorporating external genetic signals would improve both the accuracy and cell type resolution of inferred networks.

We first evaluated this approach in CD14_Mono_ cells using cell type-specific eQTLs from TenK10K (**Fig. 7A**). We applied the graph-based GLUE^18^ framework, which models peak–gene relationships using multiome data and can incorporate prior knowledge via regulatory scores (**Methods**). With eQTL guidance, the median GLUE score (scaled from -1 to 1) for peak-gene links increased substantially. For example, from 0.36 to 0.93 in CD14_Mono_ for pairs supported by regulatory evidence. We also recovered known enhancer–promoter interactions, such as a caQTL involving multiple peaks upstream of *COL1A2*, consistent with reported enhancer-driven regulation^31^ (**Supplementary Fig. 34**).

**Figure 7.**
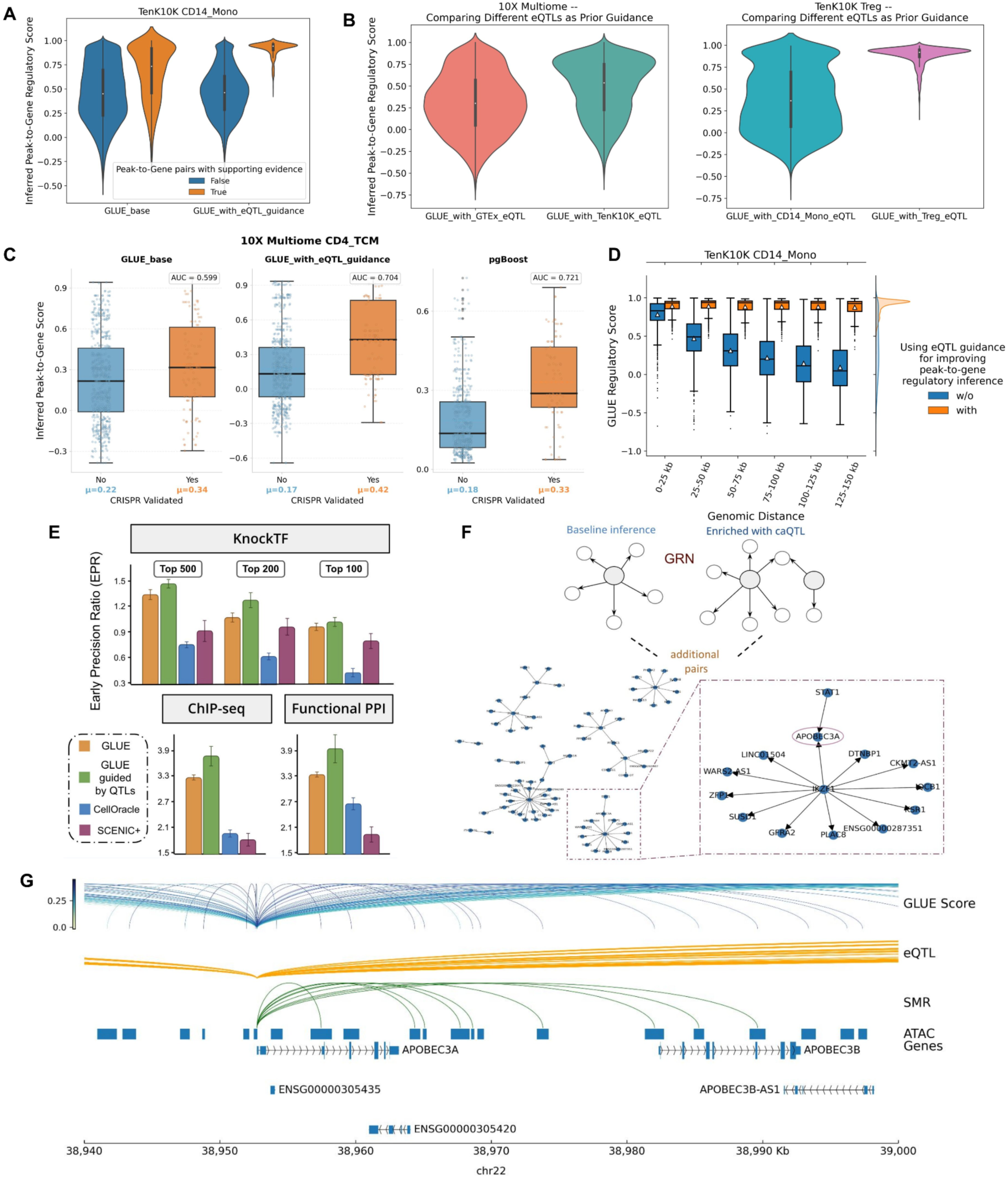
Integrative regulatory inference improves peak–gene mapping and expands gene regulatory network discovery. (**A**) Comparison of GLUE-inferred regulatory scores with and without cell type–specific eQTL guidance. Peak–gene pairs are stratified by the presence or absence of prior regulatory evidence. (**B**) GLUE scores for peak–gene pairs inferred using different eQTL sources. The first comparison contrasts GTEx bulk blood eQTLs with cell type–specific CD4Naive eQTLs in the 10x Genomics PBMC multiome dataset. The second compares two cell type–specific eQTL sets (CD14Mono and Treg) using Treg cells from the TenK10K multiome dataset. (**C**) Benchmarking of GLUE, GLUE with eQTL guidance, and pgBoost for peak–gene linkage inference. Pairs are stratified by CRISPR-based regulatory support, and performance is quantified by AUC. (**D**) Distribution of GLUE scores with (blue) and without (orange) prior regulatory guidance. (**E**) Benchmarking of GLUE against CellOracle and SCENIC+ for gene regulatory network (GRN) inference. Performance is evaluated using three independent TF–target validation datasets and quantified by the Early Precision Ratio. (**F**) Expansion of GRNs through integration of caQTL and SMR-based peak–gene links, identifying 128 additional TF–target gene pairs. Highlighted regulators include IKZF1 and STAT1, which modulate APOBEC3A via caQTL-mediated mechanisms and represent potential drug repurposing targets. (**G**) Locus plot for APOBEC3A showing SMR-based associations with chromatin accessibility and gene expression.”

To generalize this result, we compared different guidance types on the 10x Genomics PBMC 10k Multiome dataset. Using GLUE guided by GTEx v8 eQTLs (consistent with the original implementation) as a baseline, replacing bulk signals with cell type-specific eQTLs from major immune cell populations markedly improved peak–gene linkage inference. For example, a 0.30 increase in GLUE score in CD4_Naive_ for peak–gene pairs with supporting regulatory evidence (**Fig. 7B**). In contrast, guidance from Treg cells offered only modest improvement (from 0.30 to 0.34) (**Supplementary Fig. 35**). This likely reflects the limited number of Treg-specific eQTLs due to their lower abundance (∼1.46%) in the population, versus more abundant cell types like CD4_Naive_ (∼12.86%) and CD14_Mono_ (∼21.74%). This pattern was further supported when scaling the comparison to 12 cell types of varying abundance, which shows that, in general, guidance from more abundant cell types tends to produce larger increases in GLUE score. Notably, even when RNA and ATAC cell distributions diverged, for example, CD14_Mono_ cells represented only 2.3% of RNA-seq cells but 22.8% of ATAC-seq cells, accurate inference of *cis*-regulatory elements was still achievable, with a high average GLUE score of 0.77.

We next examined the importance of using matched QTL guidance from the same context for cell type-specific inference. By using 48,248 scRNA-seq and 52,042 scATAC-seq T_reg_ cells to construct baseline peak-gene links, we compared GLUE performance with eQTLs guidance from T_reg_ and CD14_Mono_ respectively (**Fig. 7B**). Despite CD14_Mono_ being ∼20× more abundant, using eQTLs guidance from T_reg_ resulted in a 0.57 increase in mean GLUE score, while using unmatched CD14_Mono_ eQTLs only showed 0.04 increase in mean GLUE score, both compared to the baseline inference. This observation confirms that cell type relevance outweighs cohort abundance in terms of inferring peak-gene links. Benchmarking analyses show that our approach achieves performance comparable to state-of-the-art methods (e.g., pgBoost^57^) for linking regulatory elements to target genes using three validation sets (GWAS, eQTL, and CRISPR), with additional gains in inference accuracy when incorporating QTL guidance (**Fig. 7C**, **Supplementary Fig. 36**, and **Methods**). Moreover, we observed that distal peaks (75–100 kb from TSS) benefited most from QTL-informed inference, reflecting greater ambiguity in their regulatory targets (**Fig. 7D**).

Finally, we extended the framework by integrating caQTL–eQTL SMR results alongside GLUE-inferred networks to improve transcription factor (TF)–target gene assignment. While GLUE leverages chromatin-expression co-variation with prior graphs, SMR uses shared genetic variants as instruments to infer causal peak–gene relationships, and as such, can leverage additional genetic association to pinpoint complementary regulatory interactions (**Supplementary Fig. 37** and **Methods**). Through comprehensive benchmarking of GLUE (**Methods**), we show that it, particularly when integrated with external QTL evidence, consistently outperforms other GRN inference methods across diverse validation datasets (**Fig. 7E**). This integration yielded 128 additional TF–target gene pairs (a 22.4% increase) (**Fig. 7F**), 66 of which are associated with GWAS loci for the 16 disease traits. For instance, at the *APOBEC3A* locus, GLUE alone produced weak regulatory scores (0.2-0.4), but SMR analysis identified a strong association with IBD risk (*p*_SMR_ = 1.44 × 10^-10^ in CD14_Mono_) and predicted *APOBEC3A* as a target of *IKZF1* and *STAT1* (**Fig. 7G**). Both TFs are druggable (*IKZF1* can be targeted using Lenalidomide and *STAT1* can be inhibited with Fludarabine), highlighting opportunities for therapeutic repurposing. To directly assess whether single-cell eQTL and caQTL data can support the identification of drug targets, we performed an enrichment analysis of target–indication pairs using the Open Targets Platform^58^ (OTP, version 25.12), and found that target–indication pairs supported by SMR evidence were enriched across all stages of clinical development (Phase I+, Phase II+, Phase III+, and Approved), with the strongest enrichment observed among approved indications. (**Supplementary Fig. 38** and **Methods**).

Together, these results show that integrating caQTL and eQTL data with multiome profiles enables more accurate, cell type-specific inference of regulatory networks. By combining data-driven co-variation with variant-mediated causal evidence, we uncover regulatory interactions—including druggable targets that would otherwise remain undetected in multiome data alone.

## Discussion

In this study, we present a comprehensive atlas of genetic effects on chromatin accessibility in human blood immune cells at single-cell resolution. By profiling over 3.5 million nuclei from 922 individuals using scATAC-seq and integrating with matched eQTL and WGS data, we identified 243,225 chromatin accessibility QTLs (caQTLs) across 28 immune cell types, with 22.8% of peaks showing genetic associations. Through statistical colocalization with matched single-cell eQTLs, we linked 31,688 of these peaks to 11,665 genes, defining candidate *cis*-regulatory elements (cCREs) and characterizing their regulatory activity in different cell types. Of these, over half (52.5%) showed statistically causal effects by Mendelian Randomization, providing causal evidence for their regulatory roles.

Our findings further improve the understanding of a long-standing challenge in human genetics: the limited overlap between GWAS loci and eQTLs. By integrating caQTL data, we demonstrate that chromatin accessibility accounts for a substantial fraction of previously uncolocalized GWAS loci. Across 16 diseases and 44 blood traits, 4.5% - 22.6% of GWAS loci colocalized exclusively with caQTLs, revealing distal regulatory effects not captured by standard eQTL analyses. We demonstrate this explicitly at the *ETS2* locus in IBD, where regulatory effects were only recovered when using caQTLs or expanding the eQTL window to ±1 Mb.

There are several biological explanations why those disease associations colocalize with caQTL but not eQTL. First, while a chromatin peak is open, it does not necessarily impact the targeted genes immediately. A genetic variant could affect the chromatin accessibility of the peak, which will be detected as a caQTL, but the peak is located in a poised or primed regulatory element, thus the gene expression levels are not variable (yet) across the genotype groups since the transcription has not started or activated. Second, transcription factor binding to the *cis*-regulatory elements could be specific to rare or developmental cell types, or require specific environmental triggers, and such effects might be identified as response-eQTL^59,60^. Third, in some cases, alterations in chromatin accessibility may not lead to detectable changes in gene expression because they impact downstream post-transcriptional regulatory activities, such as mRNA stability, splicing, or translational control, which can modulate RNA abundance independently of chromatin state^61–64^.

We further fine-mapped these regulatory interactions using multi-trait summary-based models, identifying 428 high-confidence causal SNPs that jointly affect chromatin accessibility, gene expression, and disease risk. For example, we highlight *LSP1*, where a single variant confers IBD risk through modulating its chromatin accessibility and expression levels in CD14+ monocytes. These analyses demonstrate how integrating regulatory layers can pinpoint functionally relevant variants and provide mechanistic insights into the biology of complex traits.

Beyond static regulatory maps, we discovered over 3,000 caQTL–epigenetic age interactions, revealing that genetic effects on chromatin accessibility are modulated by cellular maturation. These interactions were largely cell type-specific and frequently exhibited age-dependent amplification of genetic effects. For instance, a variant in the *HLA-C* locus exerted much stronger regulatory influence in epigenetically older NK cells, reflecting context-dependent penetrance along immune differentiation trajectories.

To enhance gene regulatory network (GRN) inference, we incorporated both eQTL and caQTL signals as prior guidance within the GLUE framework. This significantly improved peak–gene linkage accuracy, particularly when using QTL from the same cell type as the prior information. We further integrated causal peak–gene relationships derived from SMR, identifying 128 additional TF–target gene pairs, including actionable regulators such as *IKZF1* and *STAT1* at the *APOBEC3A* locus. These results demonstrate that leveraging genetic variation can substantially enrich multiome-based regulatory inference and reveal therapeutic opportunities.

Despite these advances, challenges and areas for refinement remain. caQTLs are mapped to peaks, which are not fixed genomic features and may vary between studies, depending on the scATAC-seq platform and peak-calling strategy. This complicates signal comparison and replication. Nevertheless, we observed high reproducibility across cohorts, even with differing scATAC-seq technologies and caQTL association models. Additionally, caQTL power is highly sensitive to the size of the genomic window used in association testing. While smaller windows (e.g. ±10 kb) reduce computational burden, they miss distal effects critical for accurate regulatory inference and colocalization. In some cases, variants nearly 1 Mb away exert strong influence on chromatin accessibility. This suggests that long-range regulatory interactions are also important for chromatin state variation in immune cells, likely mediated through 3D genome organization, enhancer–promoter looping, or higher-order chromatin domains^65^. Such distal effects are often missed in standard QTL studies, yet they may represent a substantial source of disease-associated regulatory variation, as evidenced by our finding that many GWAS loci colocalize exclusively with caQTLs. Our results underscore the need for more expansive genome windows (±1 Mb) when mapping caQTLs. Future work should focus on methods that can robustly detect long-range genetic regulation, capturing true distal effects without inflating technical noise from larger test windows.

Another challenge is cell type annotation in scATAC-seq data, particularly for rare populations where a gold standard does not exist. We demonstrate that utilizing multi-omics data as a bridging reference enhances annotation fidelity. We first tried to create a gene activity matrix by Signac and used the imputed RNA layer to transfer the label directly from the CITE-seq reference. However, the prediction accuracy is relatively low (more than half of the nuclei have a score < 0.5) compared to using a bridging dataset (mostly > 0.9), thus we chose the two-step annotation method as our main approach. In addition, the current annotation approach results in 28 cell types. While we consider this a balanced strategy for both annotation accuracy and caQTL discovery power, adopting a higher-resolution annotation reference^66^ could lead to the discovery of more nuanced cell type-specific regulatory effects. Future efforts should scale these references to at least 200,000 nuclei to ensure robust label transfer for human PBMCs, particularly in rare immune subsets.

Looking forward, beyond further increases in sample size, several priorities emerge. Chromatin accessibility captures only one dimension of regulatory biology. Future integrative efforts should incorporate additional layers such as histone modifications, DNA methylation, and protein expression to achieve a more comprehensive view of molecular regulation. In addition, reliance on resting cell states may obscure QTL effects that manifest only under stimulation or within specific cellular contexts. Moreover, extending caQTL mapping beyond blood to additional tissues and disease contexts will be essential to understand the generalisability and specificity of our findings^67–70^. Finally, incorporating other classes of genetic variation, such as structural variants and tandem repeats^20^, may reveal novel regulatory mechanisms that are currently overlooked by SNP- and indel-centric analyses.

Overall, this work establishes a foundation for utilising single-cell epigenomics and genetic variation to map regulatory mechanisms on a large scale. By connecting chromatin accessibility, gene expression, and complex trait variation across diverse immune cell types, the TenK10K caQTL resource offers a robust framework for mechanistic discovery and variant interpretation in human health and disease.

## Methods

### Sample collection and library processing

To build a chromatin accessibility atlas for human PBMCs, we collected blood samples from 1,042 individuals from the TenK10K phase 1 cohort^19^, which is a combination of 952 from the Tasmanian Ophthalmic Biobank (TOB) cohort, 64 from the BioHeart (BH) cohort^71^, and 26 from the Liquid Biopsy Biobank (LBIO) cohort. The 952 donors are obtained from a subset of the 982 donors of European ancestry in OneK1K^5^, the pilot cohort of TenK10K. The blood samples of TOB donors are processed in different batches from those used for processing scRNA-seq samples in Cuomo et al.^19^. The BioHEART cohort was recruited between March 29, 2016, and May 2, 2023, who presented for clinically indicated cardiovascular imaging or with acute ST-elevation myocardial infarction. The Liquid Biopsy Biobank (LBIO) for peripheral blood collection among patients with advanced cancer receiving intravenous therapy. Recruitment for LBIO commenced in December 2019 and is ongoing.

The ethics approval was approved for all three datasets: Tasmanian Ophthalmic Biobank [ethics protocol 2020/ETH02479], BioHEART study [ethics protocol 2019/ETH08376], and Liquid Biopsy Biobank [2019/ETH13113].

### Single-cell ATAC-seq

In total, 952 individuals from the TOB cohort were processed with 10x Genomics Chromium Next GEM single cell ATAC kit v2. This is the main dataset used from caQTL mapping. Briefly, cryopreserved PBMC samples were thawed, washed, counted and pooled by equal numbers of live cells per donor into 119 pools of 8 individuals. Nuclei were isolated and transposed before each pool was run across two single cell reactions, targeting either the same or different numbers of nuclei, ranging from 20,000 to 50,000. The specific number for each library can be found in **Supplementary Table 1**. A total of 238 Illumina sequencing libraries were generated, and sequenced at a read depth of ∼30,000 reads per nuclei using S4 flow cell kits on the Novaseq 6000.

### Multiome sequencing

The BioHEART and LBIO samples were processed with 10x Genomics Chromium Next GEM Single Cell Multiome ATAC + Gene Expression kit v1. The BioHEART multiome subcohort consisted of 64 individuals that were split into four pools of 16 individuals each, and the LBIO subcohort consisted of 26 individuals and was divided into four pools of 6-7 individuals each. As with the ATAC-only reaction, the cryopreserved PBMC samples were thawed, washed, counted and pooled by equal numbers of live cells per donor before nuclei were isolated and transposed. Each pooled cell suspension was then run across one single cell reaction. ATAC and Gene Expression libraries were generated from the same pool of pre-amplified transposed DNA and cDNA. ATAC and RNA libraries were sequenced independently at a read depth of ∼30,000 reads per nuclei using S4 flow cell kits on the Novaseq 6000.

### Whole genome sequencing and genotyping

DNA for whole genome sequencing (WGS) was obtained from blood samples using the Qiagen DNEasy Blood and Tissue Kit, with DNA concentrations measured by Qubit. Libraries were prepared using the KAPA PCR-Free kit and sequenced using Illumina NovaSeq 6000 at 30x coverage. More details of the quality control and processing can be found in the Cuomo et al.^19^. Genotype array data of all 952 TOB individuals were obtained from Yazar et al.^5^ and were used for demultiplexing only. 20 TOB individuals do not have WGS data, thus were excluded for caQTL mapping.

All the common variants, including SNPs and indels (minor allele frequency ≥ 5%), were subset from the WGS data and used for common variant caQTL mapping. Variants with minor allele count < 20 or missing genotype rate > 15% were also excluded. Rare variants (MAF < 5%) were extracted to perform rare variant caQTL mapping. The genotype array data generated using Illumina Infinium Global Screening Array in the OneK1K study^5^ was used for demultiplexing.

### Quality control and preprocessing

#### Read alignment (cell ranger ATAC)

Cellranger ATAC (v2.2.0) count was used to align sequencing reads to the reference genome (GRCh38 v1.2.0), the --force-cells parameter was set to 20,000, 40,000, or 50,000, depending on the targeted cell capture number for each sequencing library resulting in 9,457,411 nuclei in total (**Supplementary Table 1**). Using an older version without the --force-cells function (v2.1.0), which relied on the automatic cell calling algorithm, resulted in 5,639,779 nuclei (**Supplementary Table 2**). Initial peak calling was performed using the Cellranger built-in MACS2 method^22^. Cellranger ARC was called to used the multiome sequencing libraries (RNA+ATAC) for the BioHeart and LBIO cohorts, yielding a total of 105,887 nuclei.

### Demultiplexing + doublet detection

To assign the nuclei to each donor and identify doublets, we used *Vireo* implemented with *Demuxafy*^72^ (version: 2.1.0). Due to using the --force-cells parameter during cellranger count, a large number of nuclei of low quality were classified as unassigned during demultiplexing and were removed (1,155,771). For the TOB cohort, 3,644,297 doublets were detected and the average doublet rate was 37.0% across all 238 libraries, compared to 32.7% if no force cell was used (**Supplementary Table 3**). Two donors (comprising 10,003 nuclei) were not mapped to any donor’s genotype, thus 4,586,910 singlets from 950 donors were retained after this step. For the multiome sequencing results of the BioHEART and LBIO cohorts, in the ATAC layer, demultiplexing retained 41,248 singlets from 48 donors for BioHEART and 15,303 singlets from 12 donors for LBIO. One BioHEART library and two LBIO libraries with poor sequencing quality and demultiplexing performance were removed. In the gene expression layer, the data quality was also low, with only 11.9% of nuclei reliably assigned to a donor. Therefore, only the ATAC layer results were used for downstream analyses.

### Quality controls

Nuclei that failed the following quality control thresholds were excluded from analyses: number of peak counts > 5,000 and < 100,000, percentage of reads overlapping with peaks > 40, blacklist ratio < 0.01, nucleosome signal < 4, TSS enrichment score > 4. Seven sequencing libraries with low number of nuclei after quality control (< 300) were excluded, which resulted in the loss of one pool (8 donors excluded). After matching with whole genome sequencing data, 20 donors could not be matched and were removed. One pool (containing 8 donors) was excluded due to the extremely low number of nuclei. Seven libraries (S0268_2, S0301_1, S0302_1, S0303_1, S0304_1, S0304_2, S0331_1) were removed after all the QCs. The final data includes 3,472,552 nuclei from 922 TOB donors in 231 libraries (**Supplementary Fig. 1**). On average, each donor had 3,766 nuclei from scATAC-seq and 2,825 cells from matching scRNA-seq data.

### Cell type annotation by label transferring

To obtain a fine-grained resolution of cell types for PBMCs, we adopted a two-step strategy to annotate the scATAC-seq data. We first curated a multiome bridging dataset that combined 8 multiome datasets (99,507 nuclei), six 10x public datasets, BioHeart dataset, and LBIO dataset. The bridging dataset was merged into the same peak space by the “Reduce” method that merges all intersecting peaks. It was then annotated by label transferring from a CITE-seq reference (120,678 cells) provided by Hao et al.^21^ and the nuclei with a max prediction score < 0.3 were removed. In total, 95,348 nuclei (31 cell types excluding three non-PBMC types) were retained for the next step (**Supplementary Fig. 39-40**). After obtaining cell type labels for the bridging multiome dataset, each ATAC library was built with anchors to the ATAC layer in the multiome bridging dataset to obtain cell type labels. The number of each cell type for each library in TOB (**Supplementary Table 4**), and each donor in TOB, BioHeart, and LBIO cohorts were described (**Supplementary Tables 5-6**). The cell type annotation for each donor was also visualized in boxplot to check extreme outliers (**Supplementary Fig. 41**) and PCA plot annotated by how many cell types were outside the Q3 + 1.5 × IQR threshold (**Supplementary Fig. 42** and **Supplementary Table 16**). The two technical replicates for the same pool also shows reasonably high concordance (**Supplementary Fig. 43**).

The annotation strategy was designed to maximise both annotation accuracy and caQTL discovery power given the constraints of our study design. With approximately 2,825 nuclei per donor after quality control, accurate annotation of rare cell subtypes with an abundance below ∼0.1% is not statistically reliable, as many such types would be represented by fewer than three nuclei per donor on average. This per-donor yield is partly a consequence of the pooled library design (eight donors per pool), which places a practical ceiling on nuclei recovery. Furthermore, caQTL discovery power scales with the number of nuclei per donor per cell type (**Figure 2B**), such that many finer-grained subtypes would lack sufficient representation to support meaningful caQTL mapping. Together, these constraints motivate the use of 28 cell types as a level of granularity that balances annotation fidelity with statistical power.

### Verifying cell type annotation with imputed scRNA-seq data

After obtaining cell type labels, we checked canonical gene markers for each cell type. We used the gene activity score generated in the previous step for each library. Then we visualized the expression levels of the gene markers used in the Seurat^21,73^/Azimuth reference (**URLs**) for CD14_Mono_ and cDC1, confirmed the canonical markers have enriched scores in the corresponding cell populations (**Supplementary Figure 40**). We further visualized the gene activity scores of the used markers for all 26 cell types for the main scATAC-seq data (**Supplementary Figure 44**).

### Recalling peaks within each cell type

For each cell type, we recalled peaks from the raw fragment files using the MACS3 tool^22^ integrated into SnapATAC2^23^. The total number of peaks called in each cell type ranged from 427 (CD8_Proliferating_) to 269,371 (CD16_Mono_), with a median of 170,292 (**Fig. 1F**). The peaks were then merged using the ‘reduce’ function from Signac^74^ to define a consensus list of peaks. In total, we have 440,996 unique peaks for the merged object, and the min, median, and max widths are 201bp, 499bp, and 5,545bp. We matched the peak regions against two comprehensive datasets: ENCODE v4 [ref^75^] with 2,348,854 peaks, and a whole-body scATAC-seq atlas^24^ with 1,154,611 peaks, as well as the 10X 10k PBMCs ATAC v2 Chromium Controller dataset (164,487 peaks and 10,246 nuclei). After matching, 44,015 peaks (9.98%) did not overlap with any of these three datasets (**Supplementary Table 7**). The novel peaks are likely due to the increase of sample size so we have more power to call peaks with low accessibility, as supported by the observation that the donor-level sparsity of novel peaks is consistently higher than those of known peaks (**Supplementary Fig. 15**). ChromBPNet also shows lower prediction accuracy for novel peaks compared to known peaks (**Supplementary Fig. 14**).

The peak list was annotated based on the functional categories curated by overlapping the pre-annotated regions. In total, 437,561 peaks were mapped to 65 different functional categories. One peak could match multiple categories and when we calculated the proportion of each category we used all the matched pairs to calculate the proportion of each matched category. For ENCODE v3 annotation^76^, there were five main categories - PLS (promoter-like signature), pELS (proximal enhancer-like signature), dELS (distal enhancer-like signature), DNase-H3K4me3, and CTCF-only.

### Dimensional reduction and visualisation

We used EpiScanpy^77^ to obtain a dimension reduction space for the scATAC-seq data. Episcanpy is a toolkit for the analysis of single-cell epigenomic data including scATAC-seq. We combined the quality controlled scATAC-seq data into a single anndata object without removing the 20 donors with unmatched whole genome sequencing data (3,546,117 cells with 440,996 peaks), followed by binarisation and feature selection of top 50,000 peaks based on EpiScanpy’s inbuilt feature variability score. We then further normalize the data such that the total count of each cell equals the median of total counts for all the cells before normalisation. We further performed PCA with Scanpy^78^ as the dimensional reduction of the dataset and the cell clustering pattern is as expected (**Supplementary Fig. 45**). The scATAC-seq data was integrated with scRNA-seq in TenK10K by projecting into a co-embedding space (**Figure 1B**). There is a cluster with limited overlap between two layers, but it is due to the different proportion of monocyte population in two datasets because the blood samples were processed in different batches. More detailed explanation can be found in the *<u>Supplementary Note 1</u>* of TenK10K flagship paper^19^.

### The caQTL mapping

Overall, after the nucleus-level and sample-level QCs, a total of 3,472,552 nuclei of scATAC-seq from 922 individuals, 5,438,679 cells of scRNA-seq from 1,925 individuals (including the 922 individuals with scATAC-seq), and 96,728 nuclei of multiome data from 63 individuals (independent from other two cohorts) were used for subsequent analyses resulting in 9 million single cells/nuclei (**Fig. 1B**).

### Cell type-specific caQTL mapping

To map genetic variants to the variation of chromatin accessibility across individuals, we ran an association test between genetic variants (SNPs + indels) and chromatin accessibility levels in each of the 28 cell types. The peak counts were aggregated (by summing all nuclei for one donor) as a pseudobulk matrix for each cell type. This was done in each repeat separately. In each repeat, we corrected the GC content bias using EDAseq’s ‘withinLaneNormalization’ function^79^. Then we removed peaks with > 95% zero count across donors. We converted the count data to counts per million (CPM) and then performed z-score normalisation for all the peaks. We merged the normalized matrix from two repeats by taking the average of the *z*-scores for overlapped peaks. For the unique peaks, keep the original *z*-scores. Finally, we re-standardize each peak across all donors. The standardized matrix was used to generate chromatin accessibility principal components (PCs). We conducted a series of sensitivity analyses to optimize the parameters used for association mapping, including peak calling methods, normalization methods, window size, and number of PCs (**Supplementary Fig. 46-48**). Although choosing a smaller window size, such as 10kb, resulted in more significant associations, many associations outside the 10kb window would be missed and the number of tested variants overlap with other QTL dataset would be limited. To generate consistent SNP-peak pairs between the caQTL discovery and downstream colocalization analysis, we decide to use ±1Mb as the default window size.

The association model fits multiple covariates, including sex, age, genotype PCs, and chromatin accessibility PCs. The final number of donors per cell type ranges from 120 to 922. The two rarest cell types—CD8_Proliferating_ (3 donors) and innate lymphoid cells (ILCs) (19 donors)—were not tested due to the limited donor size. The number of PCs fitted in the model was decided by the Elbow method in R package *PCAForQTL*^80^. Each peak was tested against all the SNPs located within the ±1Mb window around the peak center, and the number of SNPs tested for each peak ranged from 293 to 30,942, with a median of 6,067. For each peak, TensorQTL^30^ runs 10,000 permutations to correct the *p*-value by a beta-approximation strategy. Then, the peak-level correction was conducted for each chromosome, and a significant caQTL was decided based on *q*-value < 0.05. GPU acceleration was utilized to speed up the association tests, which significantly reduced the computational time given the large number of peaks.

Since the scATAC-seq were measured twice for each pool, we also ran the caQTL association for each repeated library separately and then compared the caQTL signals between the two runs. The significant level, quantified by -log_10_(*q*-value), and effect sizes are largely consistent (Pearson’s correlation > 0.90) across cell types except for those rare cell types with limited power (**Supplementary Fig. 49**).

### Defining the cell type-specificity of caQTL effects

To quantify cell type-specificity of caQTLs, we assessed different models to explain the specificity using the approach described in Cuomo et al.^19^ (**Fig. 2C** and **Supplementary Fig. 6**). We defined five possible scenarios that can occur for a given caQTL identified at q-value < 5% in one cell type (cell type A) when compared pairwise with another cell type (cell type B):

● Scenario 1 (specific caPeak): The caPeak associated with the caQTL is not sufficiently accessible (<1% of cells) in cell type B.
● Scenario 2 (specific caQTL): The caPeak is sufficiently accessible in cell type B, but no caQTL is identified.
● Scenario 3 (distinct caQTL): Different caQTL variants in low linkage disequilibrium (*r*² < 0.5) are identified for the same caPeak in cell types A and B.
● Scenario 4 (shared caQTL, concordant effect): The same caQTL variant or variants in high LD (*r*² ≥ 0.5) are identified for the same caPeak in both cell types, with a concordant direction of allelic effect (i.e., the same directional impact of the minor allele on chromatin accessibility).
● Scenario 5 (shared caQTL, discordant effect): The same caQTL variant or variants in high LD (*r*² ≥ 0.5) are identified for the same caPeak in both cell types, but with a discordant direction of allelic effect.

### Functional annotation of caQTLs

There are 243,225 caQTLs, spanning 100,486 peaks and 158,251 unique top variants. Same to the annotation of chromatin peaks, we overlapped the caPeaks with the five cCRE categories in ENCODE v3, as well as the top variant for each peak (**Fig. 2F**). The annotation of caPeaks are further divided into five subgroups based on the peak width.

### Fine-mapping of causal variants for caQTLs using SuSiE

We performed fine-mapping analysis for all caPeaks using SuSiE^28^. For each peak, we considered all variants located within a 1 Mb cis-window and calculated the corresponding variant correlation matrix using PLINK^81^ (v1.9). The z-scores were obtained from TensorQTL summary statistics, and the sample size N was defined as the number of donors containing the corresponding cell type. These summary statistics were supplied to the ‘susie_rss’ function with L = 10. The analysis yielded credible sets for each peak and marginal posterior inclusion probabilities (PIPs) for individual variants.

### Replication of caQTLs in published and in-house dataset

To replicate our identified caQTL signals, we collected caQTL results from two published studies^16,29^. Because different cell type annotations were used in those published studies, we count a caQTL being replicated if it reaches significance in the closest cell type. In other words, if at least one of our cell types contained a significant caQTL that overlapped with theirs, we counted it as being replicated. The replication of direction of the caQTL effects were also quantified for Benaglio *et al*.^16^. Since peak definitions differ across studies, we counted a replication if the peak regions overlapped between datasets, which might inflate concordance estimates. Conversely, fewer than 10% of our caQTLs replicate in prior studies, as expected given the limited statistical power of those earlier cohorts. We also conducted caQTL mapping for a combined multiome dataset, which comprised 60 donors from two cohorts (BioHeart and LBIO) after QC. The cell type annotations, pseudobulking, and caQTL association followed the same protocol as the main caQTL analysis for the TOB cohort.

We also used WASP^32^, which applies a Combined Haplotype Test (CHT) that tests for genetic association with a molecular trait using counts of mapped and allele-specific reads. The CHT jointly models total read depth within the test region and allelic imbalance at phased heterozygous SNPs in the same region, while accounting for overdispersion in the read count data. We applied this analysis to 23,583 cells from 15 donors spanning 26 cell types. Prior to association testing, read depths were corrected for GC content and sample-specific peakiness, and heterozygote probabilities were updated using allele-specific read counts to account for potential genotyping error. 5 cell types (ASDC, CD4_Proliferating_, CD8_TCM_, cDC1, and NK_Proliferating_) were excluded from downstream analysis because they did not meet the minimum read count threshold of 50 and allele-specific read count threshold of 10.

To assess concordance between population-level and allele-specific estimates of caQTL effect sizes, we compared results from TensorQTL and WASP chromatin haplotype test (CHT) for each cell type. Significant caQTLs were defined as SNP–peak pairs with a *q*-value < 0.05 from TensorQTL *cis*-nominal results. For each significant peak, we retained only SNPs physically located within the peak boundaries and present in the WASP CHT output. TensorQTL effect sizes are reported as the regression slope (change in normalised accessibility per copy of the alternative allele). WASP effect sizes were computed as log_2_(BETA/ALPHA), where BETA and ALPHA represent estimated accessibility from the alternative and reference haplotypes, respectively. Prior to comparison, allele orientations were harmonised across the two methods. SNP–peak pairs with discordant or ambiguous allele assignments were excluded. On average, 5,347 SNP–peak pairs were retained, ranging from 0 for plasmablast to 26,916 for CD14 monocytes. Spearman rank correlation between TensorQTL slopes and WASP log_2_(BETA/ALPHA) values was computed genome-wide and within subsets of WASP-nominally significant SNPs (*P* < 0.05 and *P* < 0.01). Effect size estimates with non-finite values or extreme value of | log_2_(BETA/ALPHA) | > 10 were removed when calculating the correlation.

### Mapping rare caQTLs using SAIGE-QTL

We assessed the effects of rare variants on single-cell chromatin accessibility using the set-based test implemented in SAIGE-QTL. All SNPs and indels with minor allele frequency (MAF) < 5% and missingness <15% were included. For each peak, we tested the combined effects of variants within a *cis*- region defined as ±100 kb around the peak center. We used a different window size here because it maximized the number of caQTLs in a sensitivity analysis which tested chromosome 1 and CD4_Naive_ cells (**Supplementary Fig. 16**). Following the SAIGE-QTL framework, both burden and SKAT tests were performed, and peak-level *p*-values were combined using the Cauchy method, similar to the SAIGE-QTL paper. Consistent with the procedure applied to common variants above, we controlled for multiple testing using the *q*-value method and considered results significant at FDR < 5%.

To rule out the scenarios where rare signals are induced by common variants that in LD with the rare variants tested, we further performed a conditional analysis that allows for conditioning on the nearby significant common caQTLs. For each peak, we conditioned on the common variant with the lowest *p*-value within a 100 kb cis-window centred on the peak, as identified from the TensorQTL results. Following stepwise conditional analysis, 53,195 significant rare caQTLs remained significant in the set-based test.

### Regulatory effect prediction using ChromBPNet

#### Processing of single-cell ATAC-seq data

We first normalized the sequencing depth and aggregated the 238 scATAC-seq libraries. Then, we created pseudobulk fragment files by pooling all fragments from each predicted cell type. Next, we randomly split the fragments into two pseudo-replicates, and called peaks with ENCODE ATAC-seq pipeline v2.2.3 (https://github.com/ENCODE-DCC/atac-seq-pipeline) with default parameters. We used the overlap peak set for all downstream analyses.

### ChromBPNet Bias model training

To select the initial hyperparameters for the bias model, we partitioned the data into train, validation (chr8 and chr20), and test (chr1, chr3, chr6). Next,we trained bias models in three cell types with a range of available cells, including CD14_Mono_ (N = 767,874), NK (N = 432,705), and B_naive_ (N = 152,441) with bias threshold factor from 0.1 to 1 at 0.1 interval. We picked the best bias model of each cell type based on three criteria: (1) The Pearson’s *R* between predicted and observed non-peak counts was high. (2) The Pearson’s *R* between predicted and observed peak counts was low, but greater than -0.3. (3) The model only captured bias motifs (such as TN5/DNase-I) and did not capture any real TF motif in TF-MODISCO derived motifs.

Next, we trained ChromBPNet bias-factorized model^33^ using the best-performing bias models from each of the three cell types. We then selected the final bias model based on two criteria: the highest Pearson’s R between predicted and observed peak counts, and lowest Pearson’s R between predicted and observed non-peak counts while ensuring the model’s Tn5 bias motif was corrected.

Finally, we trained the final bias model (NK cell with bias threshold factor of 1) with 5-fold cross-validation. We followed the chromosome partition scheme used in the ChromBPNet method^33^, where each chromosome is used as a test set exactly once across the folds.

### ChromBPNet bias-factorized model training

We trained ChromBPNet models in 25 out of 28 cell types, except for CD8_Proliferating_ (4 cells), ILC (33 cells), and CD4_Proliferating_ (145 cells), with 5-fold cross-validation using the final bias model selected in the previous step.

### ChromBPNet variant effect prediction

For all top SNPs associated with a peak, we generated a 2,114bp (ChromBPNet input length) region centered on the variant. Then, we computed the cell type-specific variant scores as the log_2_ fold change in the predicted total counts of the alternative allele versus the reference allele using the cell type-specific ChromBPNet bias-factorized models. The same prediction was also performed using the fine-mapped SNPs with high PIP scores in each peak and the prediction accuracy was compared across using different PIP thresholds (**Supplementary Fig. 13**).

### Transcription factor motif matching

To assess motif matches of the caQTL variants, we used the JASPAR 2026 vertebrate database^82^. For each variant, we computed contribution scores and performed motif discovery using Tangermeme (v0.5.1). We then assigned the best-matching TF based on the most significant motif p-value, near the variant position.

### Colocalize caQTL and eQTL to generate peak to gene links

To identify the links between peak and gene, we performed colocalization analysis using *coloc*^39^ by integrating the eQTL and caQTL summary statistics. We test all the significant caPeaks against genes within ±1Mb distance from the peak center. To keep a consistent variant list between different colocalization analyses, we excluded all the indels in the summary statistics and only focused on SNP variants. The *coloc* results indicate the posterior probability of five different models: PP.H0 : no association with either trait; PP.H1 : association with trait 1, not with trait 2; PP.H2 : association with trait 2, not with trait 1; PP.H3 : association with trait 1 and trait 2, two independent SNPs; PP.H4 : association with trait 1 and trait 2, one shared SNP. In our case, trait 1 is the chromatin accessibility and trait 2 is gene expression.

Although a significant colocalized event suggests a peak to gene association, it does not necessarily inform the directionality or effect size. To provide causal inference support for peak-gene pairs we identified through *coloc* analysis, we further adopted the Mendelian Randomisation framework to infer causal association of the caPeaks on nearby genes using *OmicsSMR*^83^. We first curated the caQTL and eQTL summary statistics into the MatrixEQTL format, and accordingly generated the BESD files as inputs for the SMR analysis. To select a significant peak for testing, we applied a nominal *p*-value threshold of < 5 × 10^-8^. This threshold for selecting peaks is much more stringent than using *q*-value < 0.05, so the number of peak-gene pairs tested in *OmicsSMR* will be much smaller than *coloc*. For example, in CD14_Mono_, 740,024 pairs in *coloc* and 191,423 pairs in OmicsSMR were tested. After taking the overlap, 190,494 pairs were kept so the *OmicsSMR* pairs are literally a subset (99.51%) of *coloc* pairs. When comparing two methods, we only focus on the overlapped pairs.

### Colocalization of caQTL, eQTL and GWAS

To further understand the epigenetic regulatory mechanisms of GWAS associations, we conducted colocalization between caQTLs and GWAS associations from 16 complex diseases using *coloc*, as well as 25 blood serum markers and 19 blood count traits, following the same traits tested in Margoliash et al.^84^. The *coloc* results between eQTL and GWAS were acquired from Cuomo et al.^19^. For further comparison, we also obtained bulk eQTL summary statistics from eQTLGen^4^ (*N* = 31,684) and GTEx v10 whole body tissues^42^ (*N* = 70 ∼ 816) and ran the same colocalization analysis. Similar to *coloc* analyses, we also ran SMR analysis between eQTL and GWAS. In SMR between eQTL and GWAS, we tested the gene in which the top eQTL has a nominal *p*-value < 5 × 10^-8^. The same SMR analysis was performed between caQTL and GWAS.

To evaluate how much GWAS signals are colocalized with eQTLs and caQTLs, we adopted a two-step approach. First, we performed clumping analysis to select independent loci for each GWAS, using a significance threshold of *p*-value < 5 × 10^-8^, an LD *r*^2^ threshold < 0.01, and a window size of ±1Mb. Next, we examined the raw eQTL and caQTL summary statistics, along with the outputs from *coloc* and SMR analyses. For each GWAS locus, we compiled a list of genes and peaks with significant QTL associations (nominal *p*-value < 5 × 10^-8^). A locus was considered colocalized with eQTLs or caQTLs if at least one gene or peak from the lists met the following criteria: for *coloc*, PP.H4 > 0.8; for SMR, *p*_SMR_ < 5 × 10^-8^. The results from all cell types were subsequently aggregated and used to calculate the overall proportion of GWAS loci colocalization (**Fig. 5A-B**).

Conventional *coloc* function assumes a single causal variant when assessing colocalization between two phenotypes. However, in practice, both phenotypes may harbor multiple causal variants. To relax this assumption, we integrated SuSiE, testing all pairs of credible sets across the two phenotypes, as implemented in the coloc.susie method^44^. Specifically, SuSiE fine-mapping was performed separately for each phenotype, and each pair of credible sets was subsequently evaluated using conventional coloc. If any pair showed significant evidence of colocalization (PP.H4 > 0.8), the two phenotypes were considered to colocalize.

### Multi-trait fine-mapping for identifying putative causal variants

We applied *mvSuSiE*^48^, a variable selection method for multivariate linear regression to identify the putative causal variants that simultaneously affect disease risk, gene expression and chromatin accessibility. Specifically, we selected genes that significantly colocalized with GWAS (PP.H4 > 0.8) as the focal genes. Then for each focal gene, we selected caPeaks colocalizing with eQTL (PP.H4 > 0.8) with ±1Mb region of the gene body. Then we extracted the summary statistics from the eQTL, caQTL mapping and GWAS and constructed the z-score matrix where rows represent different SNPs and columns represent different phenotypes. We also set up canonical covariance matrices for all of the remaining phenotypes with mashr^26^, as well as the LD matrix of the SNPs in the focal region with PLINK^81^. We then fit an mvSuSiE model with input as the z-score matrix, covariance matrices and the LD matrix.

The key output of a fitted mvSuSiE model includes a list of credible sets that are intended to capture at least one causal SNP; a “posterior inclusion probability” (PIP) score quantifying the probability of a SNP is causal; and posterior estimates of SNP effects on each phenotype. We used mvSuSiE’s local false sign rate as a phenotype-level significance measurement to define the phenotypes affected by the credible sets (cutoff = 0.01). Collectively, we identified 1,150 credible sets that at least affect one layer of phenotypes (**Supplementary Table 15**), and we focused on the credible sets that simultaneously affect disease risk, gene expression and chromatin accessibility.

### Genetic interaction with epigenetic age

#### Modelling cell type-specific genetic interactions with epigenetic age using EpiTrace

Epigenetic age was calculated for each cell using the R package EpiTrace^51^. First all pools were merged then each annotated cell type was subset to an individual object. For each object the top 200,000 chromatin features were selected using the function *sc.pp.highly_variable_genes* from Scanpy. For scalability with R, we acknowledge that the inference of Epigenetic age is computationally infeasible to apply to the whole datasets. Each individual cell type object was capped at 250,000 cells - if any cell type object surpassed this threshold it was randomly subset down to 250,000 with an even number of cells per individual.

Epigenetic age was derived by measuring chromatin accessibility at ∼126,000 peak regions associated with chronological and mitotic age. The main idea is that as cells age, chromatin accessibility heterogeneity at those regions decreases serving as a reference model to infer relative age of a cell in the context of other cells. To test for interactions between epigenetic age and cell type specific caQTLs, the relative epigenetic age of each cell was calculated per cell type. Once epigenetic age was calculated for each cell, the median epigenetic age for each individual was used as an interaction term while also fitting additional covariates as listed previously. Significant peak-caQTL pairs were fit to the linear mixed effects model with peak as a function of the associated variant in addition to covariates listed previously.

### Inferring pseudotime as a proxy for cell state using Palantir

To model cell state progression, we performed pseudotime analysis using Palantir^52^ on 1,176,697 cells aggregated across all pools and donors (N = 942) within the CD4 lineage (CD4_Naive_, CD4_TCM_, CD4_TEM_, CD4_Proliferating_, CD4_CTL_, and Treg). Briefly, Palantir infers pseudotime by projecting cells into diffusion space, constructing a neighbourhood graph, and estimating relative progression from a user-defined starting population, while also modelling lineage probabilities toward terminal states through a Markov chain framework. We first performed PCA using 75 PCs, followed by Harmony^85^ batch correction using the sequencing pool as the batch variable. A UMAP embedding was then generated from the batch-corrected 75 PCs using a 30-nearest-neighbour graph. Next, we used these embeddings to compute 100 diffusion components, before running Palantir with knn = 50 and num_waypoints = 8000.

### Testing the cell state-dependent caQTL effect using SAIGE-Dynamic

Based on cell-specific pseudotime estimates from Palantir, we applied SAIGE-Dynamic^34^, an extension of SAIGE-QTL designed to efficiently model heterogeneous genetic effects across varying cellular contexts directly from single-cell data, without the need for pseudobulk aggregation. First, a null Poisson mixed model was fitted for each gene across all cells. The inferred pseudotime was included as the dynamic (environmental) covariate, while sex, age, genotype PCs, and chromatin accessibility PCs were incorporated as static covariates. Next, association tests were performed for each genetic marker with minor allele frequency > 0.01 and minor allele count > 0.05, which were set to process 10,000 markers per chunk. Finally, the ACAT test^86^ was used to aggregate marker-level results into gene-level p-values.

For visualization of interaction patterns, we stratified the cell population into three pseudotime bins (early, mid, and late), which correspond to pseudotime ranges of 0-0.33, 0.33-0.66, and 0.66-1, respectively.

### Quantification of cell functions score in CD4+ cells

We computed cell function scores in CD4+ cells as cell states using scDeepID, a flexible multi-task transformer that leverages biological pathway databases as prior knowledge to train the model. First, we trained a scDeepID model with 2,390,948 CD4+ cells (including CD4_NAIVE_, CD4_TCM_, CD4_TEM_, CD4_TCM_, CD4_CTL_, T_REG_, CD4_PROLIFERATING_ cells) from TenK10K phase 1 single-cell RNA sequencing data. The training used expression features of 4,000 highly variable genes selected with Scanpy function highly_variable_genes (n_top_genes=4000m min_mean=0.0123, max_mean=3, min_disp=0.5) with human Reactome database as a guide, filtered to the top 200 most informative pathways based on the presence of pathway genes in the highly variable gene list. We then extracted the attention-based latent space for UMAP representation, and extracted the cell function scores followed by transformation of each cell function score to approximate a normal distribution with a zero mean and one unit variance.

### Gene regulatory network inference using multiome data

#### Infer cell type-specific GRN with GLUE

We performed gene regulatory network inference using GLUE^18^ in a cell type-specific manner. In specific, we aggregated cells from each major cell type across both scRNA-seq and scATAC-seq datasets and subsequently generated integrated, cell type-specific objects. Following the recommendations of the GLUE framework, we adopted a standard Scanpy-based pipeline for preprocessing. For scRNA-seq data, we normalized each cell by total counts over all genes, log-transformed normalized counts, and selected 6,000 highly variable genes per major cell type. Expression for each gene was clipped by max_value = 10 and then scaled to unit variance across cells. We further applied principal component analysis (n_pcs = 100) followed by Harmony with Scanpy to account for the technical batch effects between sequencing libraries. For scATAC-seq, we adopted the latent semantic indexing method implemented in GLUE for dimensionality reduction. The derived principal components were used to construct a nearest neighbor graph and UMAP using Scanpy with default parameters.

Next, we constructed the guidance graph required by GLUE by incorporating both genomic proximity and prior regulatory knowledge derived from eQTL data. For genomic distance, an ATAC peak was connected to a gene with a positive edge in the graph if it lies within a 150 kb window surrounding the gene’s promoter region. To further incorporate regulatory evidence from eQTL data, an additional edge was added for each peak-gene pair where the peak overlaps with a genomic locus associated with gene expression (eQTL). To ensure numerical stability, self-loops were incorporated into the graph. We then applied GLUE to integrate the multi-omics data using the constructed guidance graph to learn low-dimensional cell embedding for each omics layer. The *cis-*regulatory inference was subsequently performed based on these embeddings. The resulting GLUE regulatory score was computed as the cosine similarity between gene and peak embeddings, with higher scores indicating stronger putative regulatory interactions between peak-to-gene pairs. The peak-to-gene pairs were annotated as “with support” if at least one putative regulatory signal (such as an eQTL) colocalized with the pair. From the inferred peak-gene regulatory interactions, we utilized SCENIC^55^ to build the gene regulatory network for each major cell type.

To incorporate caQTL results for enhancing GRN inference, we exploited the causal inference (SMR) results linking eQTL and caQTL (restricting to caQTL-to-eQTL associations with *p*_SMR_ < 5 × 10^-8^) to identify high-confidence peak-gene pairs with strong regulatory potential. These interactions were used to enrich the *cis*-regulatory relationships inferred by the GLUE model, which then served as input for SCENIC-based GRN construction. By integrating these curated regulatory links, the TF-target gene relationships that might otherwise be missed can be identified.

### Compare inferred cis-regulatory scores between paired and unpaired multiome data

To evaluate the benefits of incorporating eQTL data and to compare differences in inferred *cis*-regulatory scores across datasets, we used the 10k Human PBMC dataset processed with Cell Ranger ARC 2.0.0 from 10X Genomics. In parallel, we generated a randomly sampled subset of the TOB multiome cohort with a comparable size (10,000 cells) to the 10X dataset. Both datasets underwent the same preprocessing procedure, as described in the previous section. The GTEx and cell type-specific eQTLs were incorporated into the GLUE model training.

### Comparison of QTL- and multiome-derived peak–gene links

To compare peak–gene links derived from QTL colocalization with those inferred from multiome data, we performed complementary analyses in CD14 monocytes. caQTL–eQTL colocalization results (PP.H4) were compared with peak–gene links inferred from multiome data using GLUE. Concordance between the two approaches was assessed using Spearman correlation, and further evaluated using a binned analysis of GLUE scores across increasing levels of colocalization evidence. In addition, analyses were repeated after restricting to peak–gene pairs with significant colocalization evidence (PP.H4 > 0.8).

Conceptually, the QTL-based framework evaluates whether a shared genetic variant influences both chromatin accessibility and gene expression, whereas GLUE infers peak–gene links from co-variation patterns across cells. These approaches capture distinct aspects of regulatory relationships and are therefore expected to yield partially overlapping but non-identical sets of peak–gene links. As a technical constraint, GLUE analysis was limited to peak–gene pairs within 150 kb and to highly variable genes.

### Benchmarking peak-to-gene link methods and GRN inference methods

To better understand the performance of GLUE framework in peak-to-gene links and GRN inference, we conducted two sets of benchmarking with existing tools: First, we benchmarked the peak-to-gene links from GLUE^18^ and pgBoost^57^, on three validation sets (GWAS, eQTL, and CRISPR). pgBoost is a recent method that infers peak-to-gene regulatory strengths, which has demonstrated superior performance compared to other approaches. We designated pgBoost as a state-of-the-art reference to evaluate whether GLUE can achieve comparable results. Using the PBMC 10k Multiome dataset from 10x Genomics, we extracted cells from four major cell types (CD14_Mono,_ CD4_TCM_, CD4_Naive_, CD8_Naive_) which encompass more than 1,000 cells. Both GLUE (with and without TenK10K eQTL prior guidance) and pgBoost were applied separately to the four cell types. Validation followed the same sets used in the pgBoost study, and method performance was assessed by evaluating whether predicted peak-to-gene pairs from each method scored higher for pairs present in the validation sets.

Second, we benchmarked the GRN inference results against CellOracle^87^ and SCENIC+^56^, two state-of-the-art methods that can integrate single-cell multiome data to infer gene regulatory networks. We applied GLUE (with and without incorporating TenK10K eQTL and caQTL for enhancement) and the two comparison methods to the 10x Genomics PBMC dataset, deriving a list of TF-target gene pairs for each approach. For quantitative evaluation, we curated three validation sets from different types of evidence: gene expression changes following TF knockout, ChIP-seq data, and functional protein-protein interactions. Following established benchmarking practice^88^, we used Early Precision Ratio (EPR) as the evaluation metric, which measures the ratio of the fraction of true positives among predicted edges to that expected for a random predictor.

### Drug enrichment analysis

To assess whether single-cell caQTL data can support the identification of drug targets, we performed an enrichment analysis of target–indication pairs using the Open Targets Platform^58^ (OTP, version 25.12), following an approach analogous to (Henry et al. medRxiv. 2025). We first used peak-to-trait SMR results to shortlist peaks with putative causal effects on selected disease traits, then integrated these with peak-to-gene SMR results to nominate candidate effector genes for each trait. Effector gene–trait pairs were subsequently intersected with target–indication pairs from OTP, and the highest clinical development phase achieved for drugs whose mechanism of action matched the relevant target–indication pair was extracted. Pairs were stratified by maximum clinical phase reached (Phase I, II, III, Phase I+, Phase II+, Phase III+, and Approved), and for each stratum a 2×2 contingency table was constructed by enumerating target–indication pairs that were or were not supported by our caQTL-derived effector genes.

We note two important limitations of this analysis. First, the number of target–indication pairs supported by both caQTL evidence and clinical validation (true positives) is small, and estimated odds ratios should be interpreted with caution. Second, and more fundamentally, the enrichment framework assumes that the absence of a caQTL- or eQTL- derived effector gene nomination for a given target–indication pair constitutes a negative result. However, this reflects incomplete data coverage rather than a true biological negative. Many nominally unsupported pairs may yet prove relevant as QTL mapping power improves or as additional traits are integrated. The enrichment results should therefore be interpreted as a lower-bound estimate of the concordance between QTL-nominated targets and established drug mechanisms.

## Data Availability

Individual-level data (raw sequencing outputs and processed count matrices from scATAC-seq and multiome) generated in this study are available through EGA (upload in progress). WGS and scRNA-seq data generated by the TenK10K flagship project and used in this study are available through EGA and the study IDs are EGAS50000001653 (TenK10K Phase 1: Single Cell data), EGAS50000001654 (Tenk10k Phase 1: Whole Genome Sequencing data).
All key summary results, including peak annotations, caQTL summary statistics (common and rare variants), fine-mapping results (SuSiE and mvSuSiE), ChromBPNet model weights and predicted scores, cell state-dependent caQTL summary statistics, colocalization results (coloc and SMR), and GLUE peak-gene and TF-gene links, are available on the Hugging Face platform (https://huggingface.co/datasets/anglixue/TenK10K_multiome).
Any additional information required to reanalyze the data reported in this paper is available from the corresponding authors upon request.

https://github.com/powellgenomicslab/tenk10k_phase1_multiome

## Acknowledgments

We acknowledge Dr. Kai Zhang at Westlake University for guidance in running SnapATAC2 software and Dr. Bing Ren providing the quality controlled scATAC-seq objects from Zhang et al. *Cell*. 2019. We acknowledge Dr. Zhijie Cao at Peaking University for guidance in running GLUE software. We acknowledge Dr. Tim Stuart at Genome Institute of Singapore for guidance in running Signac. We acknowledge Dr. Rahul Satija at New York Genome Center for guidance in running Seurat and using Azimuth reference. We acknowledge Dr. Michael Geaghan and Eric Urng from the Garvan Data Science Platform for support. investigator grant (2009982). The contents of this manuscript are solely the responsibility of the authors and do not reflect the views of the NHMRC. A.X. is supported by NHMRC Investigator grant 2033018. J.E.P. is supported by NHMRC Investigator grant 2034556, and a Fok Family Fellowship; D.G.M. is supported by an NHMRC investigator grant (2009982). G.A.F. and the BioHEART Study have been supported by NHMRC Investigator Grant, NSW Health Office of Health and Medical Research, and the NSW Health Statewide Biobank scheme.

## Author contribution

A.X. and J.E.P. designed the study. A.X. designed and performed the single-cell processing and caQTL mapping pipelines and led the writing of the manuscript. A.X. and J.F. designed and performed the causal inference and gene regulatory network inference analyses. A.X. and O.A.D. designed and performed the caQTL and colocalization analysis. A.X., J.F., and O.A.D designed and performed the replication analysis. A.X. and O.A.D. designed and performed the WASP analysis, with the help from B.B. H.L.H designed and performed the dimensional reduction and fine-mapping analyses. E.S., E.S.Z, and C.B. designed and performed the single-cell ATAC and multiome sequencing experiments. A.X. and B.B. processed the single-cell ATAC sequencing data. B.B. performed the bioinformatic 10X pipelines for reads alignment. A.S.E.C contributed the scRNA datasets and eQTL results. A.X. performed the quality controls and cell type annotation of the scATAC-seq data, with the help from J.F. and O.A.D. A.S.E.C, A.H. and J.E.P. designed and A.H. performed the cell type specificity analysis. A.X. and O.A.D. designed and performed the rare caQTL analysis, with the help from Z.W. H.A.T. first established the colocalization pipeline and conducted functional annotations. P.C.A., A.X., J.F., and O.A.D designed and performed the cell state-specific caQTL mapping. H.L.H. and A.X. designed and performed the cell function analysis with scDeepID. A.X. and J.F. designed and performed the gene regulatory network analysis. A.X., J.F., H.L.H., P.C.A., H.A.T., D.G.M., and J.E.P contributed to downstream interpretation. J.F, H.L.H, B.B. and Z.Q. provided support with the caQTL mapping pipeline. Z.Q. provided assistance in pseudotime estimation and trajectory inference. L.C., E.B.D., and K.K.H.F. designed and performed the annotation of variants using ChromBPNet. L.C. performed the identification of rare promoter/enhancer variants associated with outlier gene expression, with the input from A.X. E.D. provided curated validation datasets and conducted benchmarking analysis for pgBoost and GLUE, with the help from J.F. and A.X. J.F. performed the benchmarking analysis for gene regulatory network inference methods with the inputs from A.X. A.H. performed the drug enrichment analysis. L.H. improved the SAIGE-QTL software and computational efficiency.

Y.C.L. developed the SAIGE-QTL Dynamic function for interaction analysis. D.R.N. assisted with the scATAC-seq sample demultiplexing. A.S.L. contributed a curated list of regulatory annotations used to annotate the results and H.A.T. performed the annotations for caQTLs. A.S. contributed to the quality control of genotype array data. R.A.M. and V.C. designed the LBIO study. W.Z. helped with the implementation of SAIGE-QTL, design of rare variant mapping, and interpretation of the results. A.L.P. provided curated validation datasets and constructive guidance on benchmarking design and interpreting the results. K.M.dL., D.E.G, and J.P. led the processing of whole genome sequencing data. G.A.F. designed the BioHEART study. A.W.H. designed the Tasmanian Ophthalmic Biobank study.

All authors contributed to editing the manuscript. A.X and J.E.P conceived the project and supervised all aspects of the work.

## Data and materials availability

Individual-level data (raw sequencing outputs and processed count matrices from scATAC-seq and multiome) generated in this study are available through EGA (upload in progress). WGS and scRNA-seq data generated by the TenK10K flagship project and used in this study are available through EGA and the study IDs are EGAS50000001653 (TenK10K Phase 1: Single Cell data), EGAS50000001654 (Tenk10k Phase 1: Whole Genome Sequencing data).

All key summary results, including peak annotations, caQTL summary statistics (common and rare variants), fine-mapping results (SuSiE and mvSuSiE), ChromBPNet model weights and predicted scores, cell state-dependent caQTL summary statistics, colocalization results (coloc and SMR), and GLUE peak–gene and TF–gene links, are available on the Hugging Face platform (https://huggingface.co/datasets/anglixue/TenK10K_multiome).

Any additional information required to reanalyze the data reported in this paper is available from the corresponding authors upon request.

## Code availability

All the analysis code is available at https://github.com/powellgenomicslab/tenk10k_phase1_multiome

## URLs

scDeepID: https://github.com/powellgenomicslab/scDeepID_TenK10K_manuscripts

Azimuth reference: https://azimuth.hubmapconsortium.org/references/#Human%20-%20PBMC

