## Supplementary Figures for "Genetic regulation of cell type–specific chromatin accessibility shapes immune function and disease risk"

Xue et al. 2025

**Supplementary Information**

**Content**

Supplementary Notes
Supplementary Figures

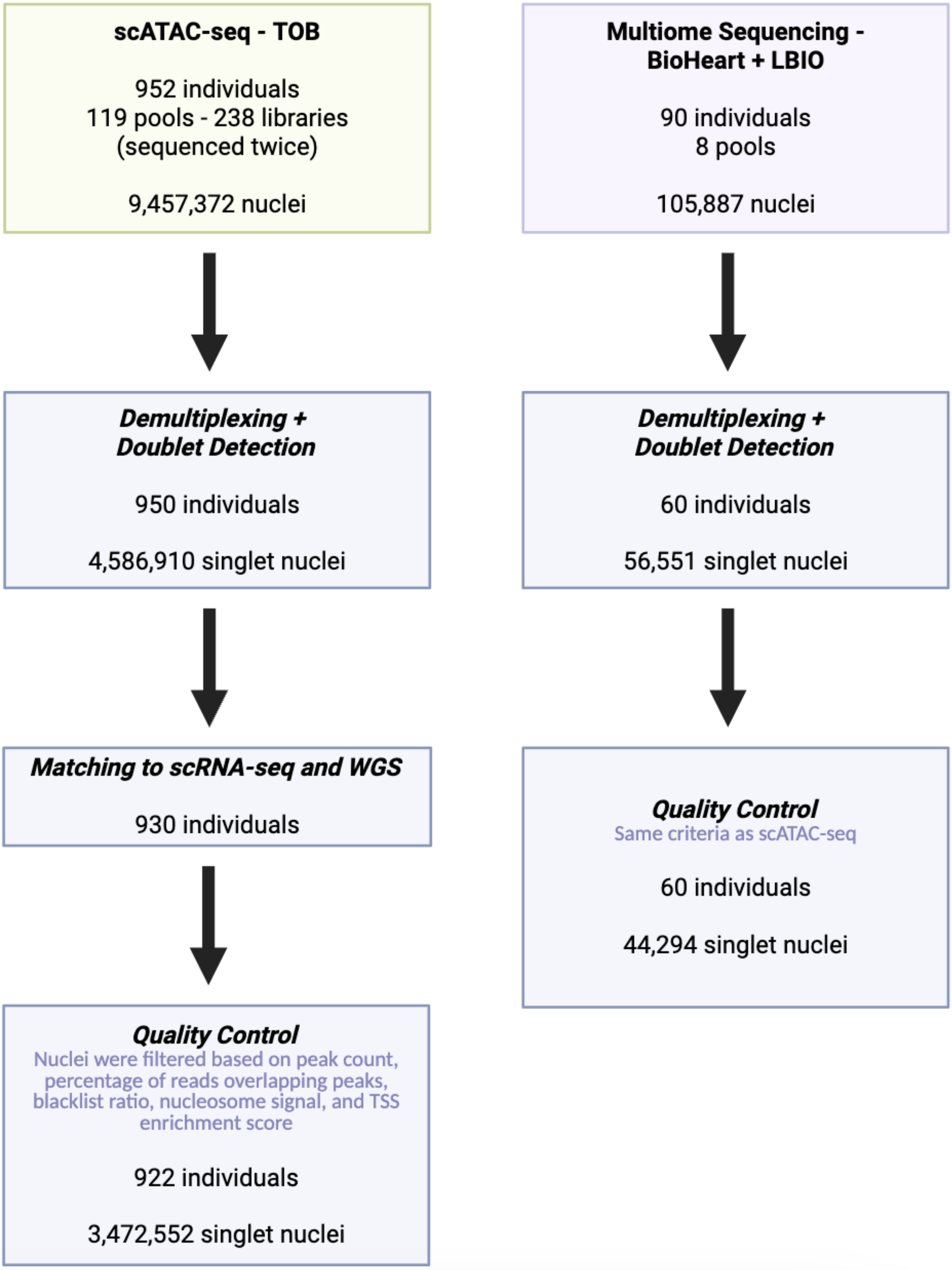


**Supplementary Figure 1. Workflow of sample collection and quality control for the scATAC-seq and multiome datasets**. The number of donors and nuclei for each step was described.

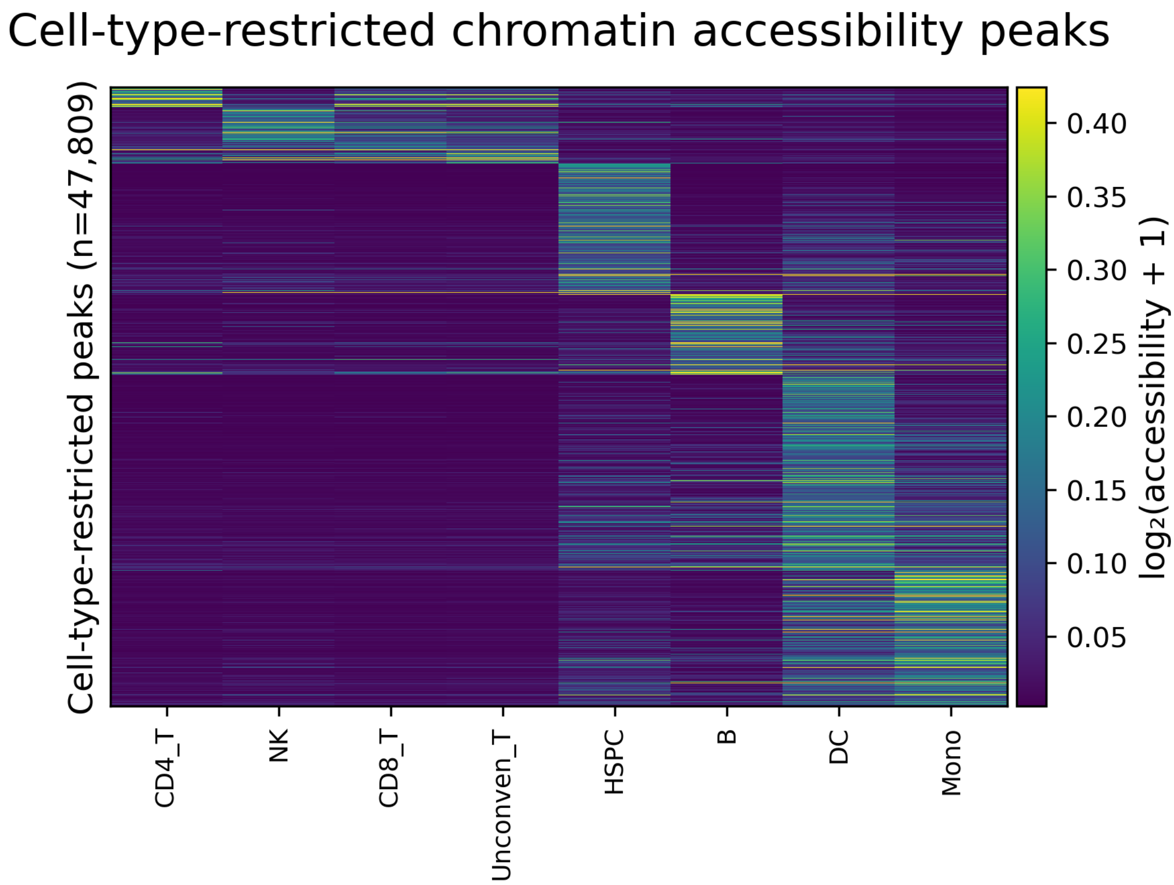


**Supplementary Figure 2. Heatmap of cell type-specific marker peaks across 8 main cell type categories**. Color scale reflects log2(RPKM+1) chromatin accessibility. The x-axis represents each of the 8 major cell type groups. The y-axis represents the 47,809 cell type-restricted peaks inferred from the Shannon entropy-based method (Schug et al., 2005) as implemented in Zhang et al. (Cell, 2019).

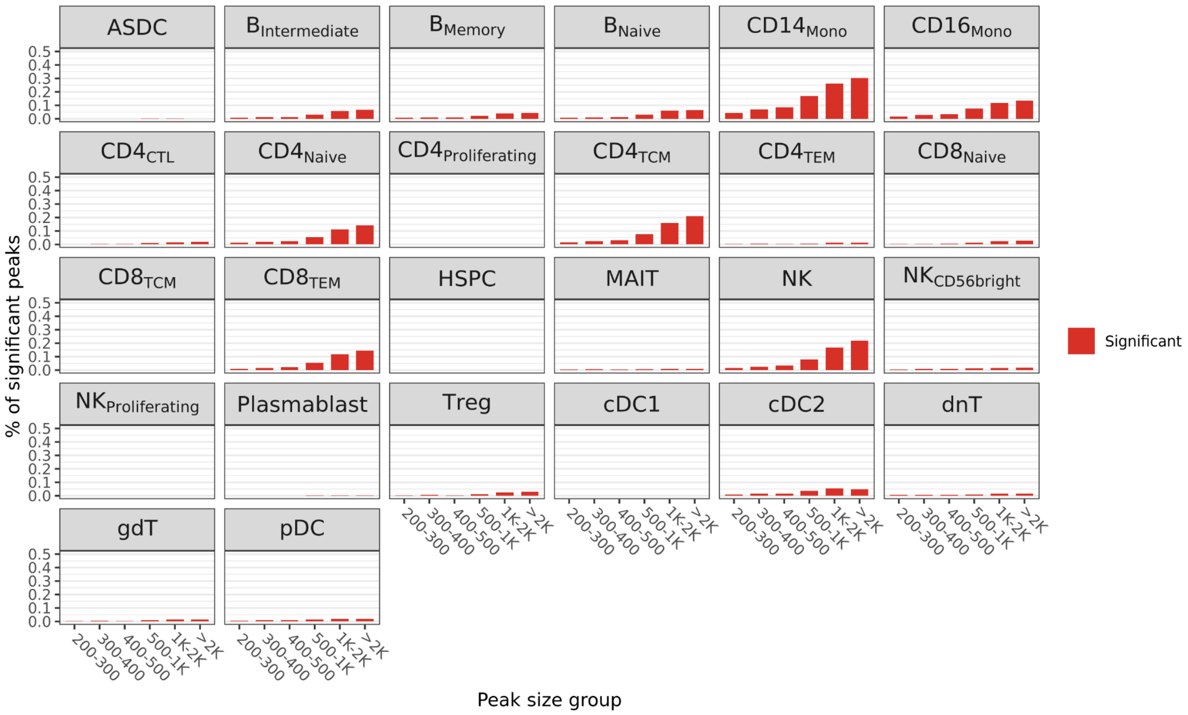


**Supplementary Figure 3. Proportion of significant peaks in different width groups across 26 cell types**. The x-axis indicates the group of peak width in base pairs. Peaks are grouped into six size bins (200–300 bp, 300–400 bp, 400–500 bp, 500 bp–1 kb, 1–2 kb, and >2 kb). The y-axis indicates the proportion of significant peaks in each group. Each panel represents a cell type.

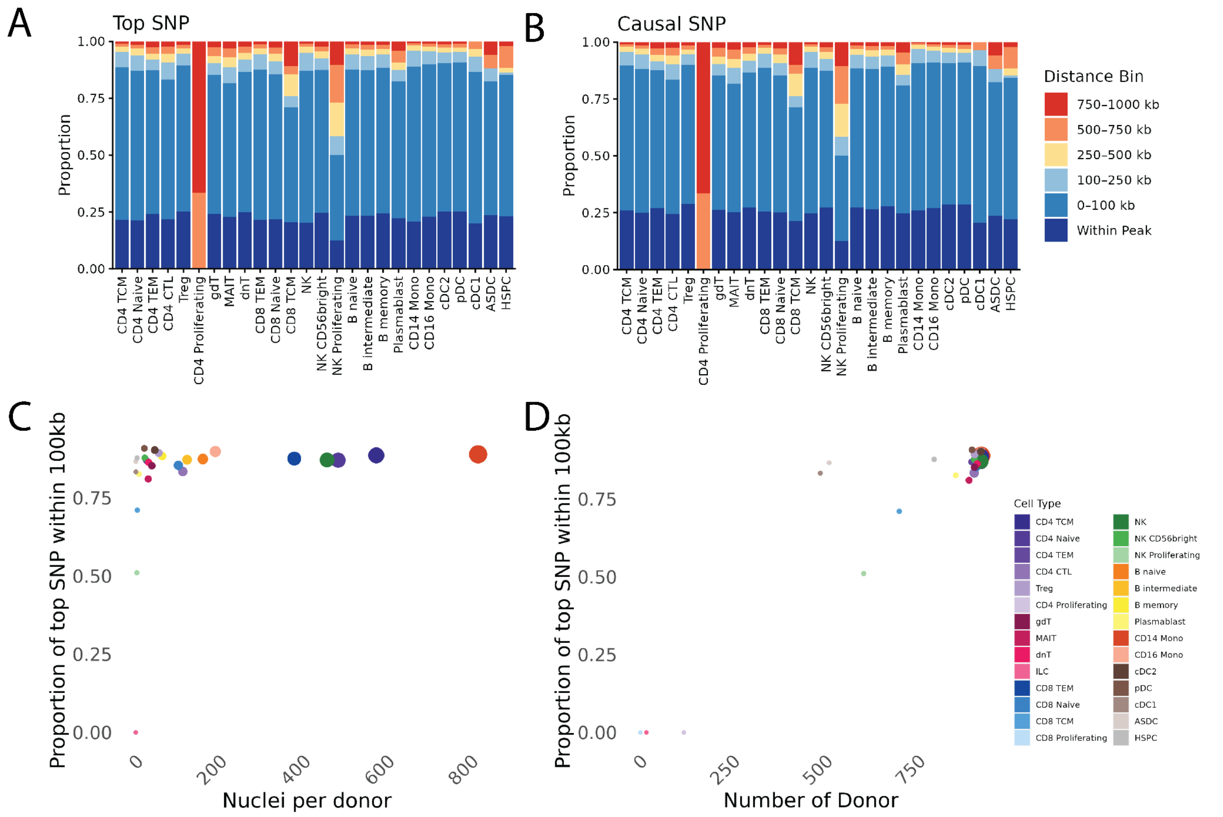


**Supplementary Figure 4. Distance between the top caQTL variant and peak center across different cell types.** (**A**) Stacked bar plot showing the distribution of distances between top caQTL variants and the centre of their associated peaks. (**B**) Stacked bar plot showing the distribution of distances between fine-mapped variants in credible sets and the centre of their associated peaks. (**C**) Proportion of top SNPs within 100 kb of peak centres plotted against the number of nuclei per donor for each cell type. (**D**) Proportion of top SNPs within 100 kb of peak centres plotted against the number of donors per cell type.

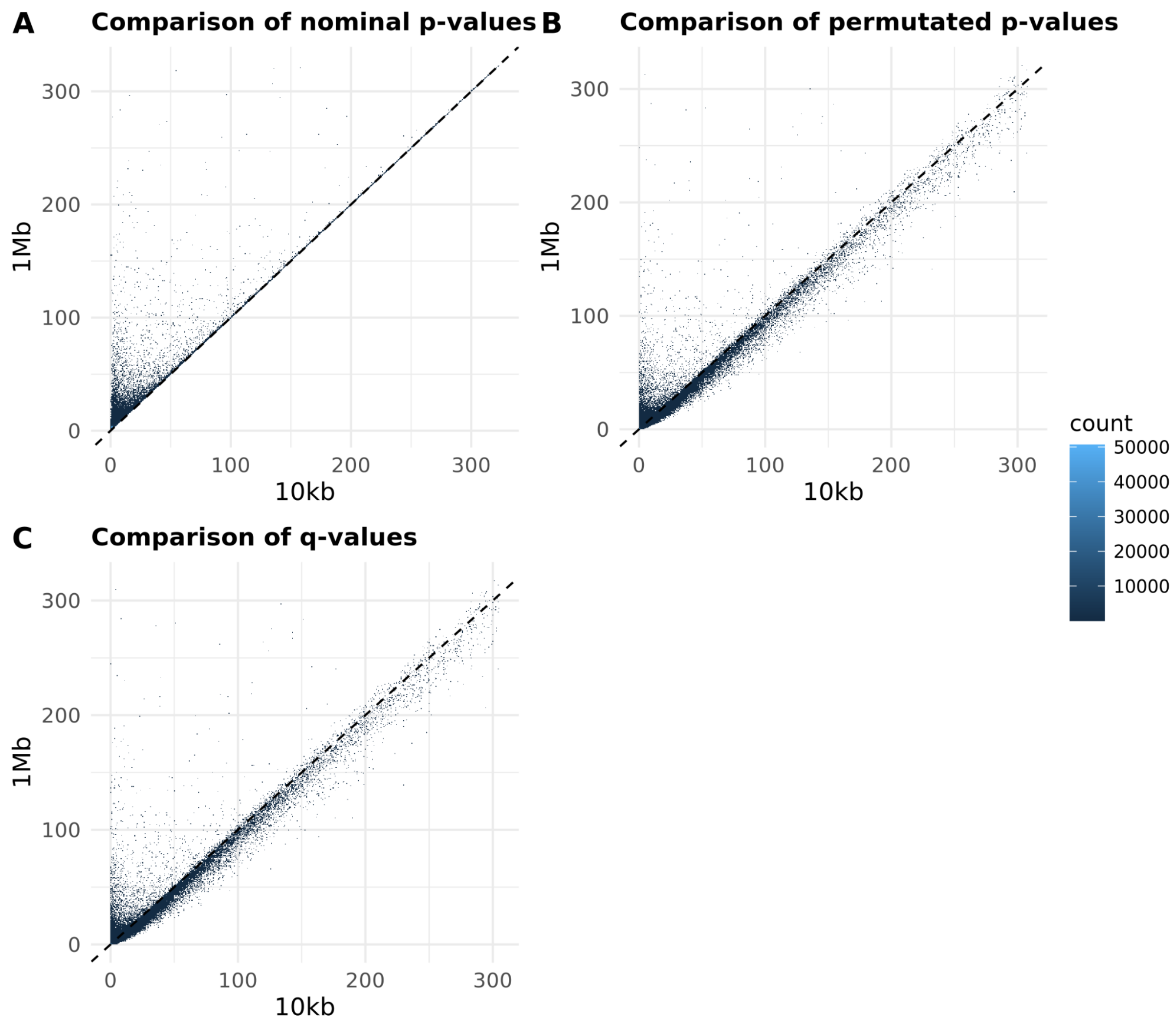


**Supplementary Figure 5. Comparison of significant levels of top variants for all peaks tested for CD14$_{\text{Mono}}$ between 10kb and 1Mb run**. Three metrics are compared in each panel, including (**A**) Nominal p-value. (**B**) Beta-approximation corrected p-value. (**C**) q-value. The scale of both axes is based on -log10. Dots are merged as bins, and the color indicates the density of each bin. Dots are merged into bins and the color indicates the size of each bin.

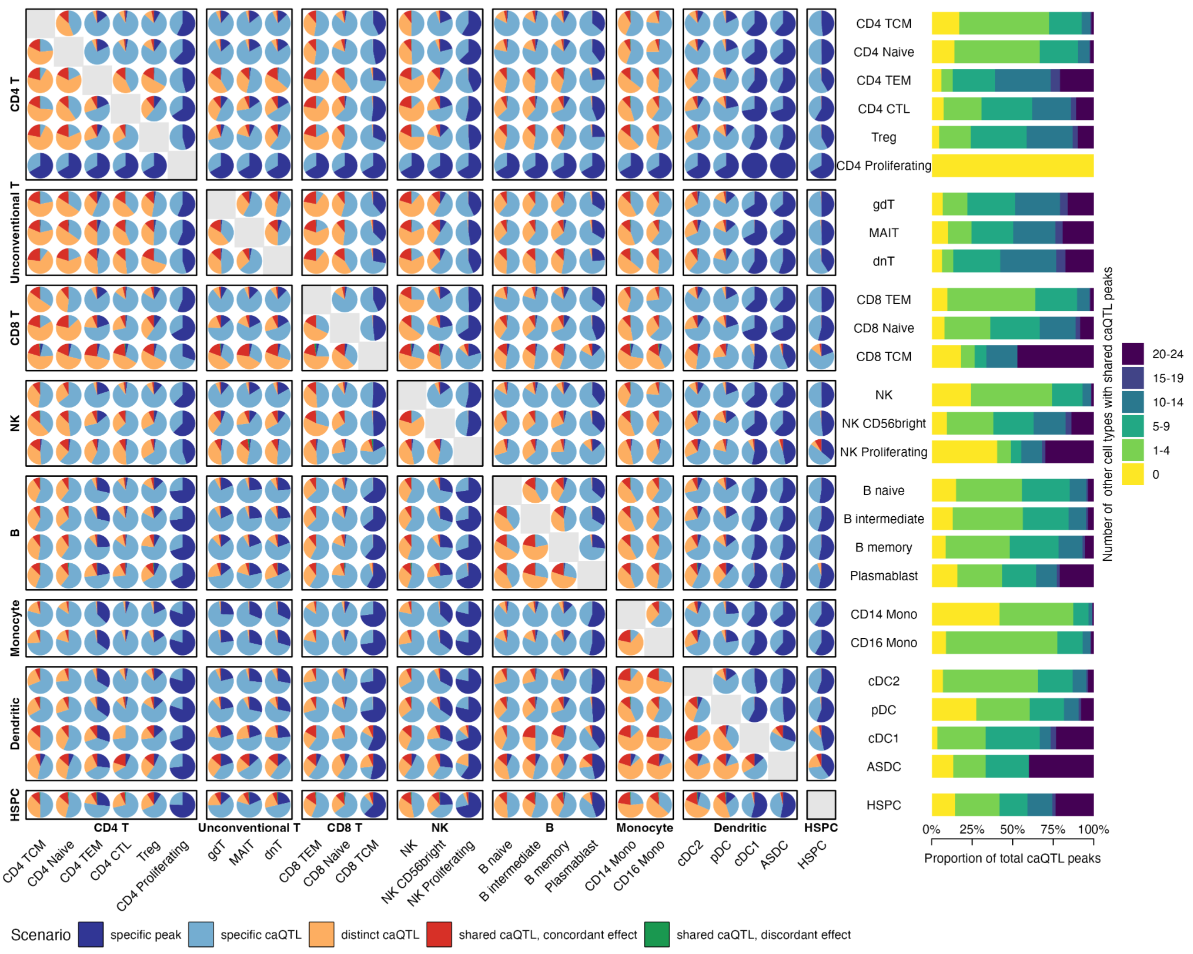


**Supplementary Figure 6. Quantifying the cell-type-specificity of caQTL signals**. Cell-type specificity of caQTLs, categorized into five different scenarios (pie chart). The upper triangle indicates the scenario of the caQTL from the cell type on the left side, corresponding to the cell type along the bottom. Bar plot (right) shows the distribution of caPeaks by the number of cell types in which the effects are shared (scenarios 3-5).

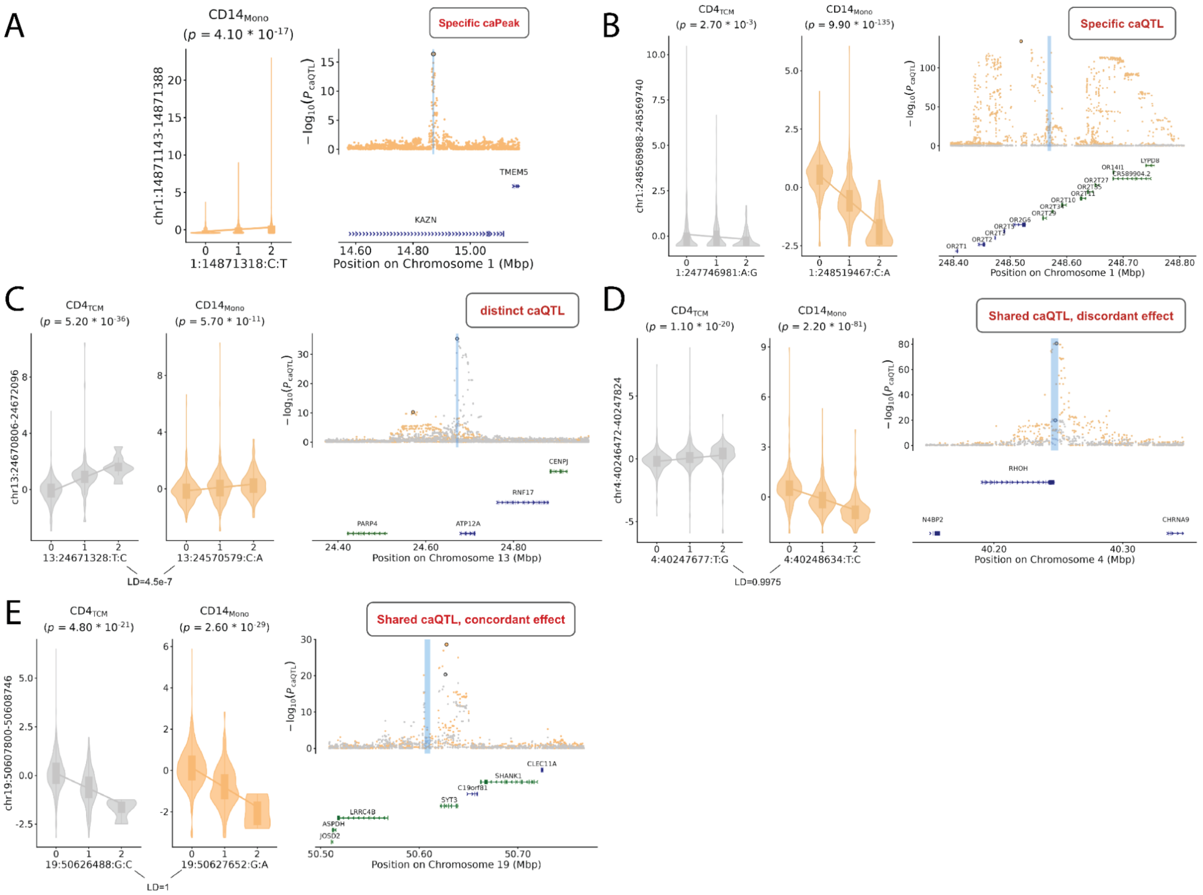


**Supplementary Figure 7. Schematic locus plot for cell-type specificity of caQTLs into five different scenarios**. This combined plot demonstrates the five cell type-specific caQTL scenarios. Each panel contains the caQTL boxplot in $CD4_{TCM}$ and $CD14_{Mono}$ and one locus plot showing the caQTL results in two cell types. The boxplot and scatter in grey indicates the $CD4_{TCM}$ results and orange indicates the $CD14_{Mono}$ results. The vertical blue shade indicates the genomic interval of the peak tested. (**A**) Scenario 1 - specific caPeak: chr1:14871143-14871388. The peak is not open in $CD4_{TCM}$ so not association test was performed. (**B**) Scenario 2 - specific caQTL: chr1:248568988-248569740. (**C**) Scenario 3 - distinct caQTL: chr13:24670806-24672096. (**D**) Scenario 4 - shared caQTL, concordant effect: chr19:50607800-50608746. **E**) Scenario 5 - shared caQTL, discordant effect: chr4:40246472-40247824.

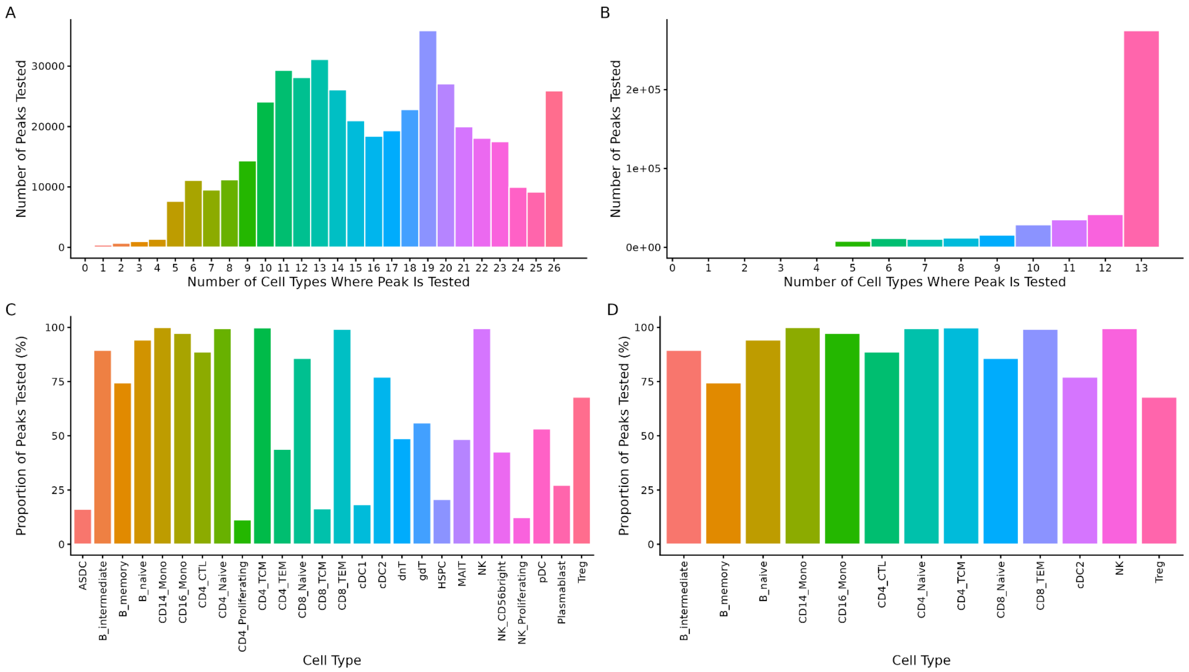


**Supplementary Figure 8. Restriction of the number of tested peaks in mashR**. (A) Distribution of peaks by the number of cell types with accessibility. Each bar represents the number of peaks accessible in n cell types. 25,892 out of 440,962 peaks were tested in all 26 cell types (5.87%). (B) Same as panel A, but only focusing on abundant cell types, which contain more than 40,000 nuclei. There are 13 abundant cell types in total. (C) Proportion of peaks tested in each cell type (D) Proportion of peaks tested in 13 abundant cell types.

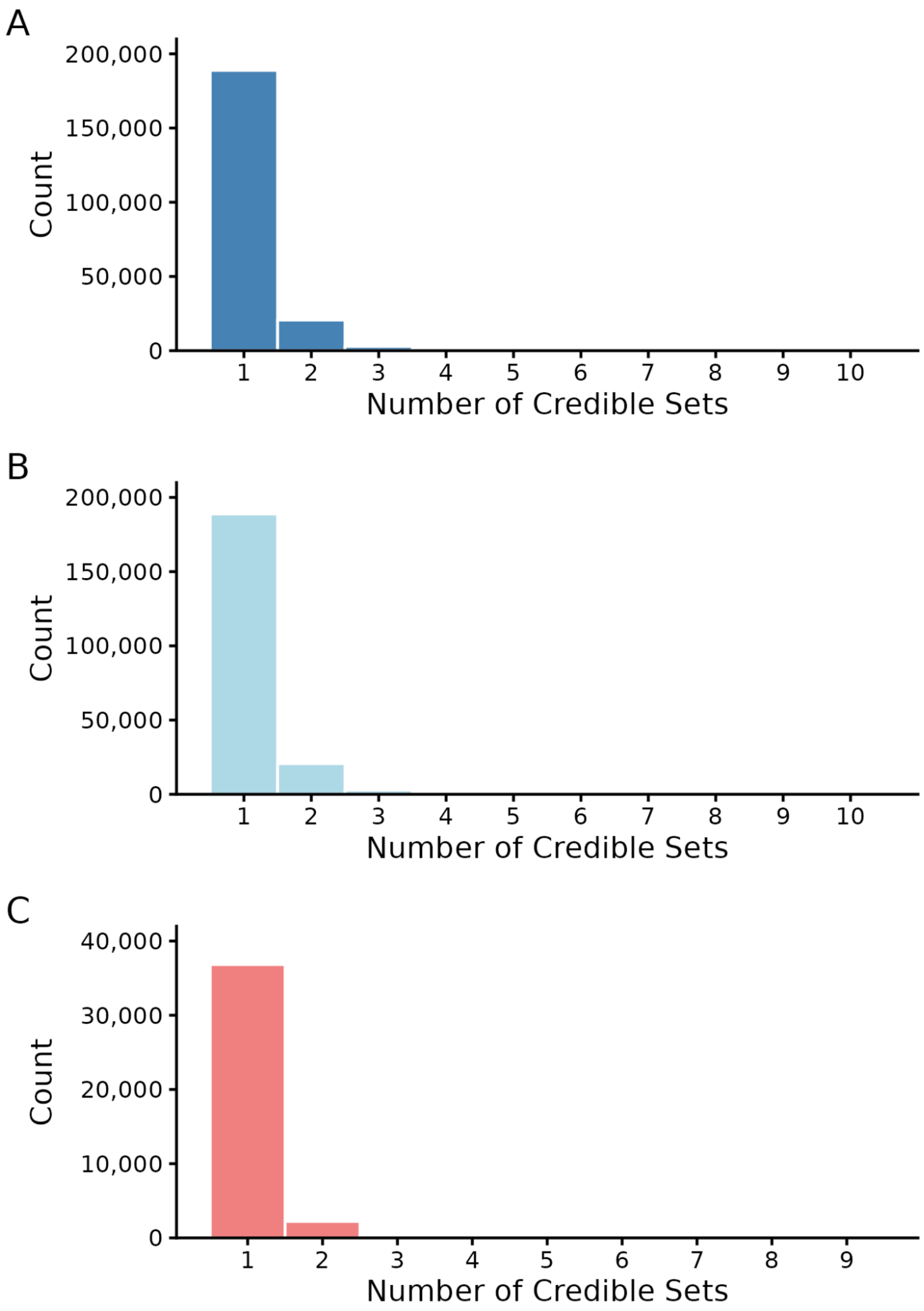


**Supplementary Figure 9. Number of credible sets for caQTL signals in each peak**. The histogram indicates the number caPeaks with different numbers of credible sets under different PIP thresholds (A) caPeaks with no PIP threshold; (B) caPeaks with the threshold of sum of PIPs > 0.9; (C) caPeaks with at least one variant with PIP > 0.9.

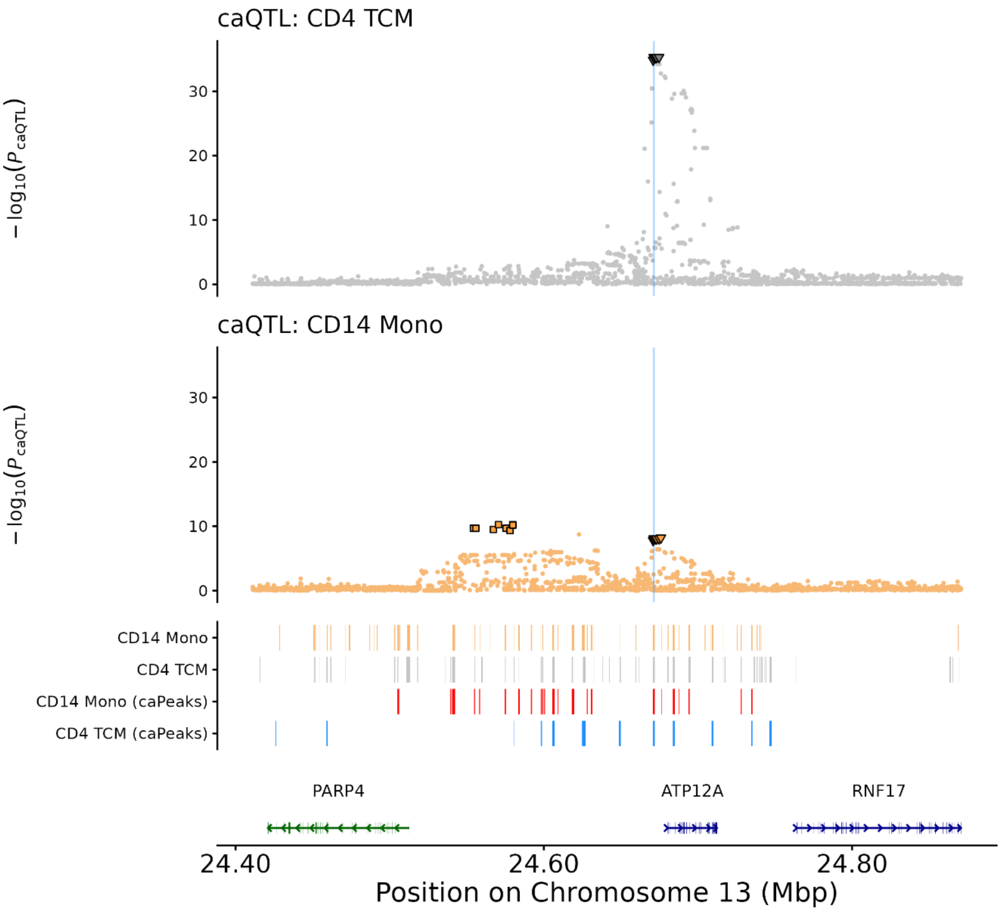


**Supplementary Figure 10. Fine-mapped credible sets explain the distinct caQTL signals between two cell types**. The caQTL locus plot of peak chr13:24670806-24672096 in $CD4_{TCM}$ and $CD14_{Mono}$, annotated by credible set(s). The inverted triangle represents the variants in credible set #1, which overlaps with the peak. The square represents the variants in credible set #2, which is located in the intergenic region upstream of *ATP12A*. The bottom track indicates the location of peaks and genes. For the peak track, the first two rows indicate the peaks tested and the next two rows indicates caPeaks in two cell types. For the gene track, the color indicates the strand direction.

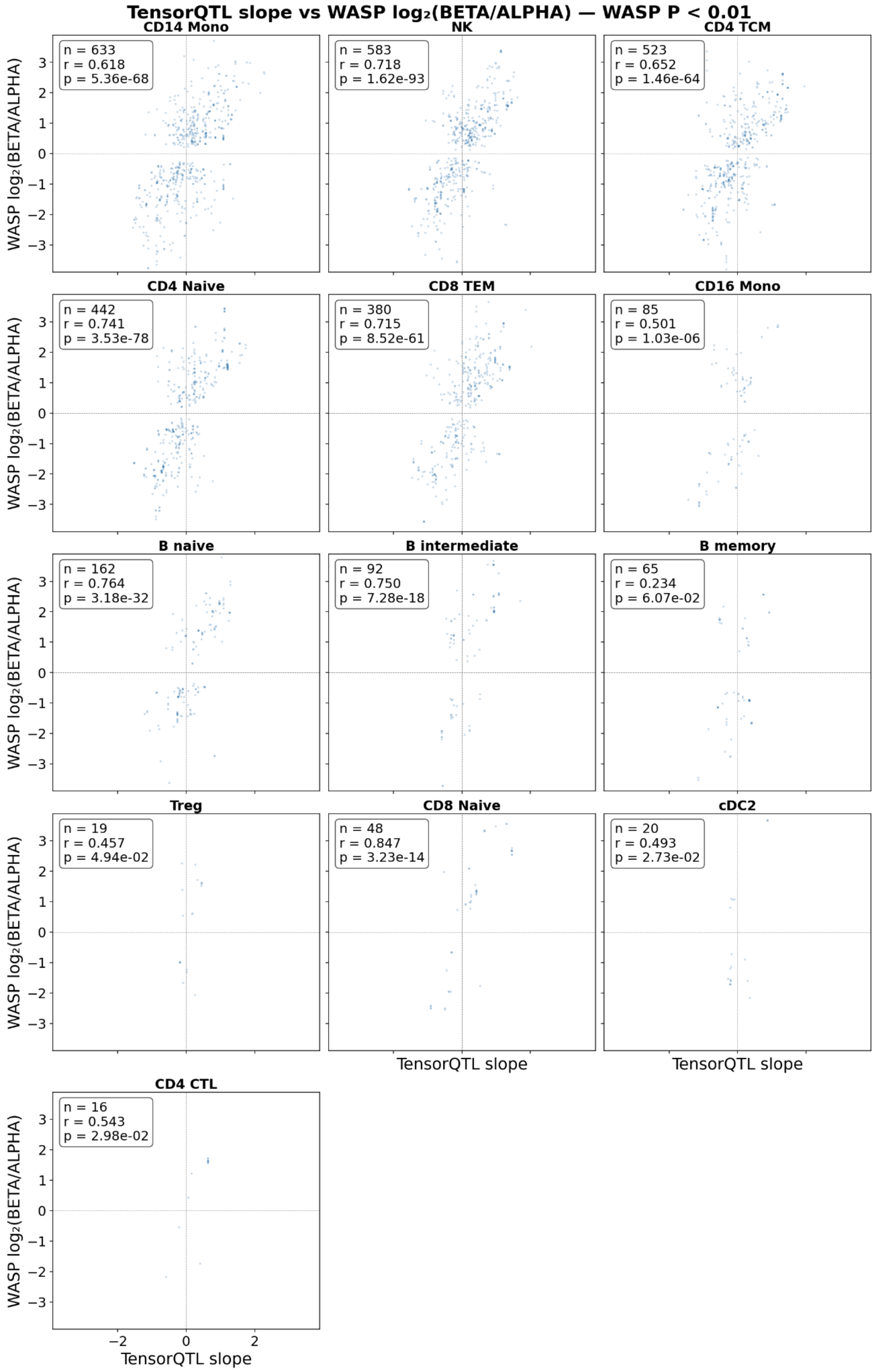


**Supplementary Figure 11. Comparison of allelic effect on chromatin accessibility between TensorQTL and WASP**. The slope of significant caQTLs from TensorQTL is compared to the WASP estimation of log2 of (expression level of alternative allele / expression level of reference allele). Each panel indicates a cell type with significant results in both analyses. In the top-left corner of each panel, it shows the number of SNP-peak pairs (n), Spearman's correlation between two methods (r), p-value of the correlation.

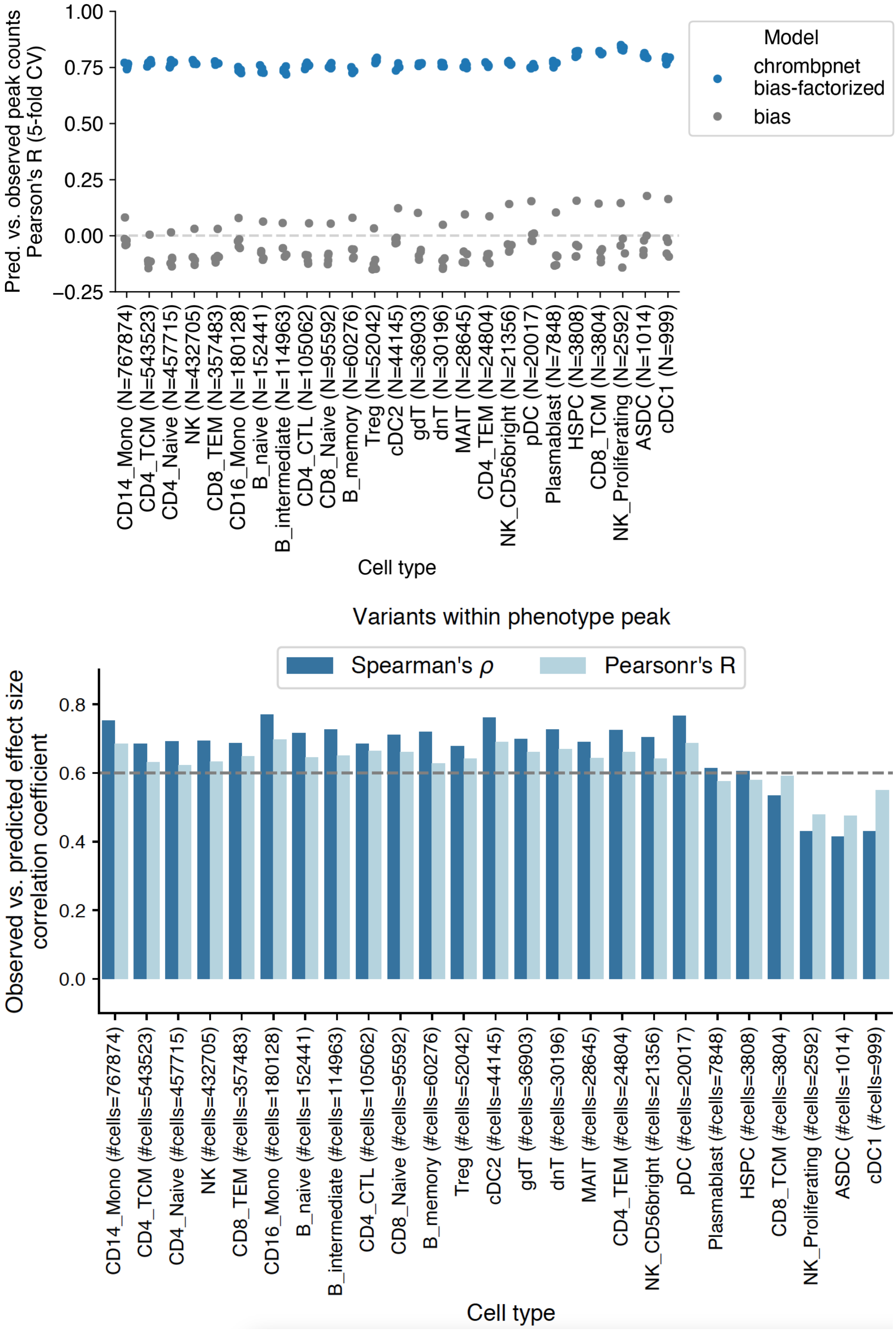


**Supplementary Figure 12. ChromBPNet variant scores correlate with observed caQTL effect sizes.** (**A**) ChromBPNet performance in 26 cell types. (**B**) The correlation between predicted and observed effect sizes stratified by cell types and the estimates are both calculated based on Spearman's and Pearson's correlation coefficient. The x-axis represents the 26 cell types and the number in the bracket indicates the number of peaks tested.

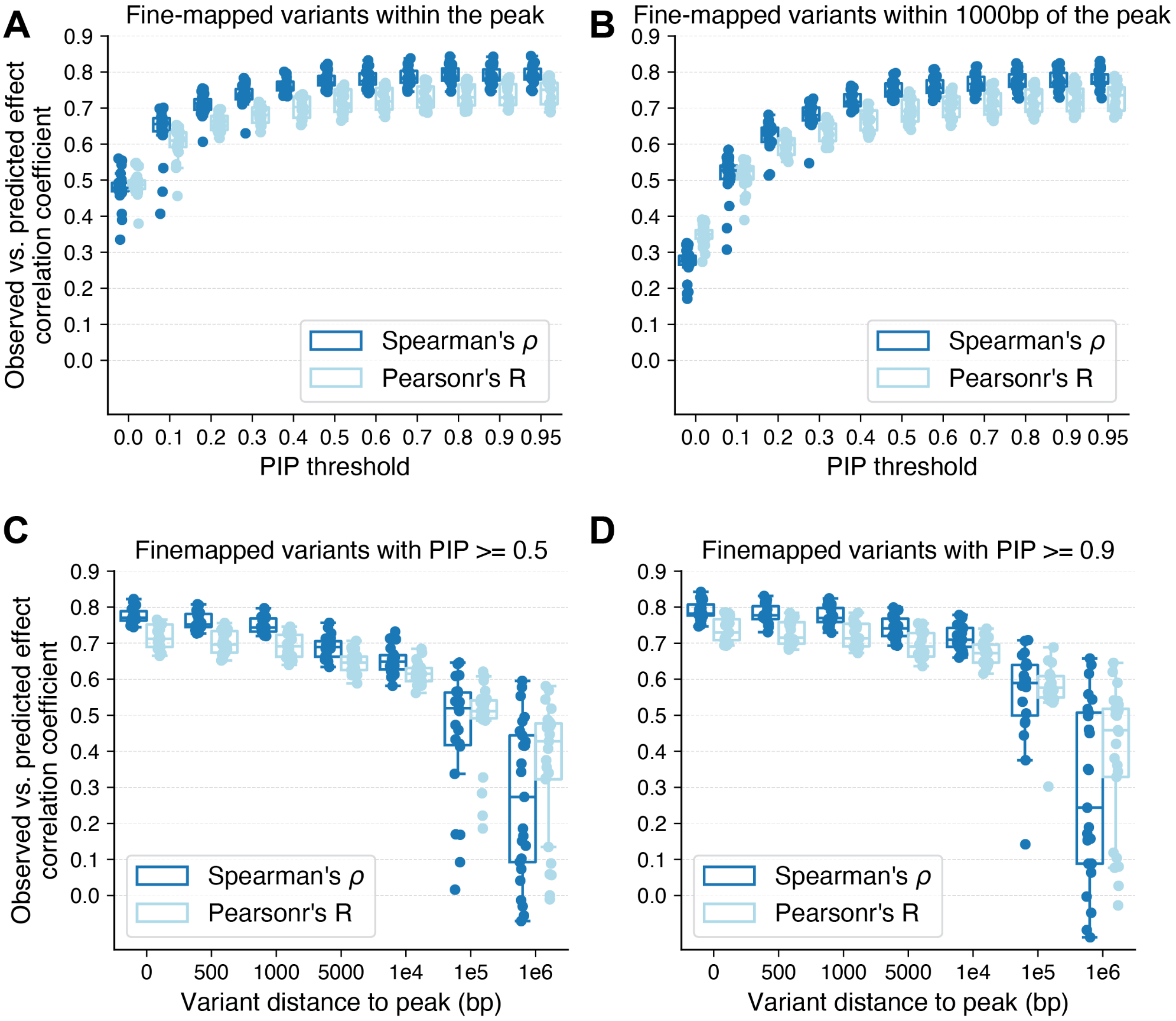


**Supplementary Figure 13. The correlation between predicted and observed effect sizes stratified by the variant-peak distance and fine-mapped PIP values.** (A-B) The x-axis indicates the group of fine-mapped variants stratified by the PIP threshold. The y-axis indicates the correlation coefficient. The fine-mapped variants within the peak are presented in panel A while those 500-1000 bp away are presented in panel B. (C-D) The x-axis indicates the group of fine-mapped variants stratified by the variant-peak distance. The y-axis indicates the estimates of the correlation coefficient. The fine-mapped variants with PIP >=0.5 are presented in panel C while those with PIP >=0.9 are presented in panel D.

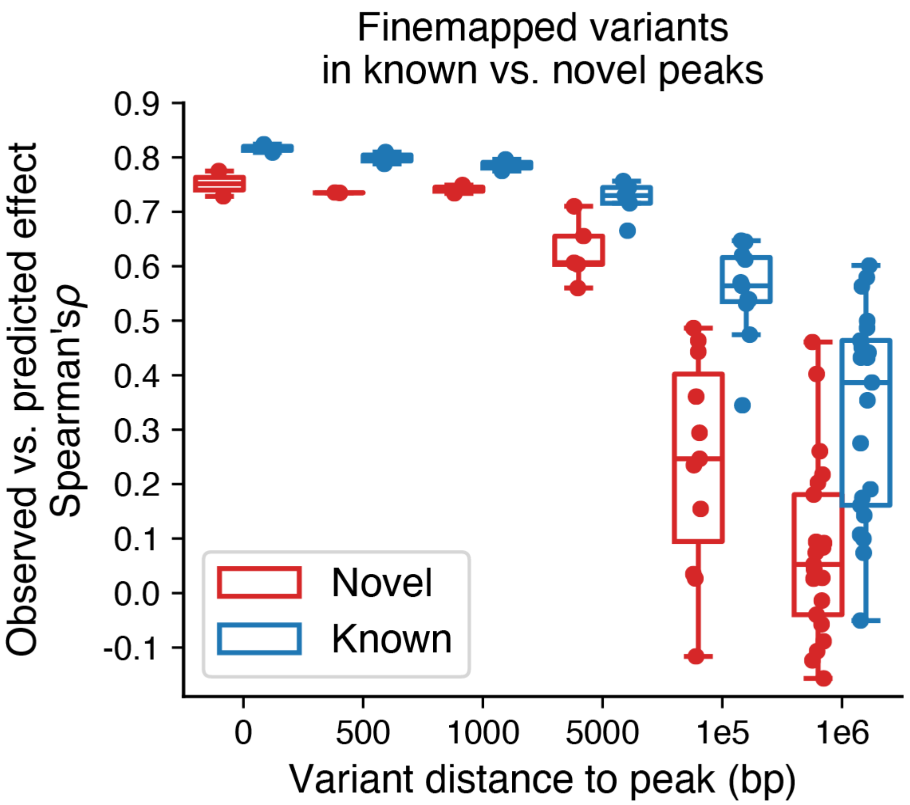


**Supplementary Figure 14**. **ChromBPNet prediction accuracy comparison between novel and known peaks.** The x-axis indicates the group of peaks stratified by variant-peak distance. The y-axis indicates the Spearman's correlation coefficient. Data points are grouped by novel and known peaks. There are only a few cell types with enough data points, so it is set to at least 20 variants to allow relatively robust estimation for correlation coefficients.

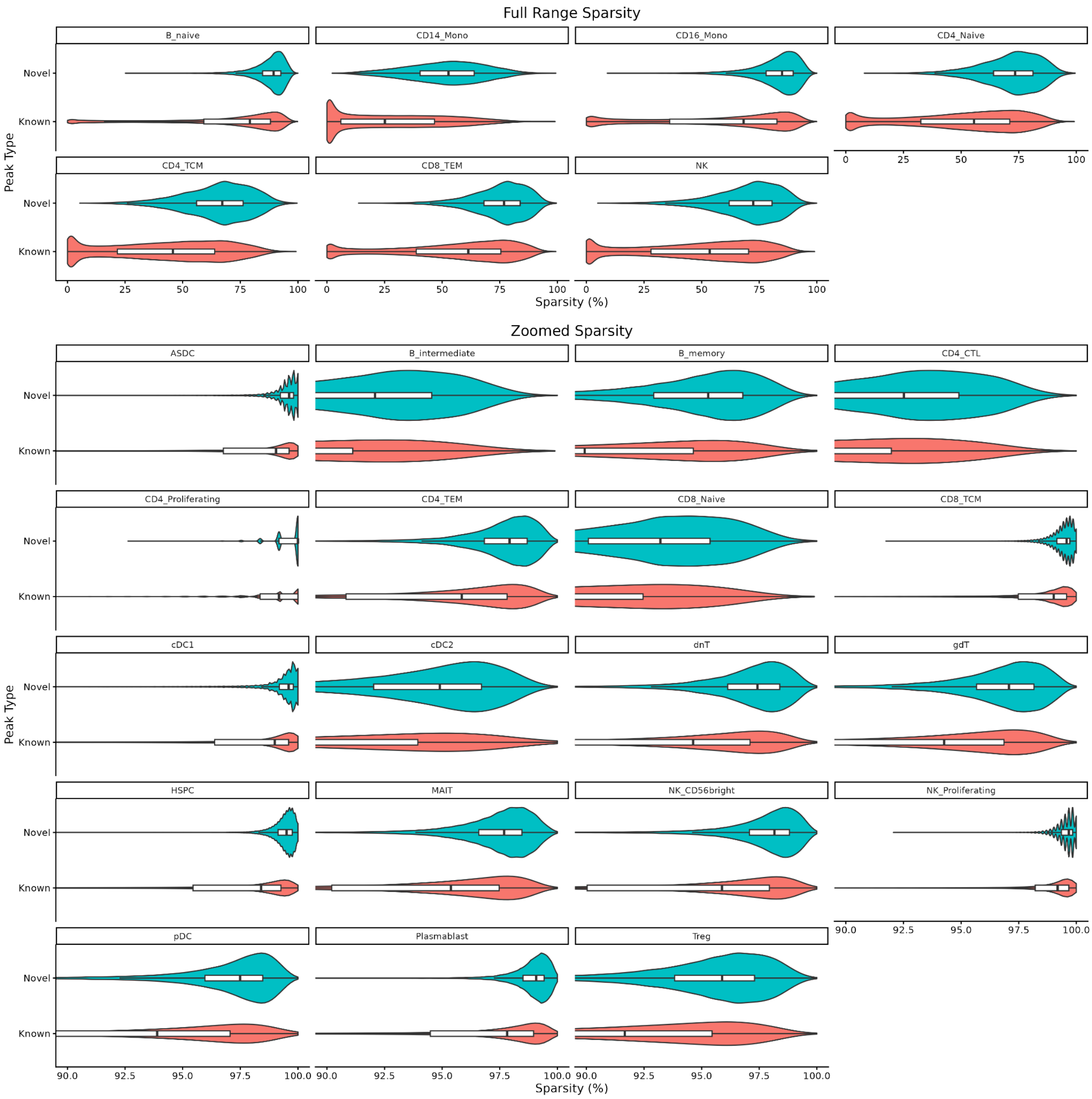


**Supplementary Figure 15**. **Donor-level sparsity for the novel and known peaks in each cell type.** The sparsity level on the x-axis is calculated based on the proportion of donors with zero accessible level for the peaks. The y-axis represents the novel and known peaks group. The upper layer displays the 7 cell types with relatively low sparsity level (≤ 90%) and the bottom layer displays the 16 cell types with high sparsity level (> 90%).

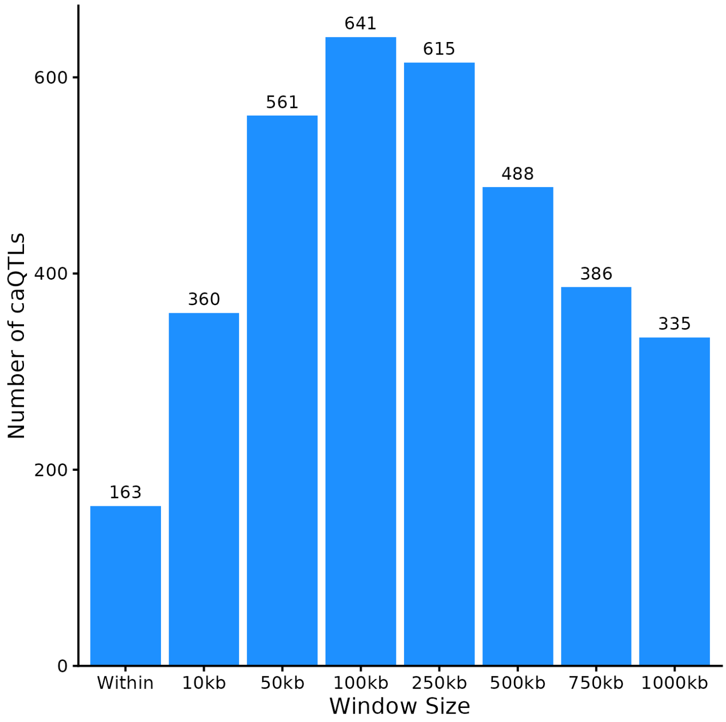


**Supplementary Figure 16. Sensitivity test for window size used for rare caQTL mapping with SAIGE-QTL**. The x-axis indicates different window sizes used for the rare caQTL mapping. The y-axis indicates the number of significant caQTLs. The test was done in CD4 Naive cell type.

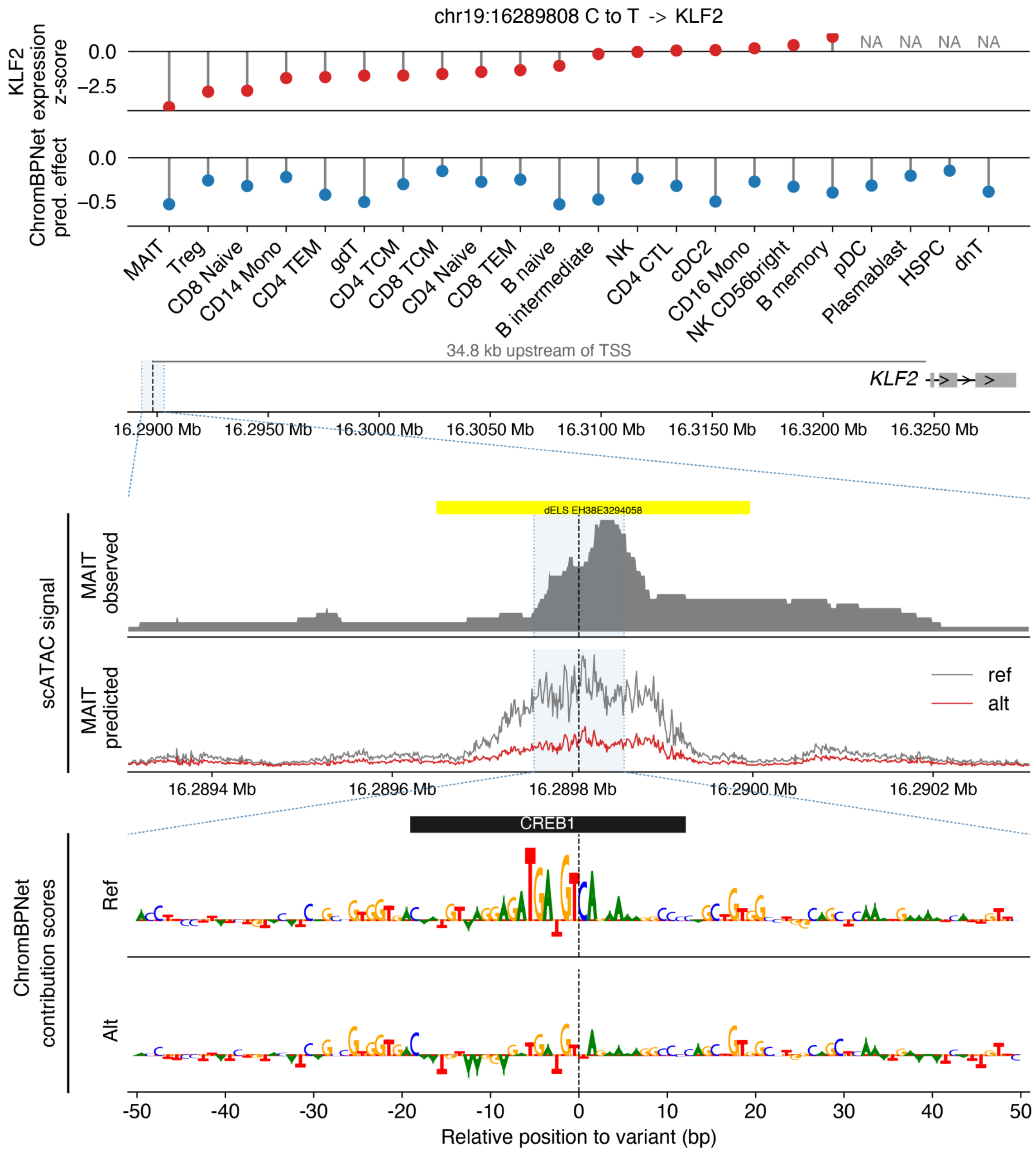


**Supplementary Figure 17. Rare caQTL in the distal enhancer of *KLF2* and *CREB1* binding site**. (A) Red lollipop plot of target gene expression z-scores, ordered from the most to the least affected. The expression z-score represents the difference in expression for individuals carrying the alternative allele relative to those carrying the reference allele. Blue lollipop plot represents the difference in ChromBPNet predicted chromatin accessibility. (B) Genomic context of the variant relative to the target gene. The black dashed line marks the variant position. (C) Observed and predicted cell-type–specific scATAC-seq signal in the cellular context showing the largest ChromBPNet-predicted log-counts difference between the alternative and reference alleles, plotted over a ±500 bp window centered on the variant. The black dashed line indicates the variant position. The H3K27Ac peaks as ENCODE dELS (distal enhancer-like sequence) are annotated as yellow bars. (D) ChromBPNet contribution scores for the reference and alternative alleles across a ±50 bp window centered on the variant. The highest-scoring transcription factor motif overlapping the variant, identified from the reference sequence contribution scores, is shown. The black dashed line indicates the variant position.

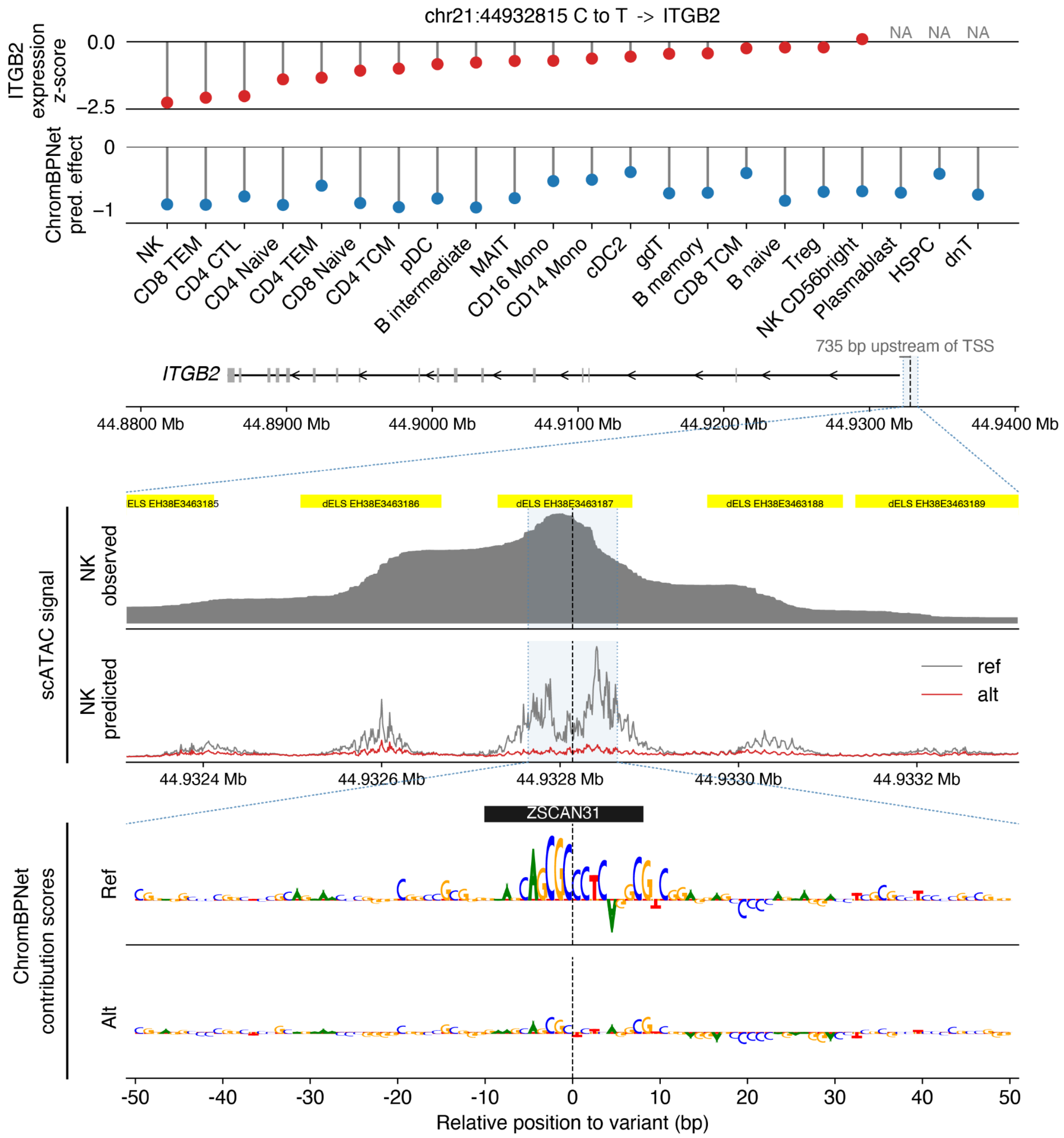


**Supplementary Figure 18. Rare caQTL in the distal enhancer of *ITGB2* and *ZSCAN31* binding site**. (**A**) Red lollipop plot of target gene expression z-scores, ordered from the most to the least affected. The expression z-score represents the difference in expression for individuals carrying the alternative allele relative to those carrying the reference allele. Blue lollipop plot represents the difference in ChromBPNet predicted chromatin accessibility for peak chr21:44932408–44933489. (**B**) Genomic context of the variant relative to the target gene. The black dashed line marks the variant position. (**C**) Observed and predicted cell type–specific scATAC-seq signal in the cellular context showing the largest ChromBPNet-predicted log-counts difference between the alternative and reference alleles, plotted over a ±500 bp window centered on the variant. The black dashed line indicates the variant position. The H3K27Ac peaks as ENCODE dELS (distal enhancer-like sequence) are annotated as yellow bars. (**D**) ChromBPNet contribution scores for the reference and alternative alleles across a ±50 bp window centered on the variant. The highest-scoring transcription factor *ZSCAN31* motif overlapping the variant, identified from the reference sequence contribution scores, is shown. The black dashed line indicates the variant position.

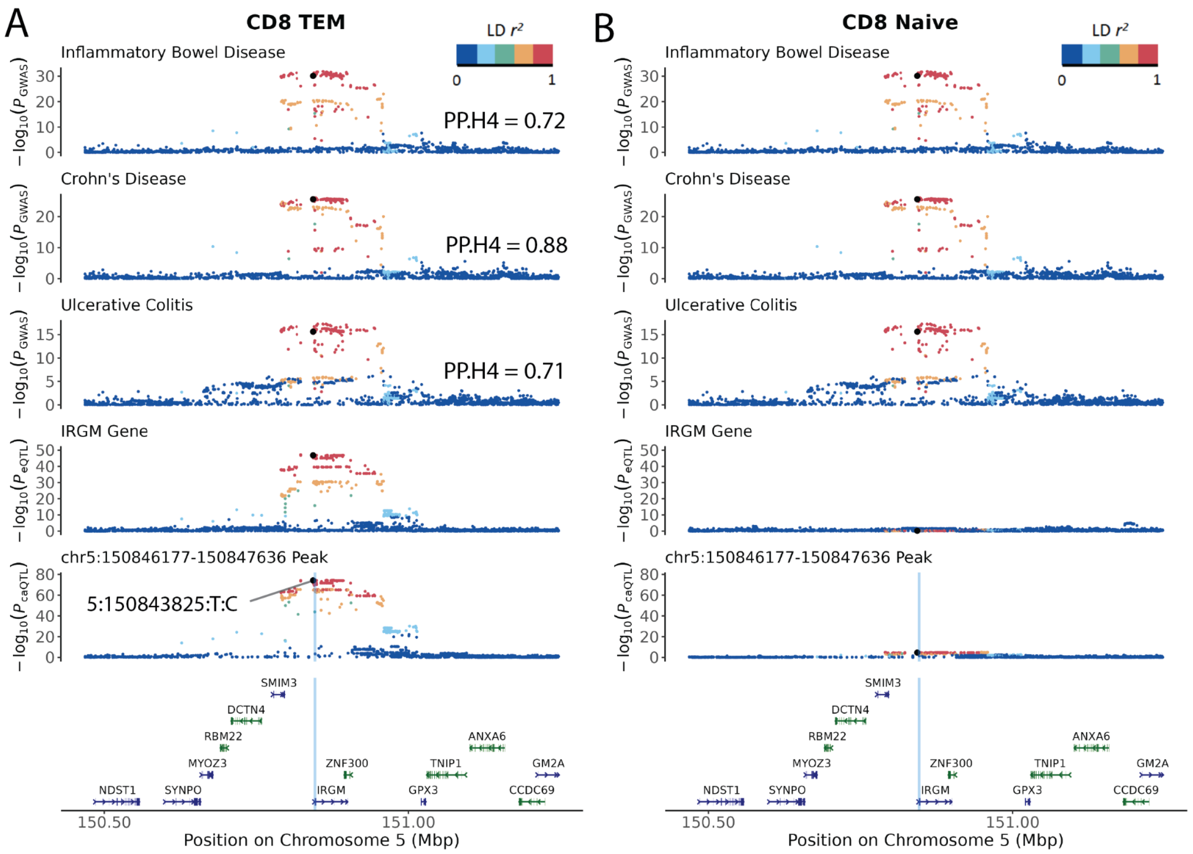


**Supplementary Figure 19**. **Cell-type-specific pattern of peak-to-gene links for gene *IRGM*.** Locus plots of GWAS, eQTL and caQTL in $CD8_{TEM}$ and $CD8_{Naive}$ cells. The six layers from top to bottom are IBD GWAS, CD GWAS, UC GWAS, eQTL of *IRGM*, caQTL of peak chr5:150846177-150847636, and gene location track. (**A**) Locus plot in $CD8_{TEM}$ cells. The y-axis indicates the $-\log_{10}(p_{GWAS})$, $-\log_{10}(p_{eQTL})$ and $-\log_{10}(p_{caQTL})$ repestively. The coloc PP.H4 results are annotated for each of the three GWAS. The x-axis demonstrates the genomic location of chromosome 5. The top caQTL variant (5:150843825:T:C) is highlighted and the vertical blue line indicates the location of the caPeak. The actual top caQTL variant is an INDEL variant, 5:150843825:T:C, but it not observed in the GWAS datasets. So for visualization, we only highlighted the top SNP variant. The color of each dot indicates the LD $r^2$ with the top caQTL variant. (**B**) Locus plot of the same six layers in $CD8_{Naive}$ cells. The three GWAS layers are exactly the same as panel A.

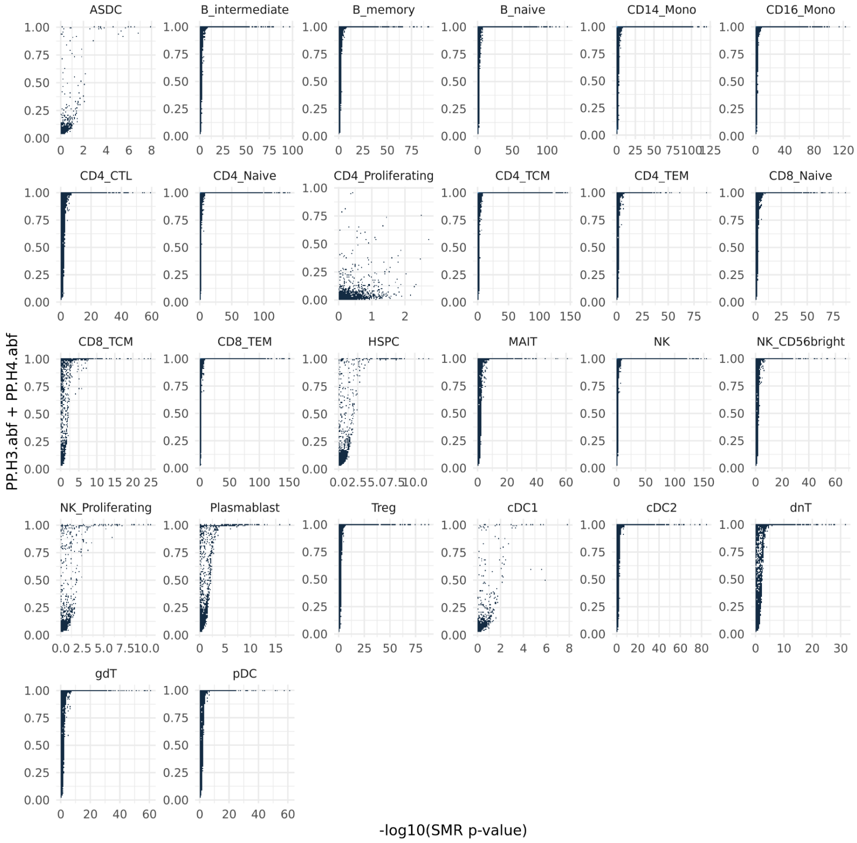
ASDC
B_intermediate
B_memory
B_naive
CD14_Mono
CD16_Mono
CD4_CTL
CD4_Naive
CD4_Proliferating
CD4_TCM
CD4_TEM
CD8_Naive
CD8_TCM
CD8_TEM
HSPC
MAIT
NK
NK_CD56bright
NK_Proliferating
Plasmablast
Treg
cDC1
cDC2
dnT
gdT
pDC
PP.H3.abf + PP.H4.abf
-log10(SMR p-value)

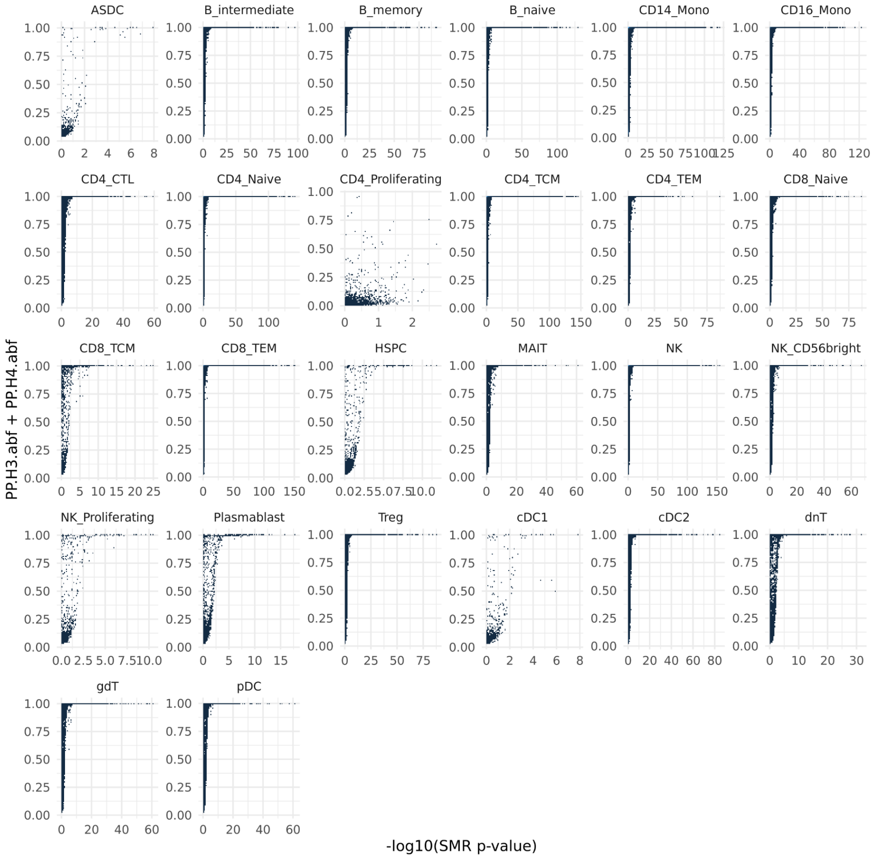
ASDC
B_intermediate
B_memory
B_naive
CD14_Mono
CD16_Mono
CD4_CTL
CD4_Naive
CD4_Proliferating
CD4_TCM
CD4_TEM
CD8_Naive
CD8_TCM
CD8_TEM
HSPC
MAIT
NK
NK_CD56bright
NK_Proliferating
Plasmablast
Treg
cDC1
cDC2
dnT
gdT
pDC
PP.H3.abf + PP.H4.abf
-log10(SMR p-value)

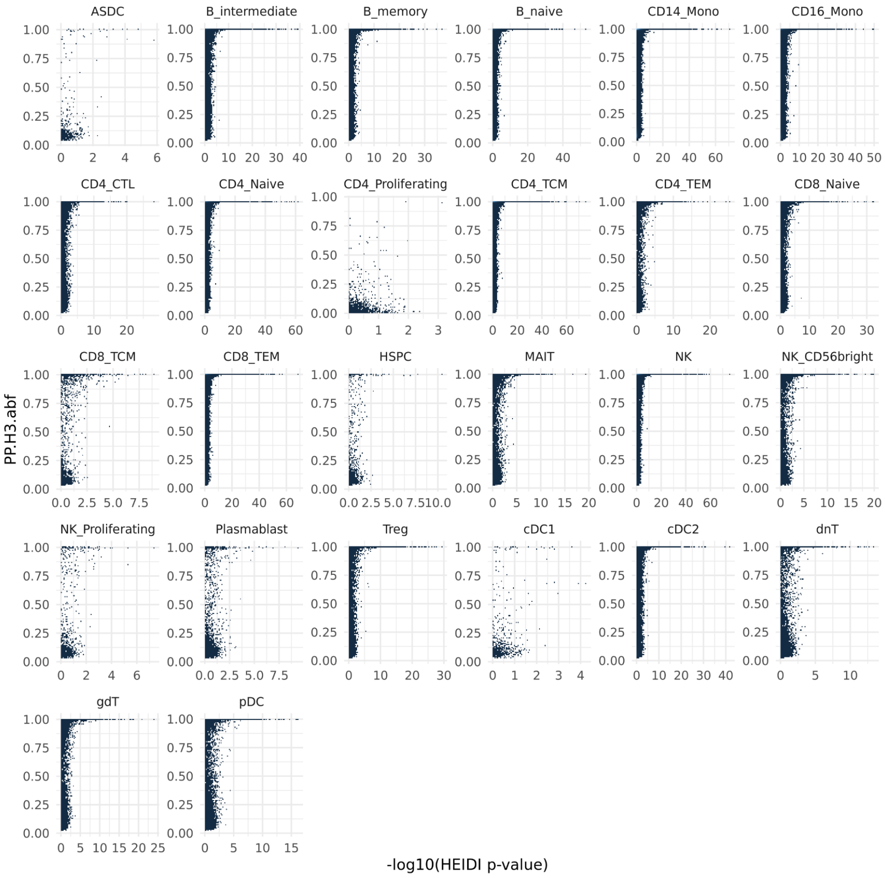


**Supplementary Figure 20. Comparison between coloc and SMR regarding different metrics**. This plot shows the coloc and SMR results for peak-gene links across 26 cell types. Each dot indicates a peak-gene pair. (Top panel) The x-axis indicates the -log10($p_{\text{SMR}}$) and the y-axis indicates the PP.H3 + PP.H4 from coloc. (Middle panel) The x-axis indicates the -log10(pSMR) and the y-axis indicates the PP.H4 from coloc. (Bottom panel) The x-axis indicates the -log10($p_{\text{HEIDI}}$) and the y-axis indicates the PP.H3 from coloc.

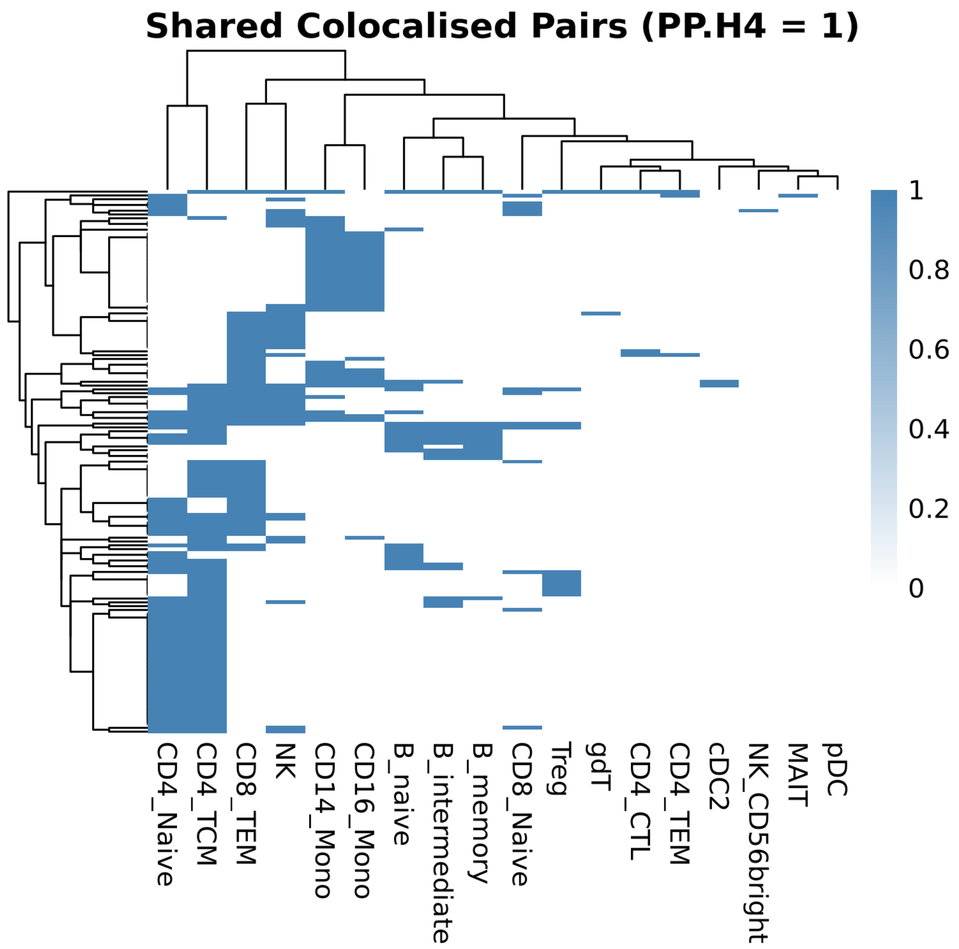


**Supplementary Figure 21. Strongly colocalized peak-to-gene pairs are shared across different cell types**. Heatmap of PP.H4 values for 446 peak-to-gene pairs with PP.H4 =1 in at least one cell type. Rows represent peak–gene pairs; columns represent cell types. Values indicate posterior probability of a shared causal variant per cell type, highlighting lineage-specific and shared regulatory links

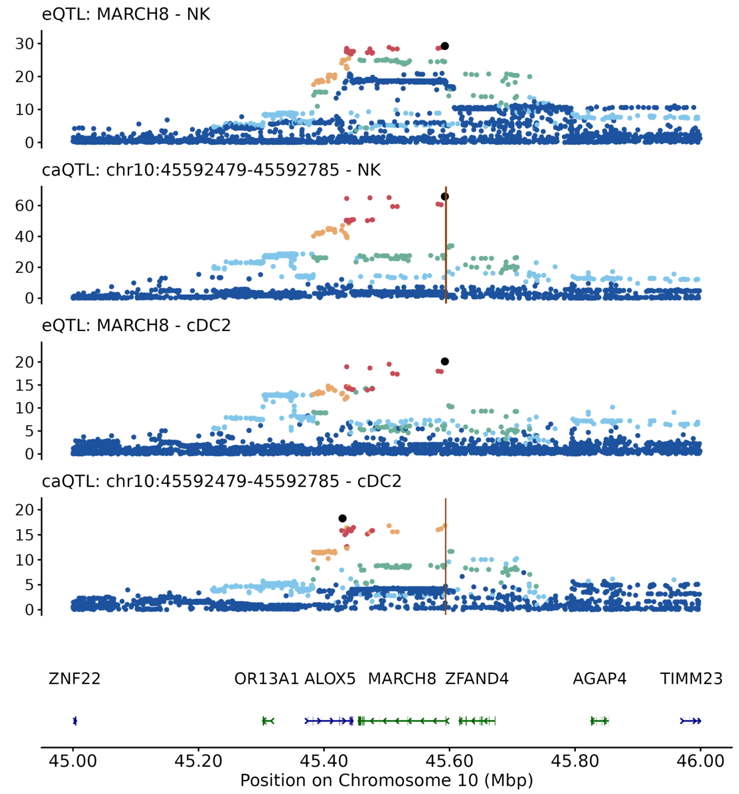


**Supplementary Figure 22. Locus plot of *MARCH8* in NK cells and cDC2**. Top: The eQTL and caQTL locus plot for gene *MARCH8* and peak chr10:45592479-45592785 in NK cells. Mid: the same gene and peak in cDC2. Bottom: the genome track at this locus. The top variant is highlighted in purple, and other SNPs are colored based on their LD strength with the top variant. The vertical brown line indicates the location of the peak.

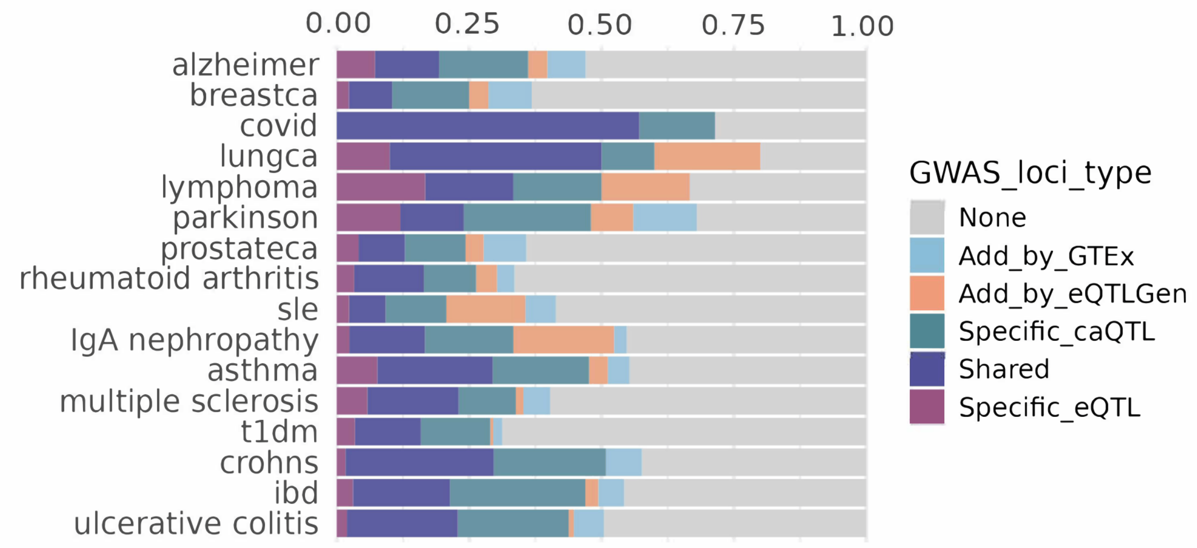


**Supplementary Figure 23. GWAS colocalization pattern using TenK10K eQTL and caQTL as baseline.** The stacked bar plot indicates the percentage of GWAS loci showing significant colocalization with eQTL or caQTL datasets. The baseline is TenK10K sc- eQTL and caQTL. The "Shared" group indicates those loci colocalizing with both TenK10K sc-eQTL and caQTL. Additional datasets are included in the order of bulk eQTLs from the eQTLGen consortium and GTEx whole body tissues.

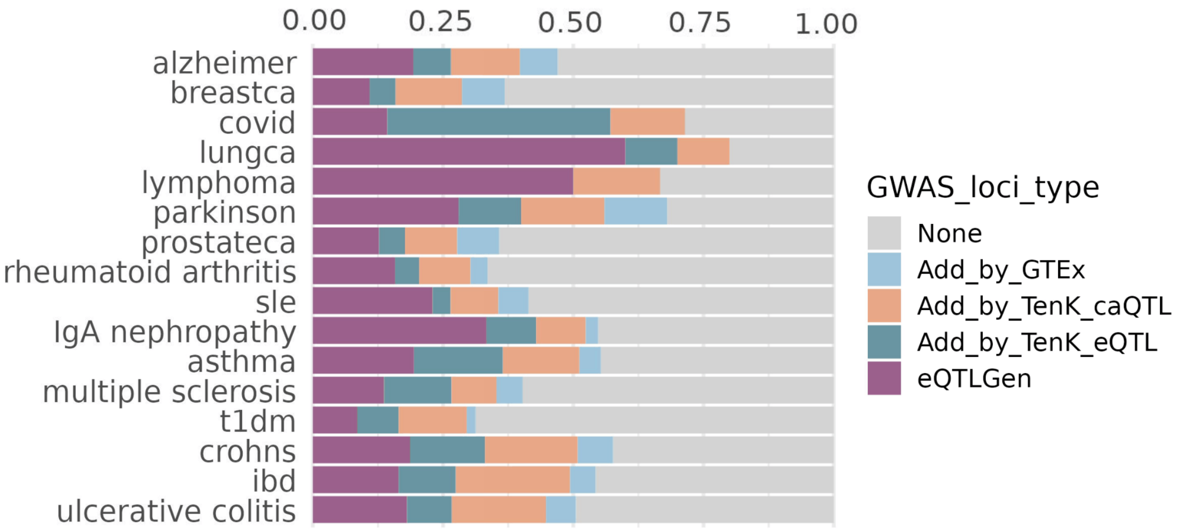


**Supplementary Figure 24. GWAS and xQTL colocalization pattern using eQTLGen as the baseline.** The stacked bar plot indicates the percentage of GWAS loci showing significant colocalization with eQTL or caQTL datasets. The baseline is the bulk eQTL from the eQTLGen consortium. Additional datasets are included in the order of TenK10K sc- eQTL, caQTL, and bulk eQTL from GTEx whole body tissues.

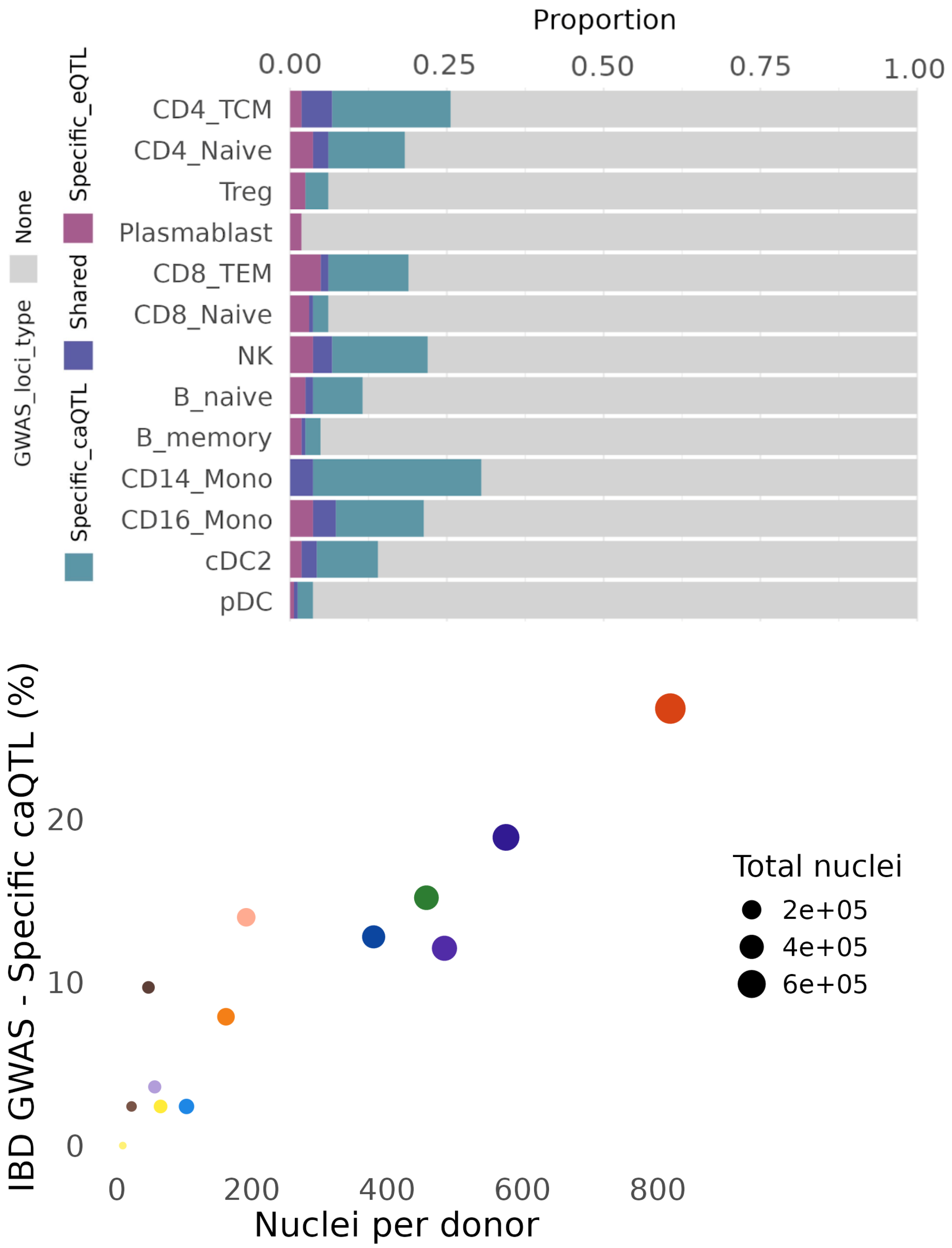


**Supplementary Figure 25. Cell type-specific colocalization signals for IBD GWAS**. Top: The stacked bar plot depicting the % of eQTL and caQTL colocalization signals with IBDG GWAS in each cell type; Bottom: the percentage of caQTL-specific colocalization per cell type is dependent on the cell type abundance level. The dot size represents the total number of nuclei for the cell type. The color of the dot indicates the cell type and the color scheme is consistent with Figure 2B.

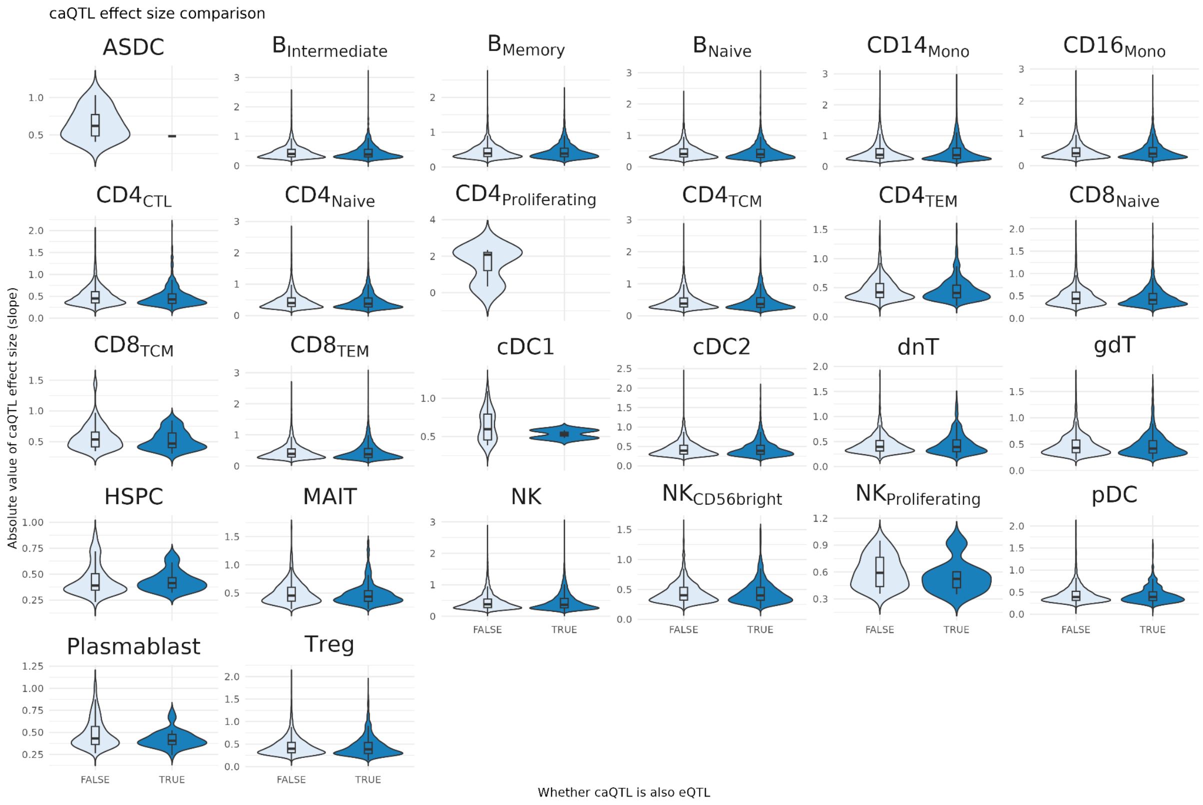


**Supplementary Figure 26. Effect size comparison of caQTLs with and without eQTL overlap**. The x-axis represents two groups of caQTLs that overlap with eQTL or not. The y-axis indicates the effect size of the caQTL, which is the slope estimate from TensorQTL. Each panel indicates the results from one of the 26 cell types.

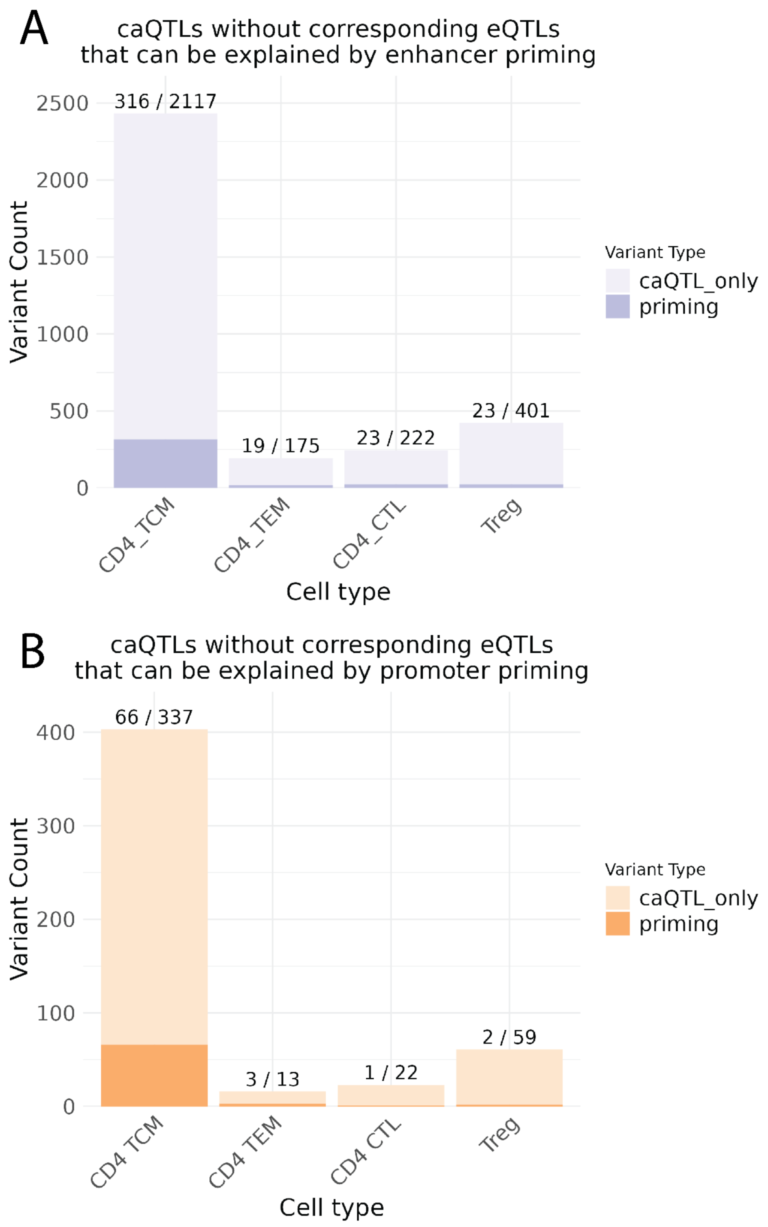


**Supplementary Figure 27. Enhancer and promoter priming of caQTLs without corresponding eQTLs in the CD4+ T cell population**. Summary of the number of caQTLs without corresponding eQTLs in each differentiated CD4+ T cell type compared to $CD4_{Naive}$ cell as baseline. The dark stack bar indicates the number due to priming and the light stack bar indicates those that are not. Panel A represents the results for enhancer priming, and panel B represents the results for promoter priming.

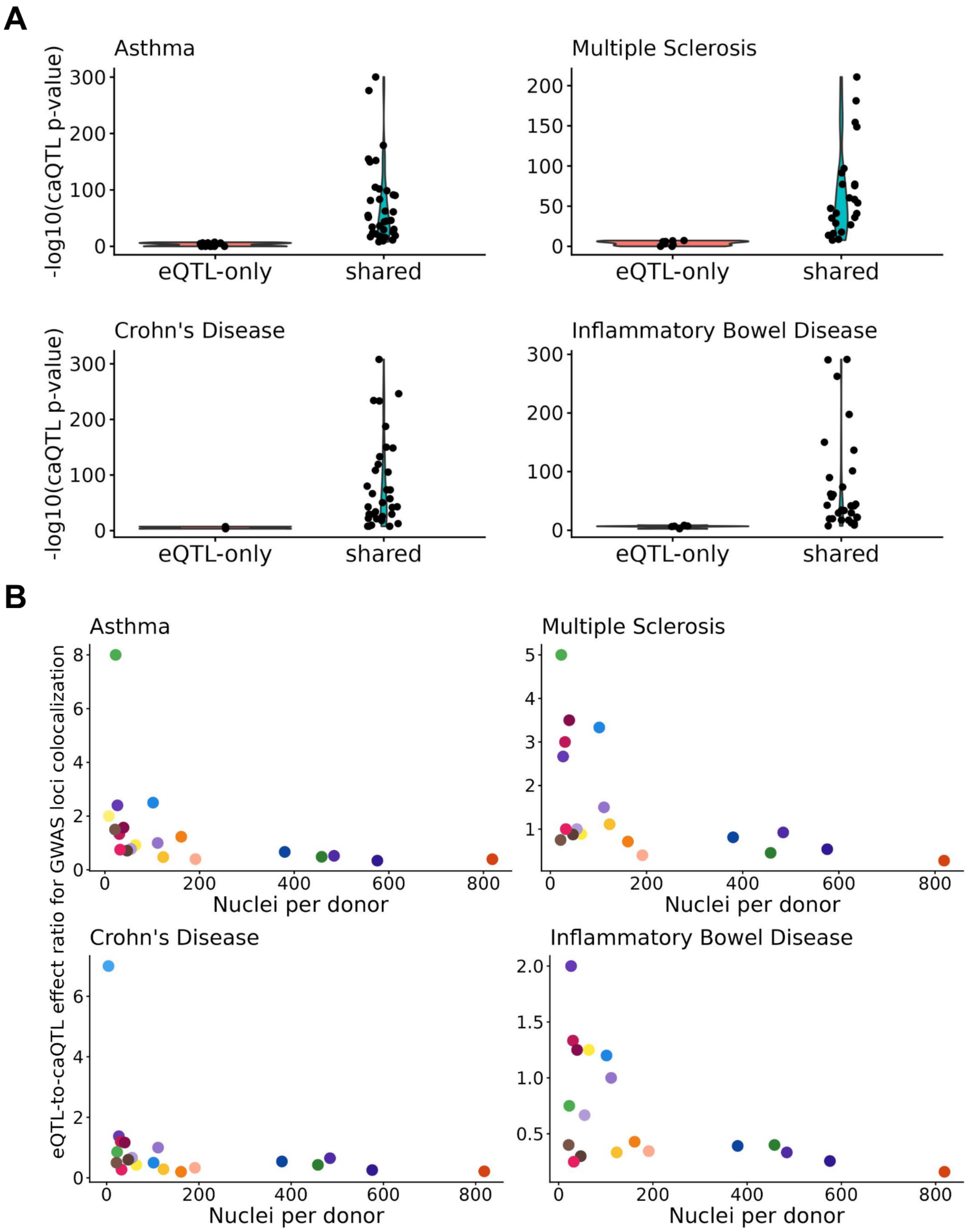


**Supplementary Figure 28. GWAS colocalization only with eQTL due to power limit.** (**A**) Evaluation of caQTL significance level between GWAS loci colocalized with both caQTL and eQTL versus those colocalized with eQTL only. The red violin group indicates the GWAS loci with eQTL-only colocalization. Blue violin group indicates the GWAS loci with both eQTL and caQTL colocalization. The y-axis indicates the -log10 of the caQTL nominal p-value. (**B**) Comparison of the proportion of GWAS loci that colocalize with eQTLs versus those that colocalize with caQTLs across different cell types. The y-axis represents the ratio, and the x-axis indicates the number of nuclei per donor for each cell type. Each colored dot corresponds to a distinct cell type.

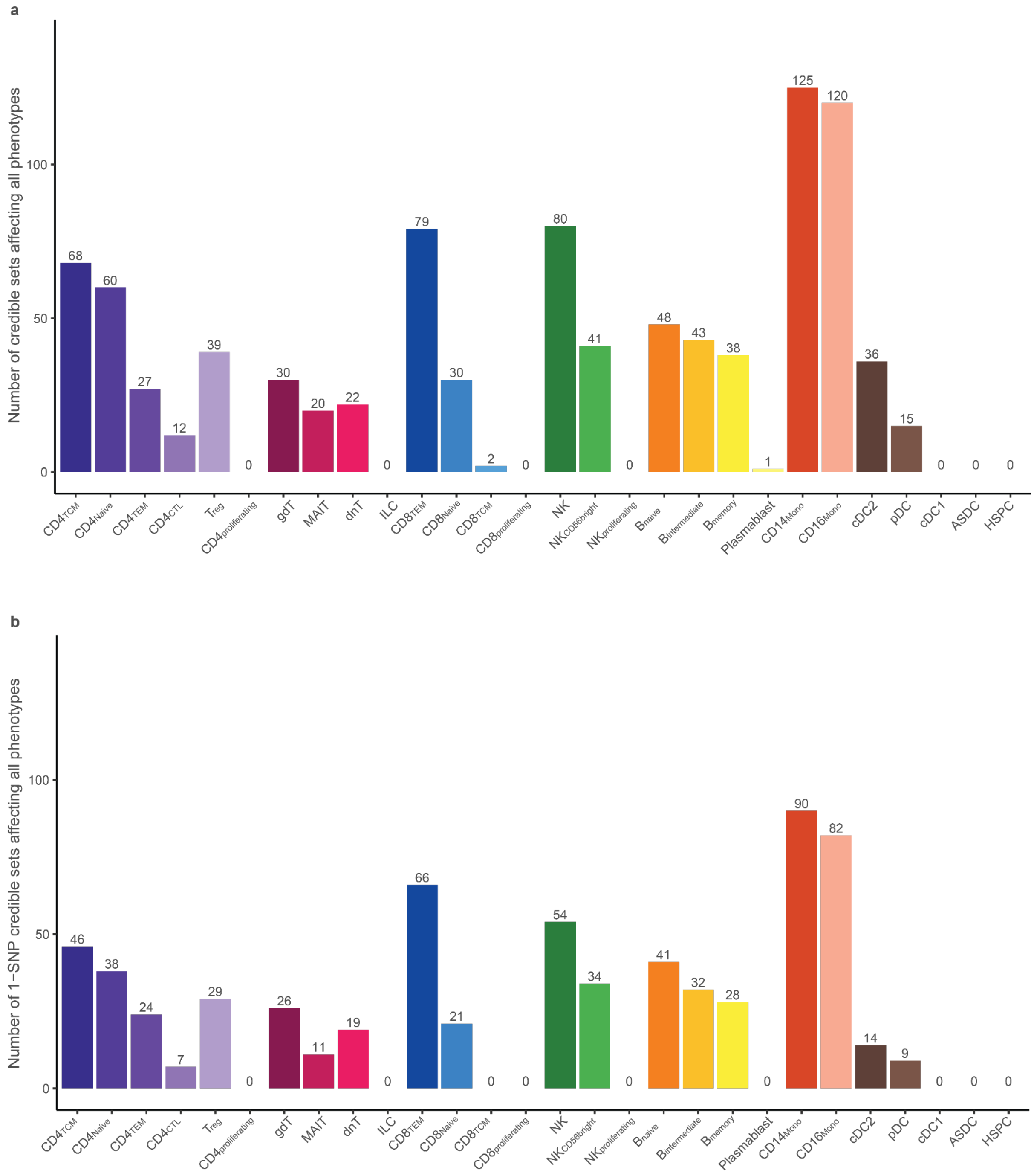


**Supplementary Figure 29. Summary of fine-mapping results by *mvSuSiE*. (a)** Unique credible sets that affect disease risk, gene expression and chromatin accessibility simultaneously. Each bar indicates the number of credible sets from fine-mapping. **(b)** The summary of 1-SNP credible sets from panel **(a)**. Only 19 cell types were detected with this type of credible set.

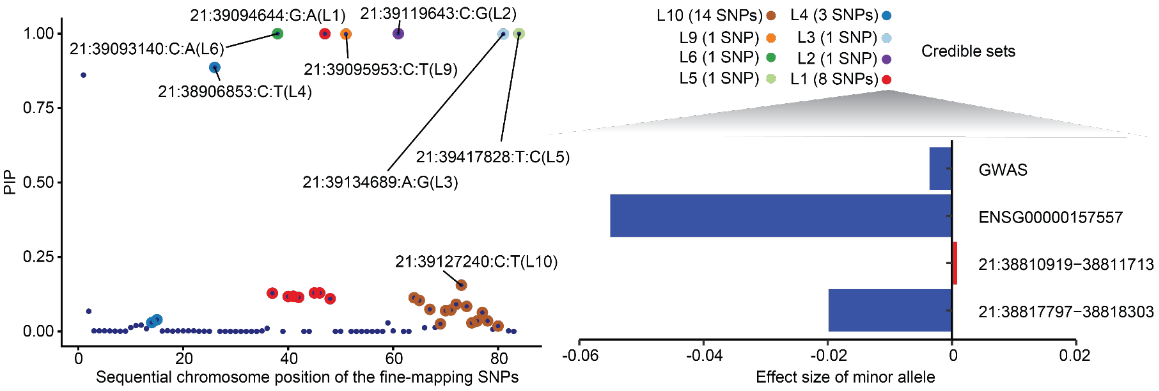


**Supplementary Figure 30.** Fine-mapping results of *ETS2* locus using *mvSuSiE* in $CD14_{Mono}$, identifying a credible set (L1) that affects IBD risk, gene expression and chromatin accessibility with posterior inclusion probability (PIP) = 1 for the causal SNP. The zoom-in session shows the estimated effect sizes of minor allele from the causal variant on different phenotypes.

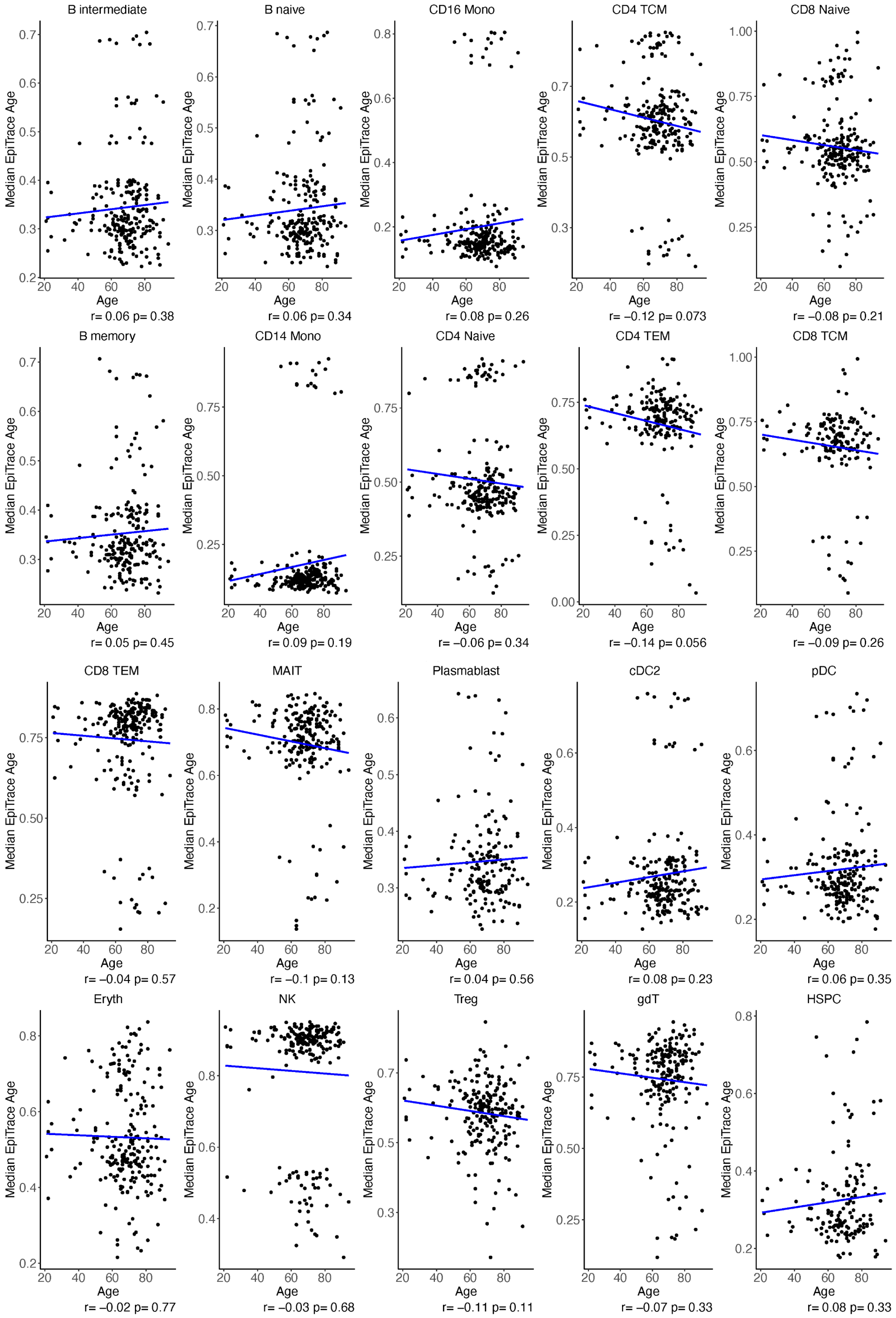
B intermediate
r= 0.06 p= 0.38
B naive
r= 0.06 p= 0.34
CD16 Mono
r= 0.08 p= 0.26
CD4 TCM
r= −0.12 p= 0.073
CD8 Naive
r= −0.08 p= 0.21
B memory
r= 0.05 p= 0.45
CD14 Mono
r= 0.09 p= 0.19
CD4 Naive
r= −0.06 p= 0.34
CD4 TEM
r= −0.14 p= 0.056
CD8 TCM
r= −0.09 p= 0.26
CD8 TEM
r= −0.04 p= 0.57
MAIT
r= −0.1 p= 0.13
Plasmablast
r= 0.04 p= 0.56
cDC2
r= 0.08 p= 0.23
pDC
r= 0.06 p= 0.35
Eryth
r= −0.02 p= 0.77
NK
r= −0.03 p= 0.68
Treg
r= −0.11 p= 0.11
gdT
r= −0.07 p= 0.33
HSPC
r= 0.08 p= 0.33
Median EpiTrace Age
Age

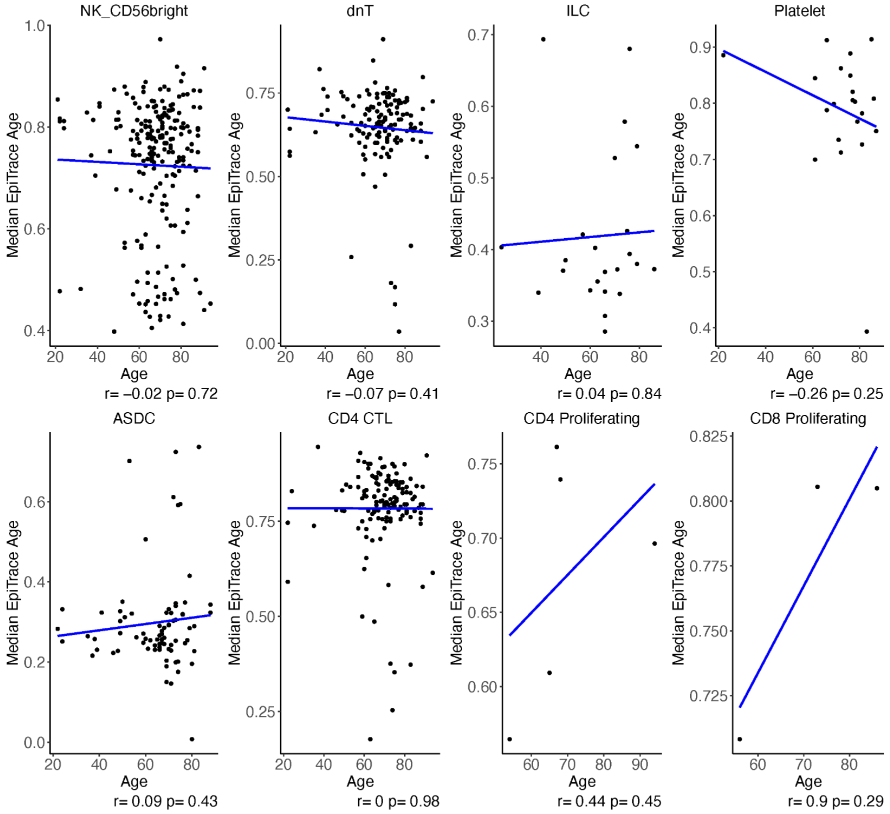


**Supplementary Figure 31. Correlation between chronological age and estimated epigenetic age across donors**. The x-axis indicates the chronological age of each donor. The y-axis represents the median of estimated epigenetic age across cells of each donor. Pearson's correlation coefficient (r) and its p-value are printed under each panel.

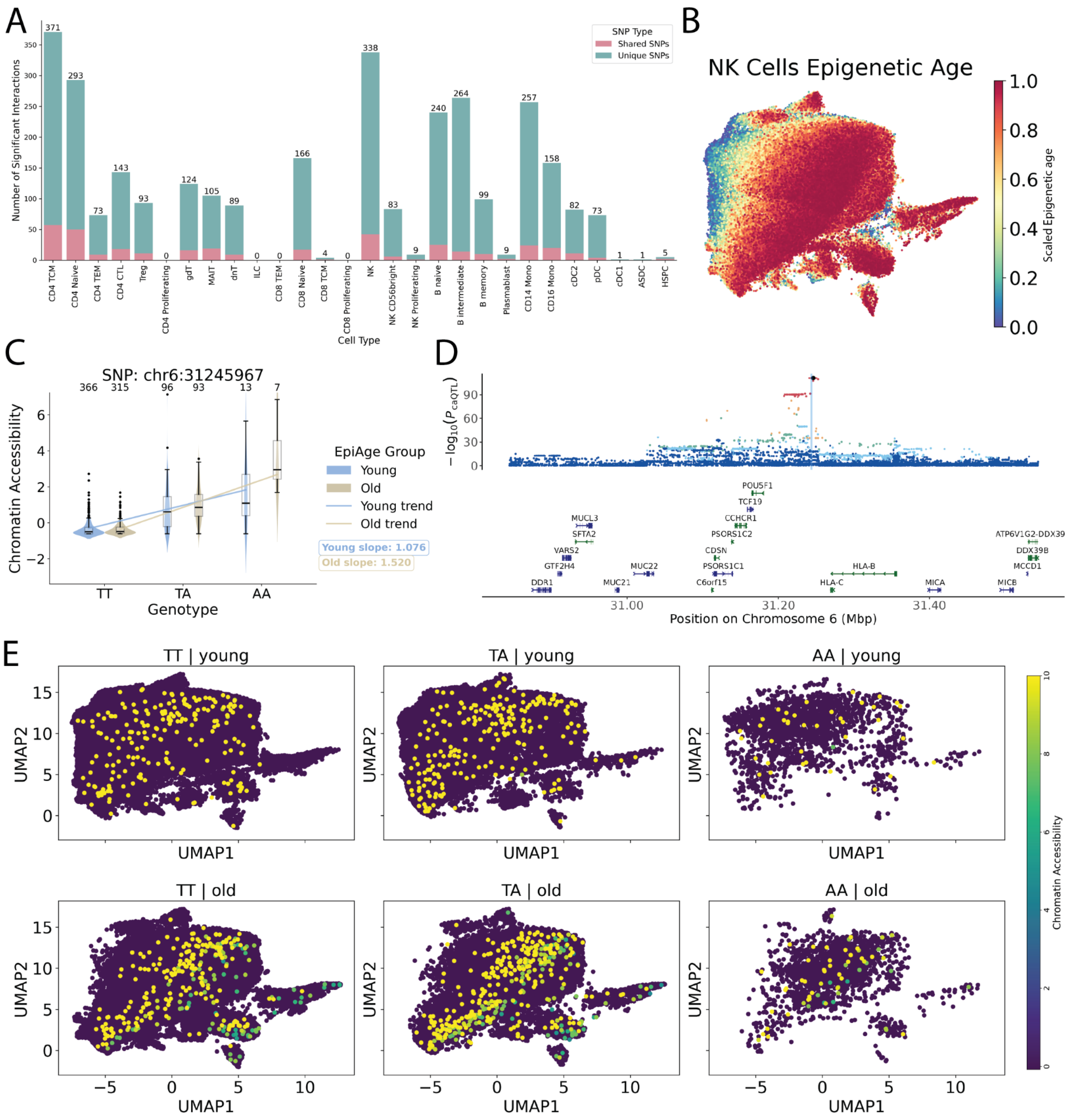


**Supplementary Figure 32. Genetic effects on chromatin accessibility vary with epigenetic age, revealing context-dependent regulatory patterns**. (**A**) Number of caQTLs showing significant genotype-by-epigenetic-age interaction (FDR < 0.05) across all immune cell types and breakdown of significant caQTL–age interactions by specificity: number of interactions unique to a single cell type versus those shared across multiple cell types. (**B**) Density plot of the inferred epigenetic age within the NK cell population (N = 205,330). The color of the bin indicates the scaled (0-1) epigenetic ages, with larger values indicating epigenetically older cells. (**C**) Example of an EpiAge-dependent caQTL (chr6:31243369-31243947) in NK cells. Boxplot shows chromatin accessibility at a peak in the *HLA-C* locus (stratified by 6:31245967:T:A genotype and epigenetic age group, illustrating increased effect size in epigenetically older cells. (**D**) Locus plot of the caQTL. The upper panel depicts the significant level of all associated variants. The color of the dots indicates the LD with the top variant. The x-axis indicates the genomic location and the y-axis indicates -$\log_{10}$ of the nominal *p*-values.

The vertical blue line represents the location of the caPeak. The bottom panel shows the gene location track. (**E**) Density plot of chromatin accessibility separated by genotype of the SNP 6:31245967:T:A and epigenetic age groups (old vs young). Each dot indicates a cell and the color represents the scaled (0-10) chromatin accessibility the caPeak chr6:31243369-31243947, with a larger value indicating higher accessibility.

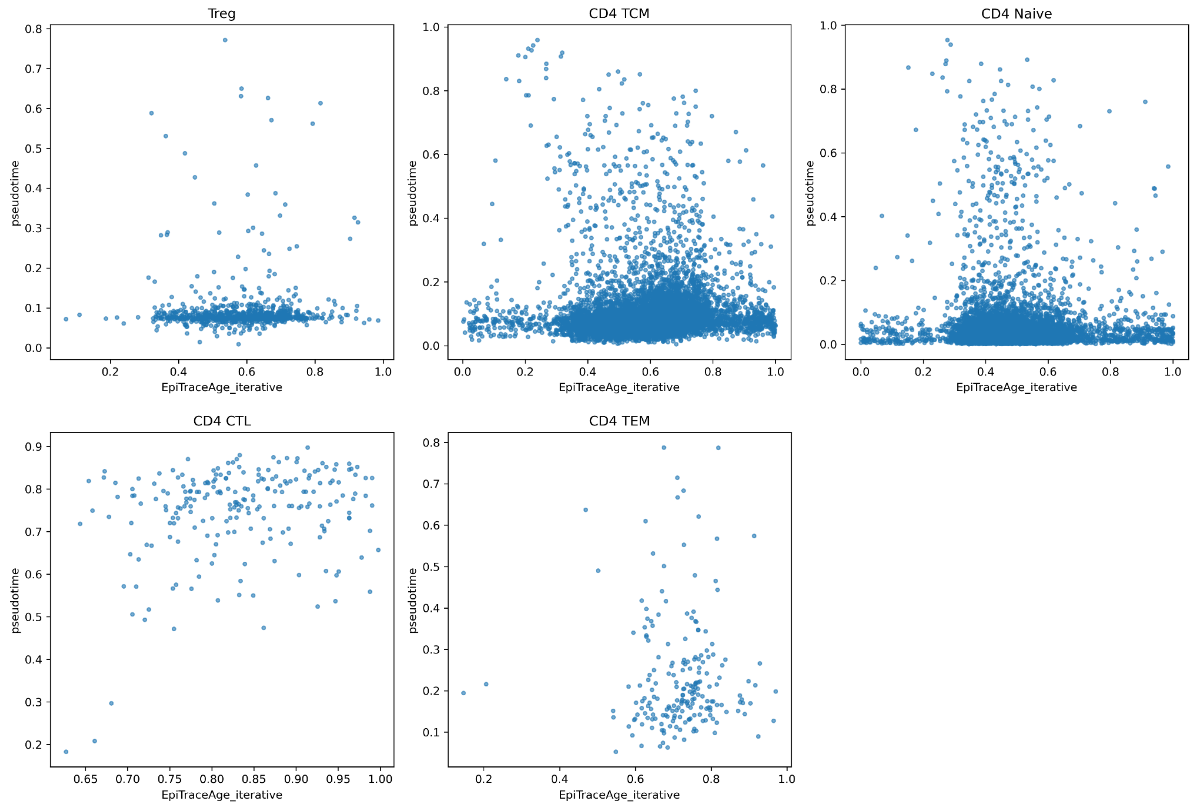


**Supplementary Figure 33. Comparison between Epigenetic age and pseudotime for the CD4+ T cell population at the cellular level**. The x-axis represents the Epigenetic age, and the y-axis represents the inferred pseudotime. Each panel indicates one of the five cell types in the CD4+ T cell population.

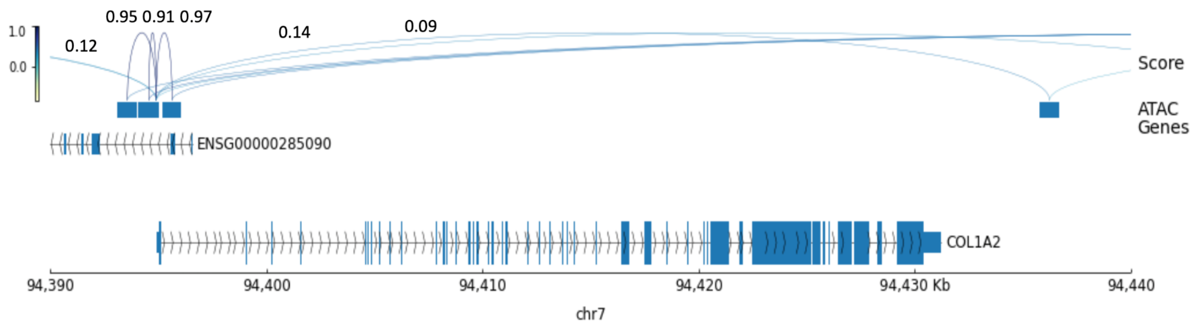


**Supplementary Figure 34.** Genomic tracks of the *COL1A2* gene and neighboring regions with predicted GLUE regulatory scores.

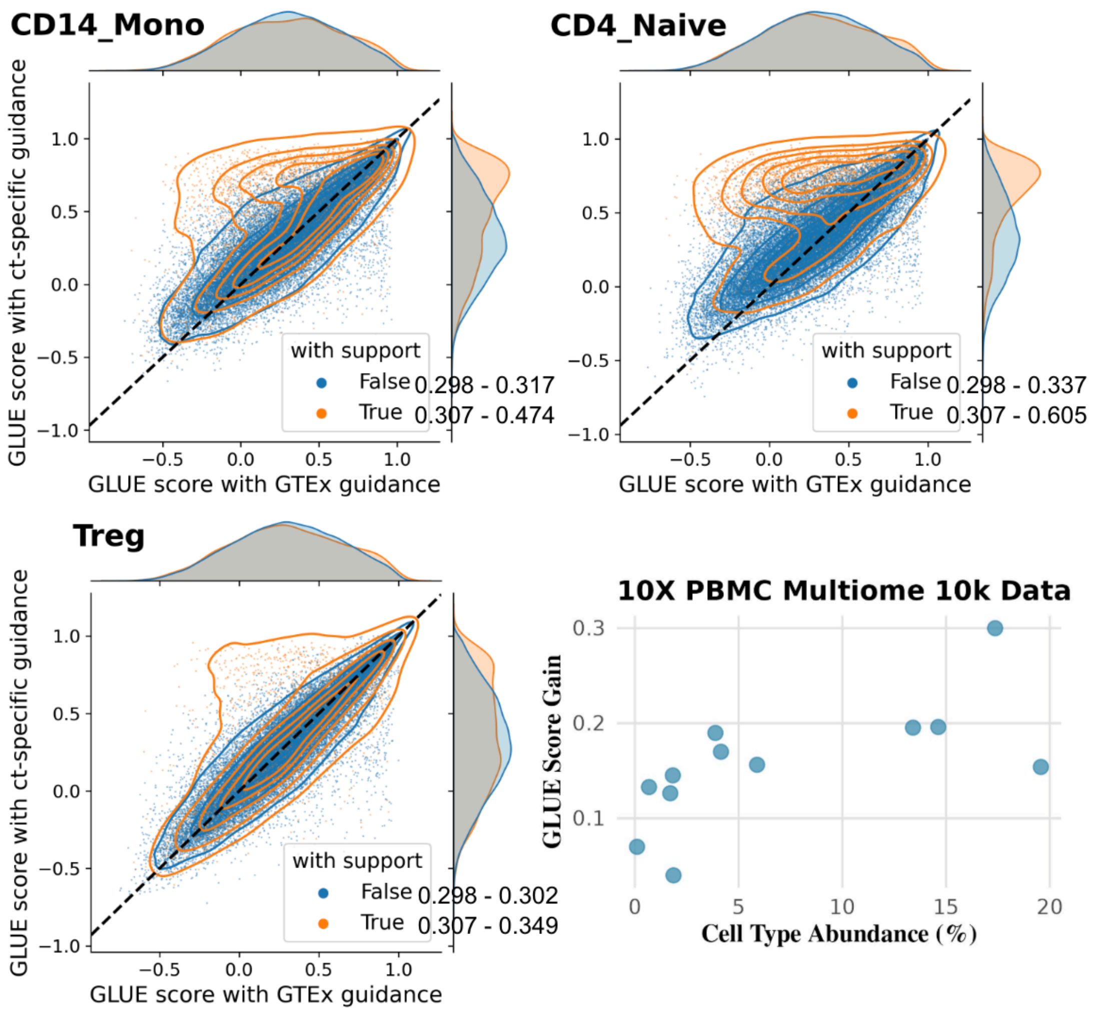


**Supplementary Figure 35.** Extended evaluation of the impact of different guidance for cis-regulatory inference, using a subset of TOB comparable in size to the 10X dataset. The scatter plot provides a broader view of GLUE score improvements by feeding eQTLs across 12 cell types with varying abundance.

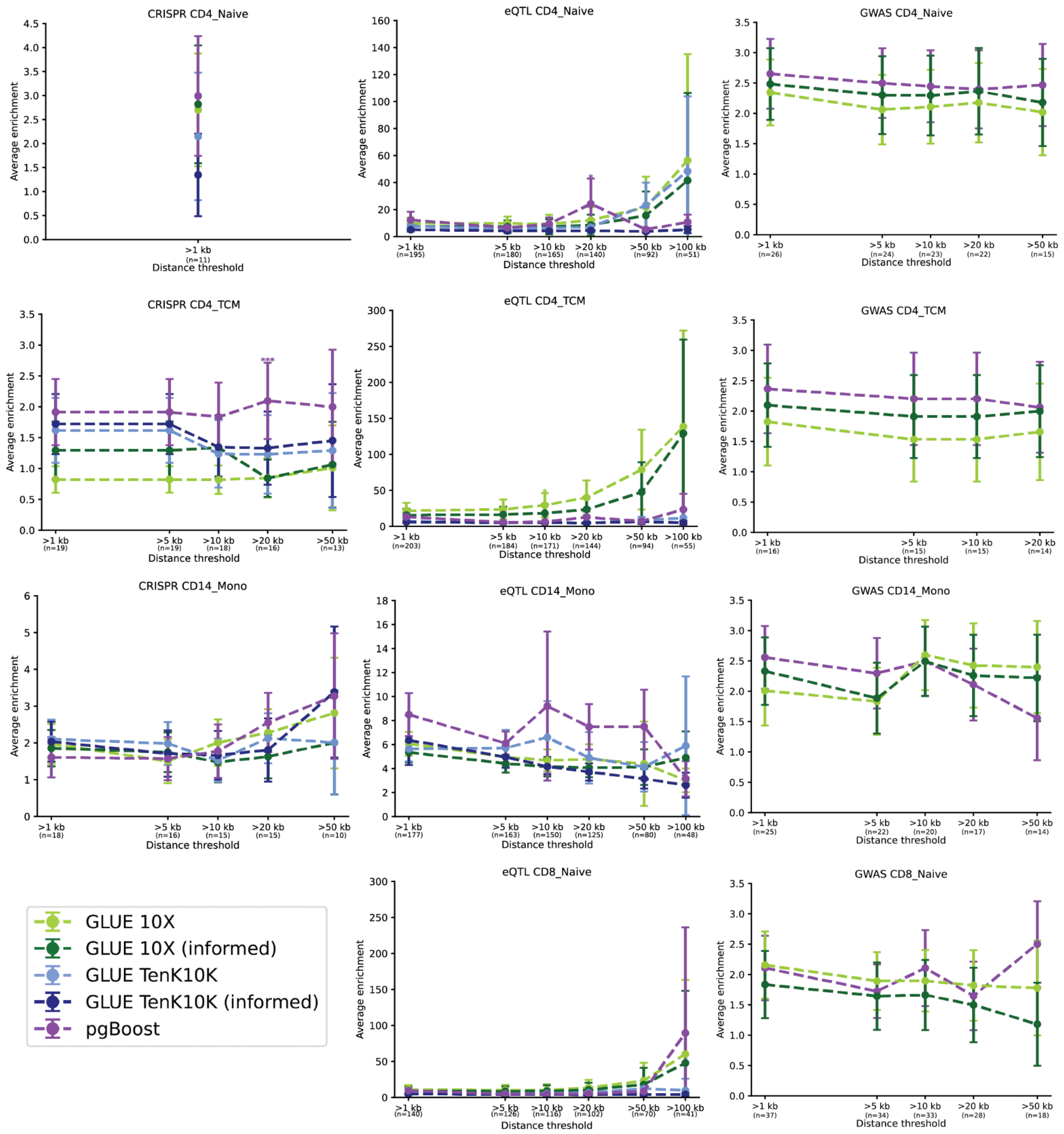


**Supplementary Figure 36. Benchmarking performance of peak-to-gene links between pgBoost and GLUE using three validation datasets**. The three columns indicate the three different validation datasets (CRISPR, eQTL, and GWAS). The four rows indicate different cell types tested. In each panel, the x-axis represents the peak-to-gene distance and the number of links tested (n). The y-axis represents the average enrichment score calculated by pgBoost. The error bar indicates the standard error of the score. The color of the dots represents different methods used.

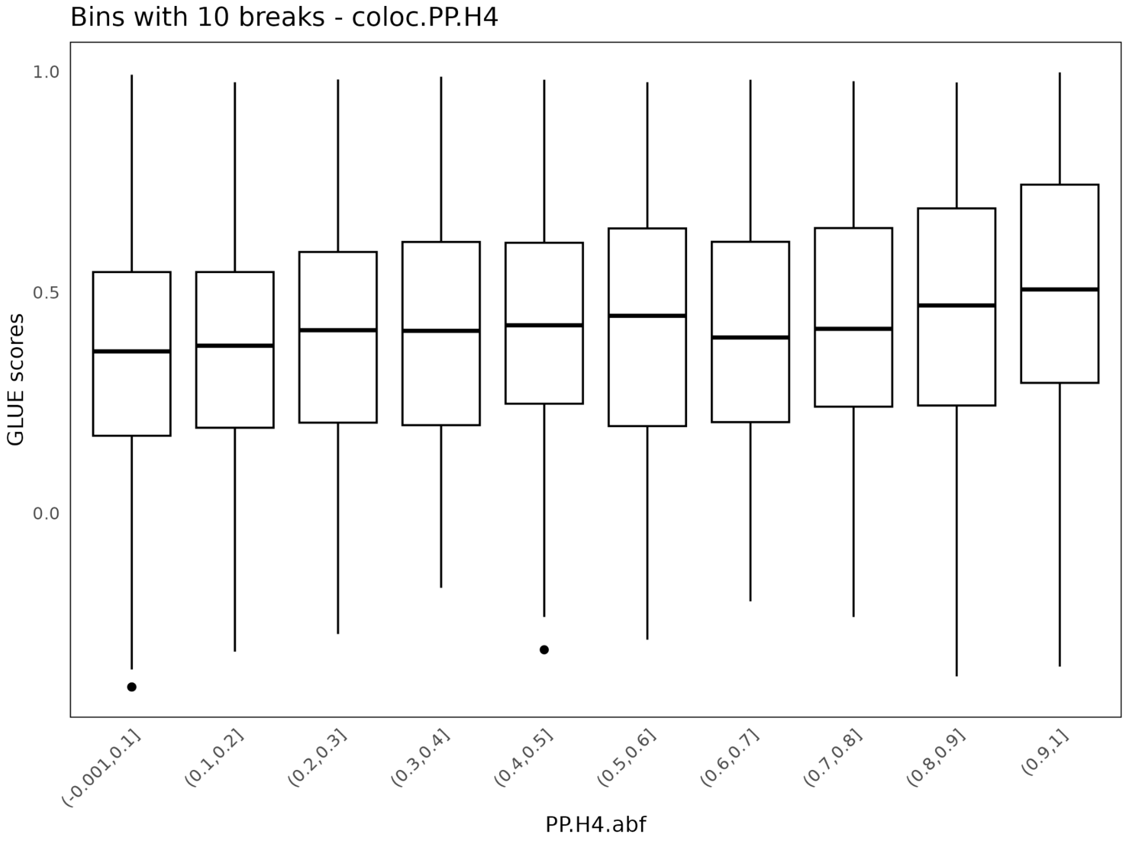


**Supplementary Figure 37. Box plots of GLUE scores and coloc PP.H4 values.** Data are binned by PP.H4.abf in intervals of 0.1.

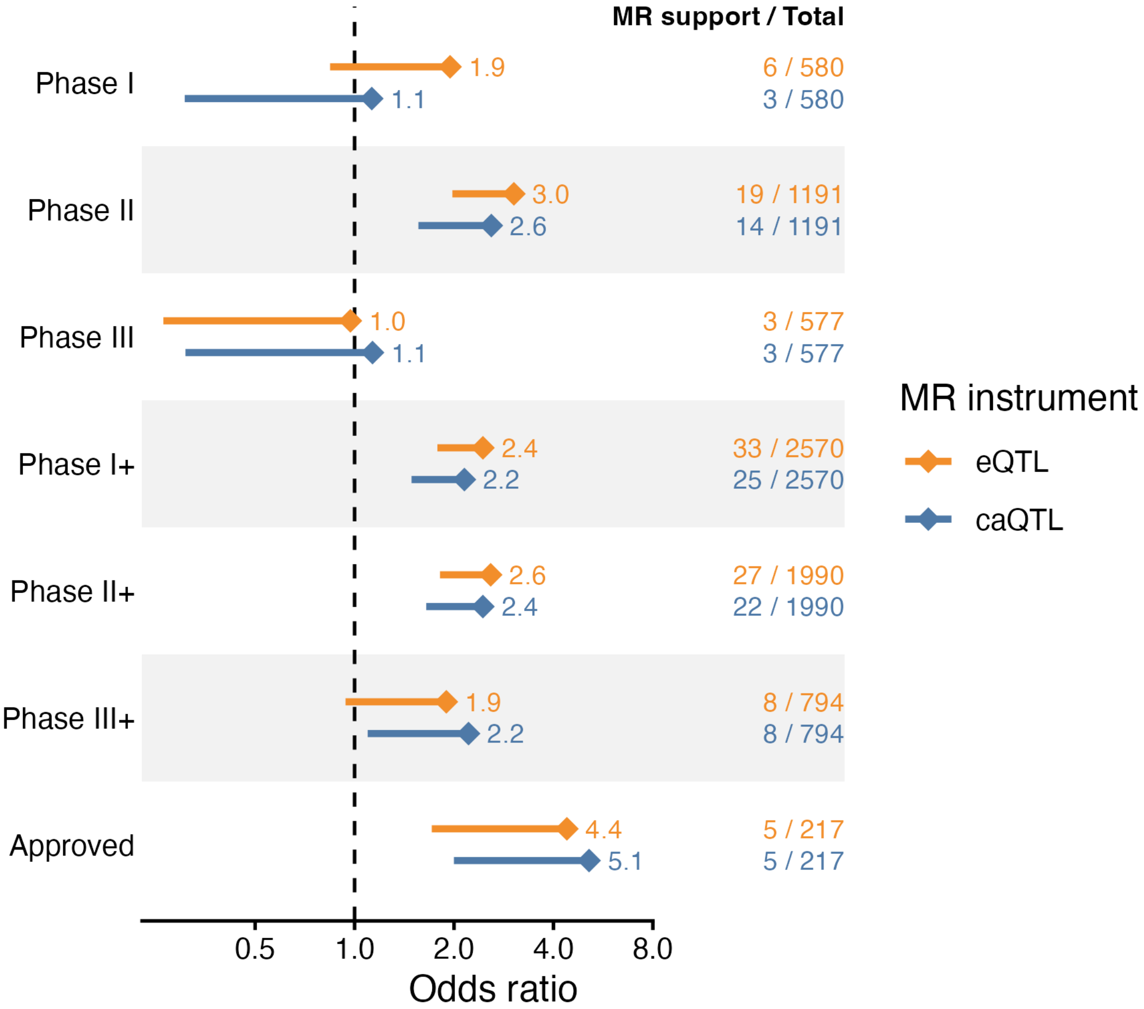


**Supplementary Figure 38. Drug enrichment of candidate genes derived from eQTL and caQTL SMR analysis**. Enrichment of SMR-supported target–indication pairs across clinically investigated indications. Odds ratios (ORs) with lower 95% confidence intervals are shown comparing pairs with versus without SMR support, stratified by their highest clinical development phase. Annotations on the right indicate the number of SMR-supported pairs relative to the total number of clinically investigated pairs in each phase or category.

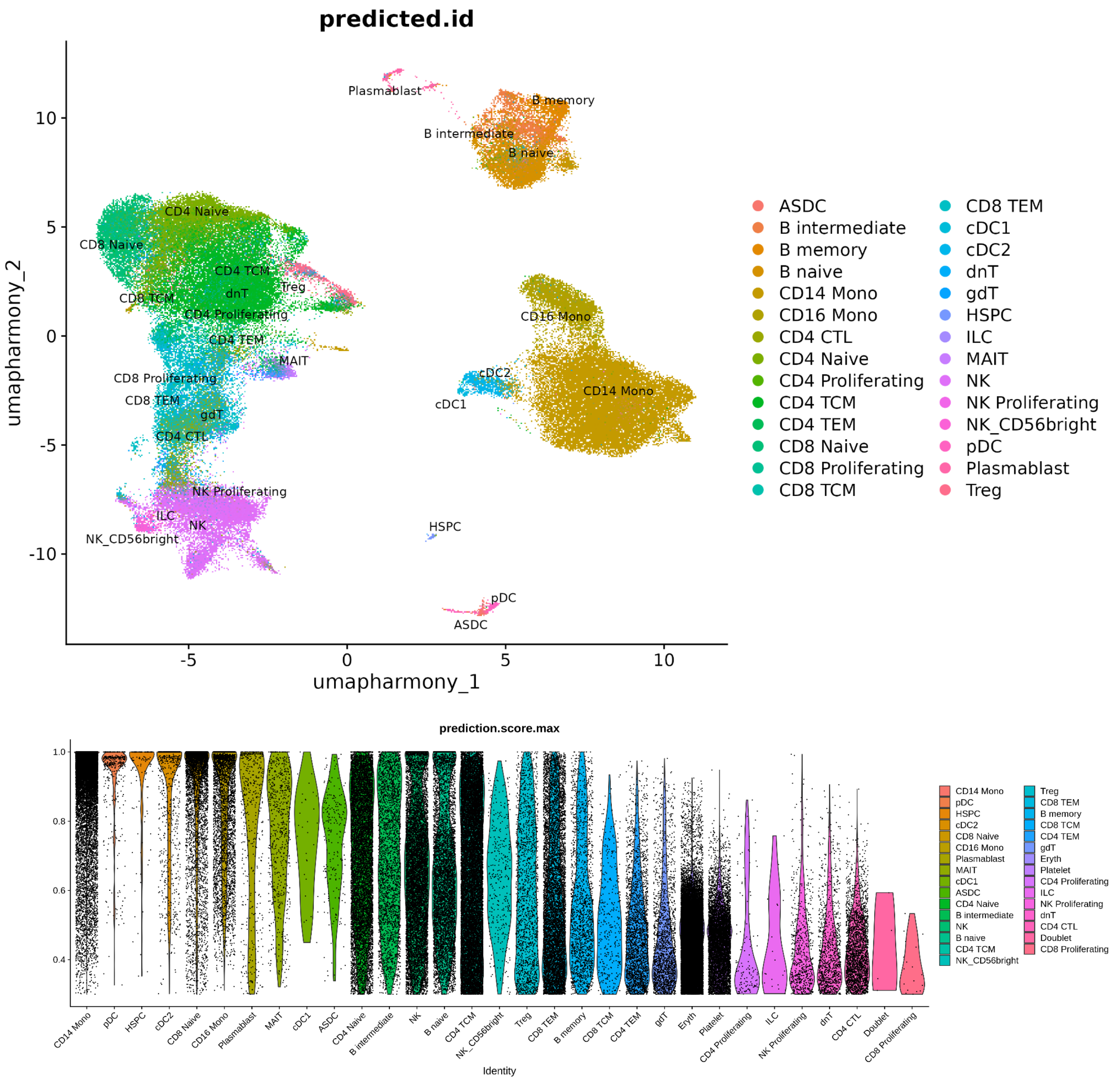


**Supplementary Figure 39. Cell type annotation of multiome bridging reference.** (**A**) 95,348 nuclei of 28 PBMC cell types for the combined multiome datasets. The x-axis and y-axis are the latent variables after removing batch effects using Harmony. (**B**) Prediction accuracy score across all cell types. The x-axis represents the 28 cell types annotated based on Azimuth reference. The y-axis indicates the prediction score. A cut-off at 0.3 was made to remove extremely low-accuracy nuclei.

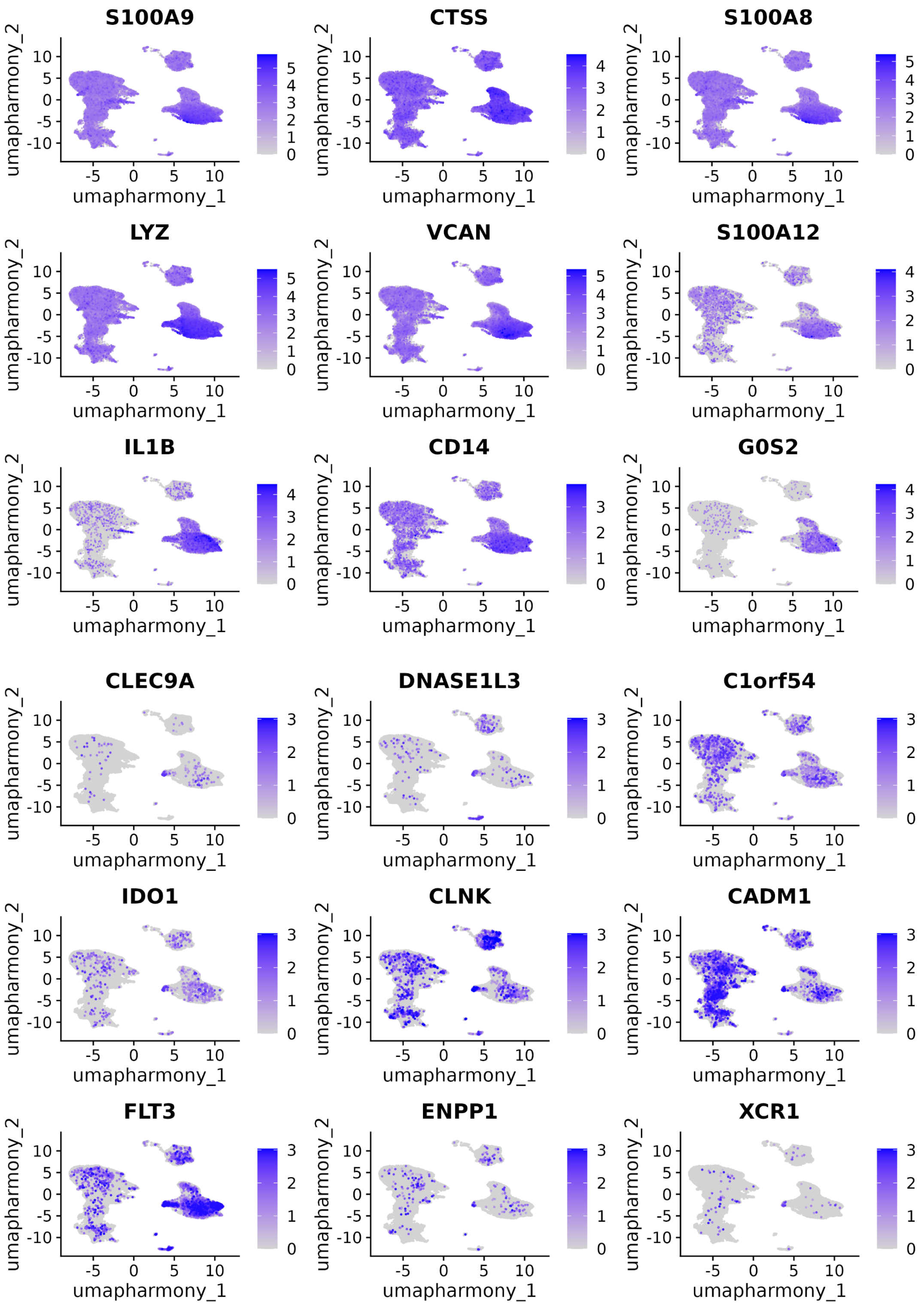


**Supplementary Figure 40. Canonical gene markers based on the gene activity scores for the multiome bridging dataset.** This plot shows the RNA levels of canonical markers for CD14+ monocytes (upper panel) and conventional type 1 dendritic cells (lower panel). The marker list are obtained from the Azimuth reference. The x-axis and y-axis are the latent variables after removing batch effects using Harmony. The color of each dot indicates the normalized expression values for the corresponding genes.

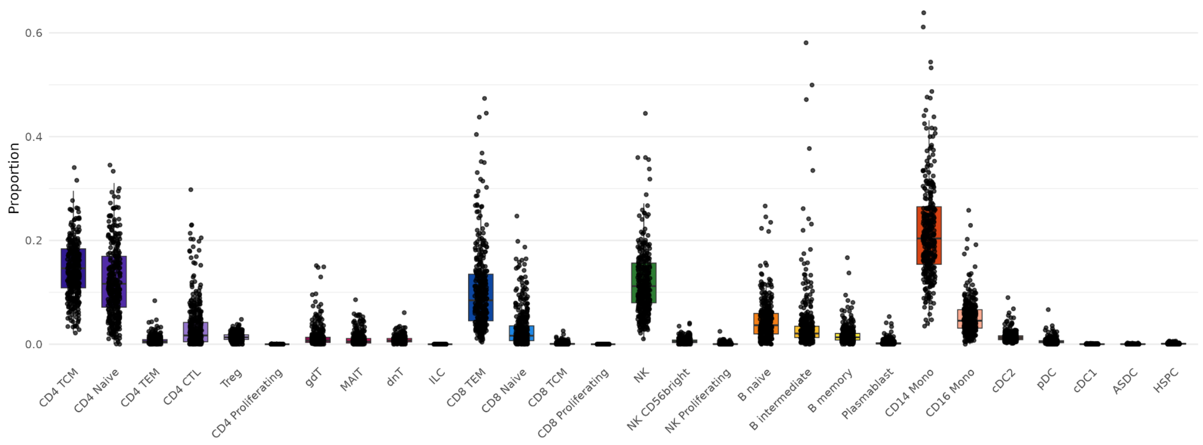


**Supplementary Figure 41. Distribution of the cell type composition at the donor level**. The x-axis indicates each of the 26 cell types. The y-axis indicates the proportion of each cell type within a donor.

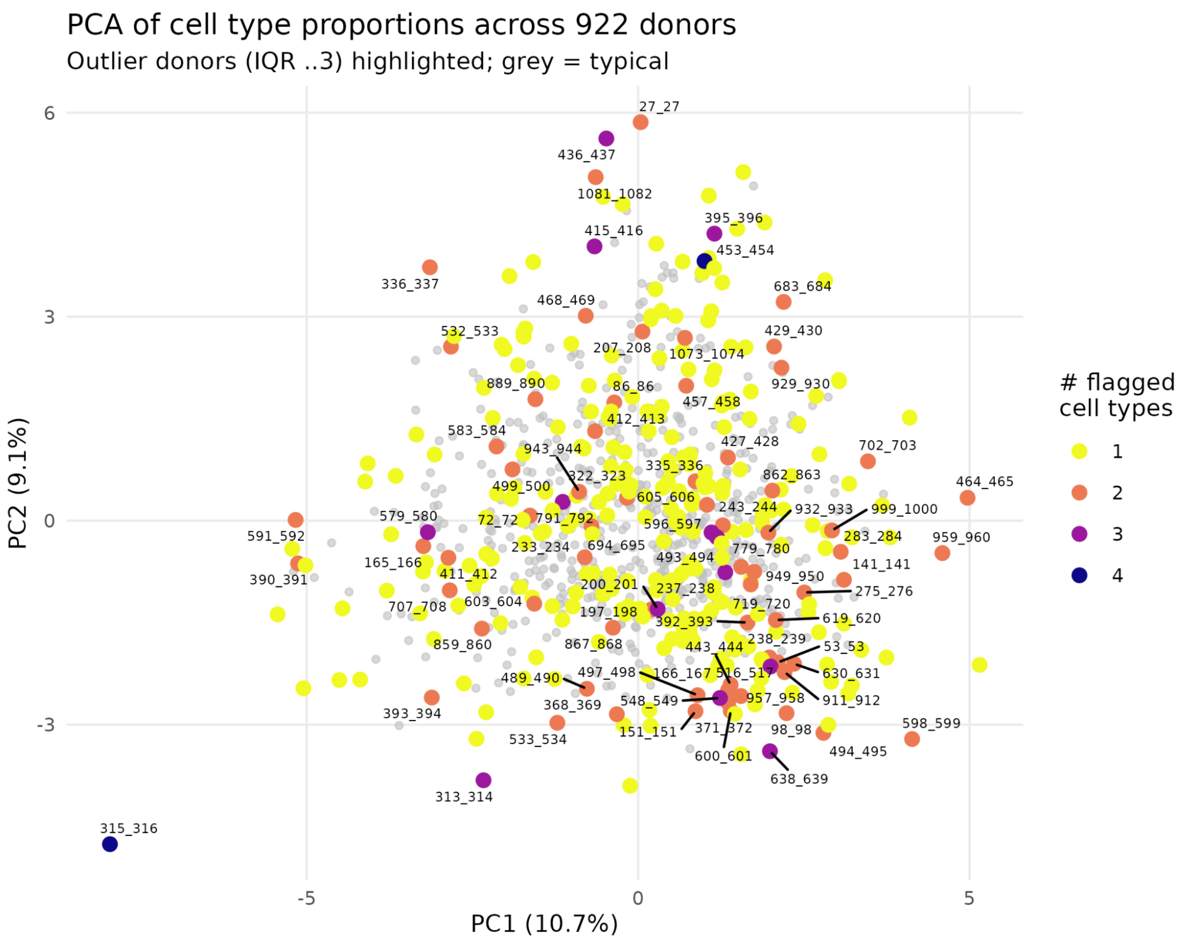


**Supplementary Figure 42. PCA plot of the cell type proportion across 922 donors**. Each dot represents one donor and the color indicates how many cell types are with proportion is outside the first and third quartiles of the interquartile range. A darker color indicates the donor is more likely to have abnormal cell type proportion.

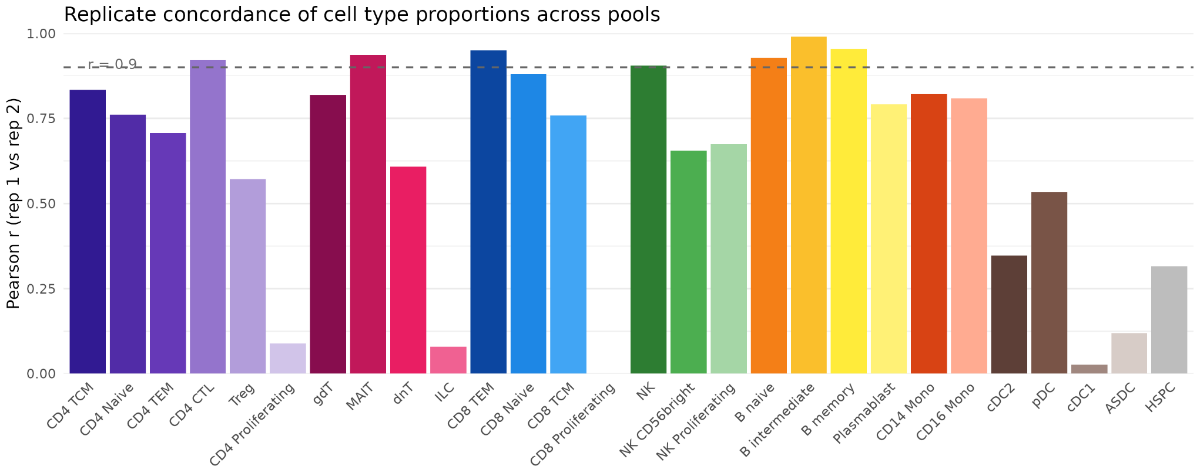


**Supplementary Figure 43. Concordance of cell type proportion between two repeated libraries for the same pool**. The x-axis indicates each of 26 cell types. The y-axis indicates the Pearson's correlation coefficient r between repeat 1 and repeat 2 across 139 pools. The horizontal dashed line indicates the coefficient of 0.9.

### CD4 Naive

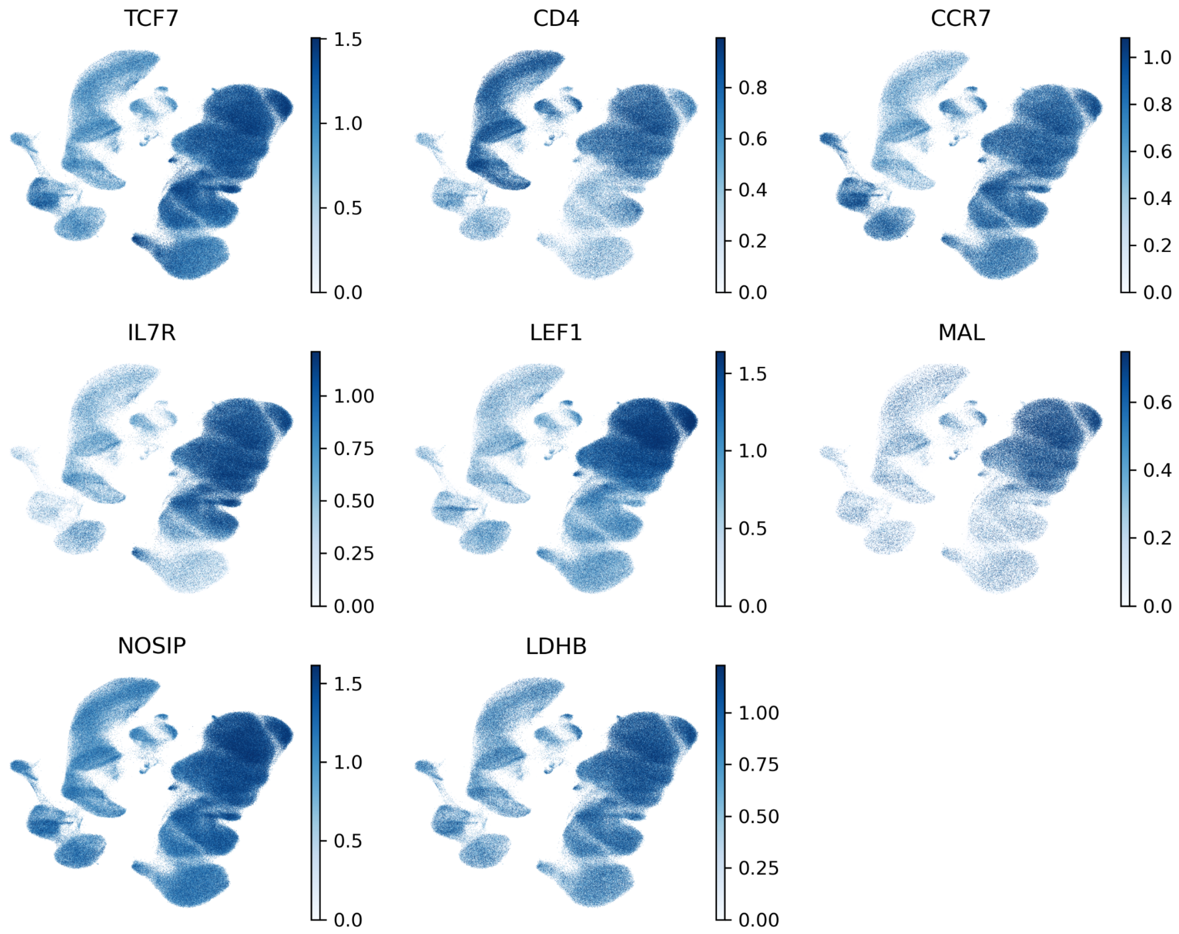


**Supplementary Figure 44. Canonical gene markers based on the gene activity scores for the main scATAC-seq dataset**. The UMAP coordinates are obtained from **Figure 1B**. The gene marker list is acquired from the Azimuth reference. The color scale indicates the log-normalized gene activity score for each marker gene. Due to the page limit, we have deposited the marker UMAP for the remaining 25 cell types on the project GitHub page, which is publicly accessible.

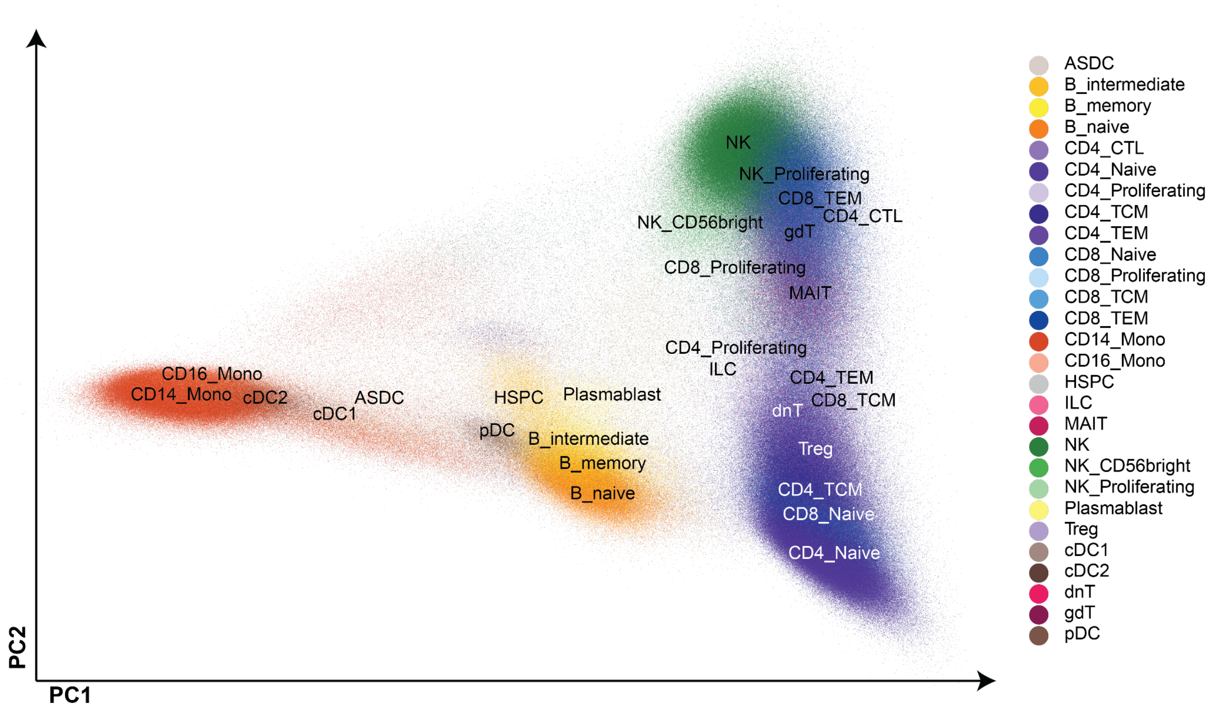


**Supplementary Figure 45. PCA plot of scATAC-seq data for 28 cell types**. All 3,546,117 nuclei were plotted and annotated with predicted cell types. Top 50,000 peaks were selected based on the feature variability score for dimensional reduction.

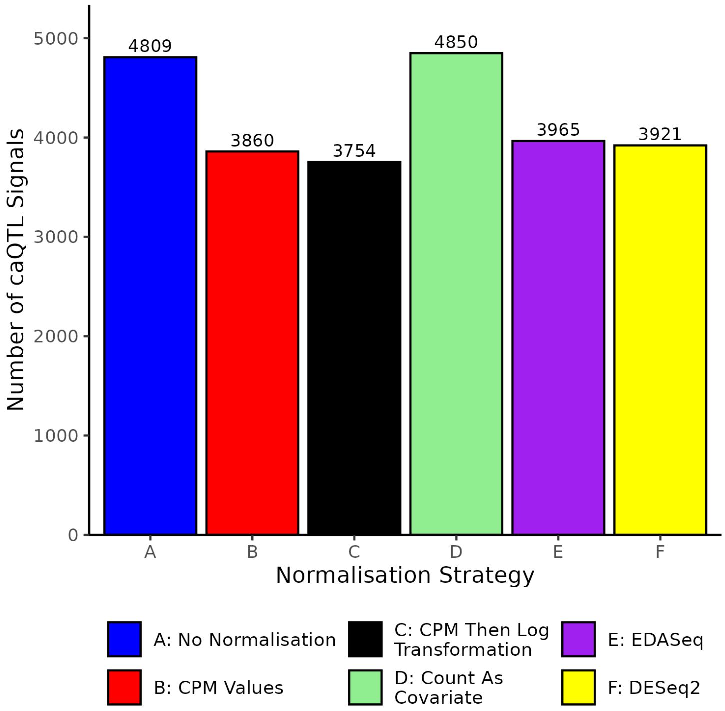


**Supplementary Figure 46. Comparison of different normalisation strategies for caQTL mapping.** Bars represent the number of significant caQTLs identified on Chromosome 1 in $CD14_{Mono}$ cells at FDR < 0.05.

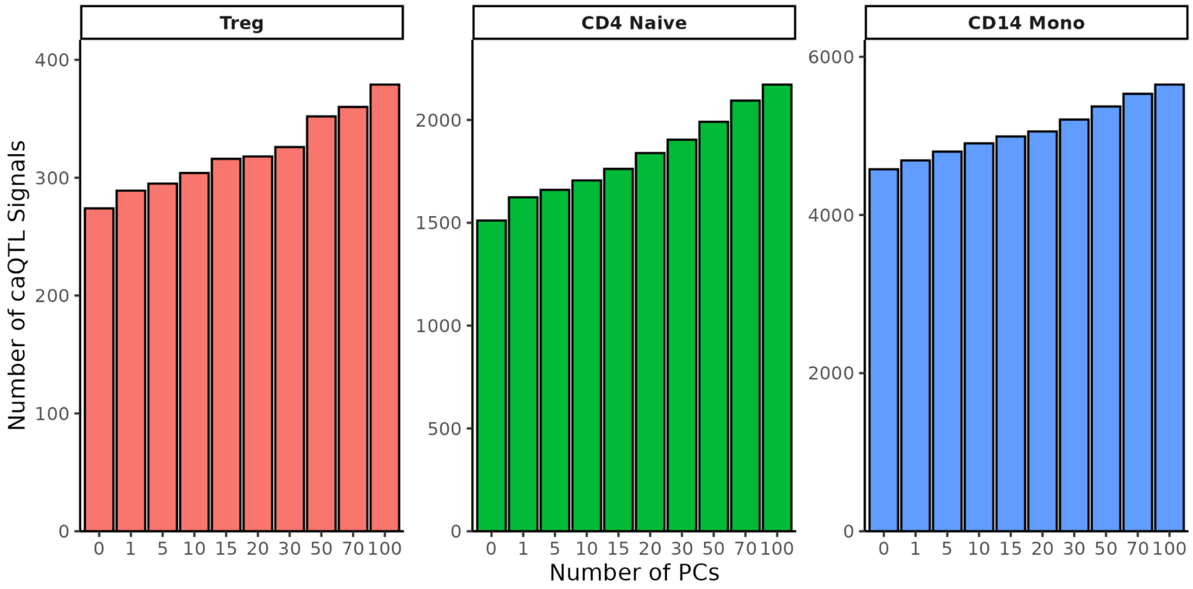


**Supplementary Figure 47. Effect of including different numbers of ATAC PCs as covariates on caQTL discovery.** Bars represent the number of significant caQTLs detected in $T_{reg}$, $CD4_{Naive}$, and $CD14_{Mono}$ cells on Chromosome 1 at FDR < 0.05.

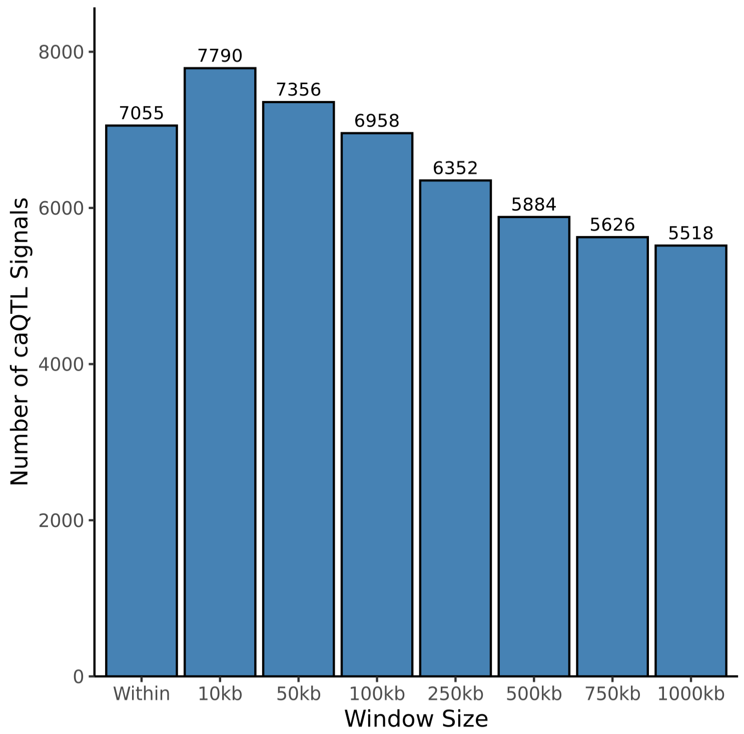


**Supplementary Figure 48. Effect of cis-window size on caQTL detection.** Bars show the number of significant caQTLs detected in $CD14_{Mono}$ cells on Chromosome 1 at FDR < 0.05, using window sizes ranging from 10 kb to 1000 kb (1 Mb). *'Within'* refers to restricting variant testing to those located entirely within the ATAC peak.

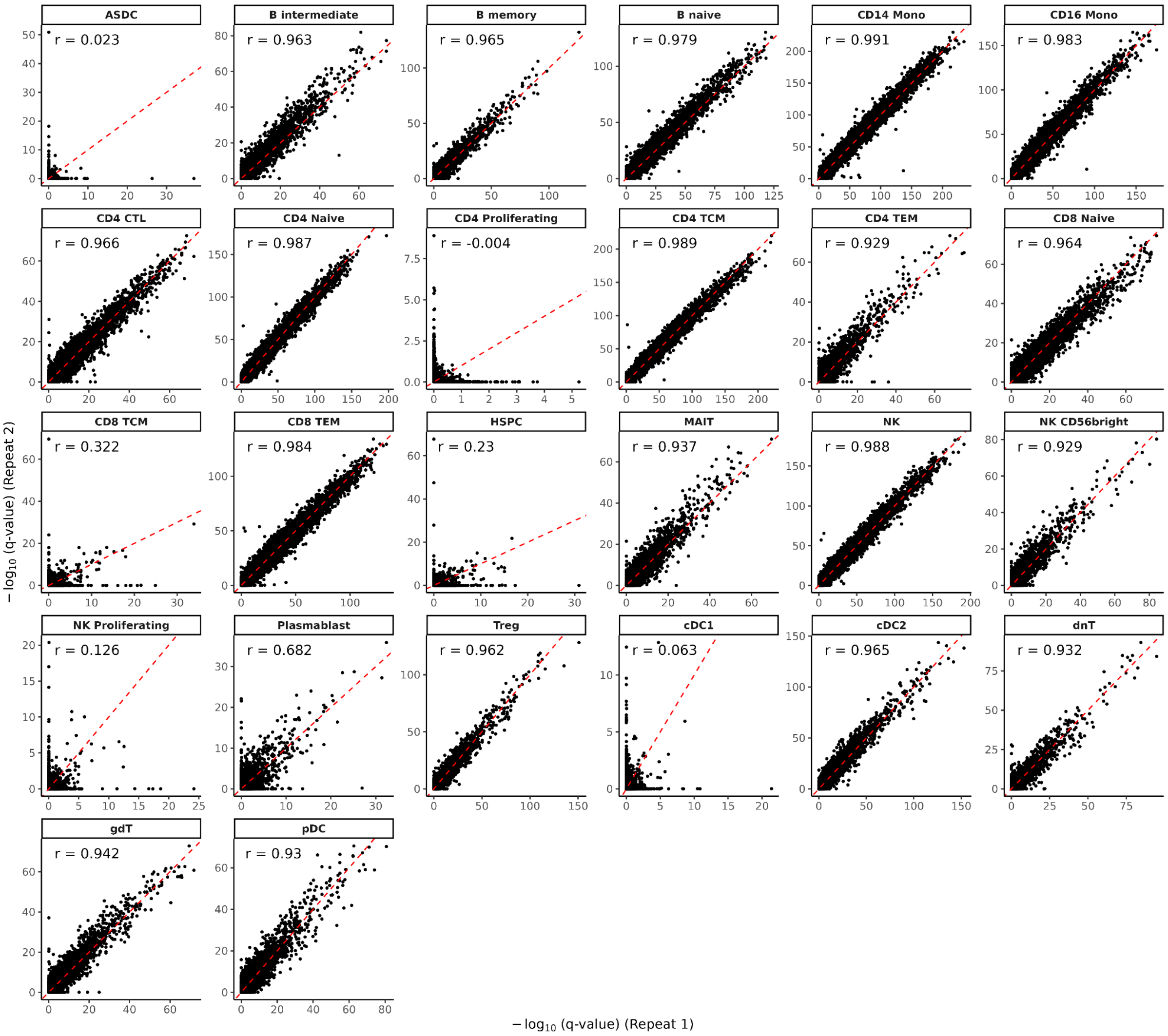
ASDC
r = 0.023
B intermediate
r = 0.963
B memory
r = 0.965
B naive
r = 0.979
CD14 Mono
r = 0.991
CD16 Mono
r = 0.983
CD4 CTL
r = 0.966
CD4 Naive
r = 0.987
CD4 Proliferating
r = -0.004
CD4 TCM
r = 0.989
CD4 TEM
r = 0.929
CD8 Naive
r = 0.964
CD8 TCM
r = 0.322
CD8 TEM
r = 0.984
HSPC
r = 0.23
MAIT
r = 0.937
NK
r = 0.988
NK CD56bright
r = 0.929
NK Proliferating
r = 0.126
Plasmablast
r = 0.682
Treg
r = 0.962
cDC1
r = 0.063
cDC2
r = 0.965
dnT
r = 0.932
gdT
r = 0.942
pDC
r = 0.93
$-\log_{10}$ (q-value) (Repeat 2)
$-\log_{10}$ (q-value) (Repeat 1)

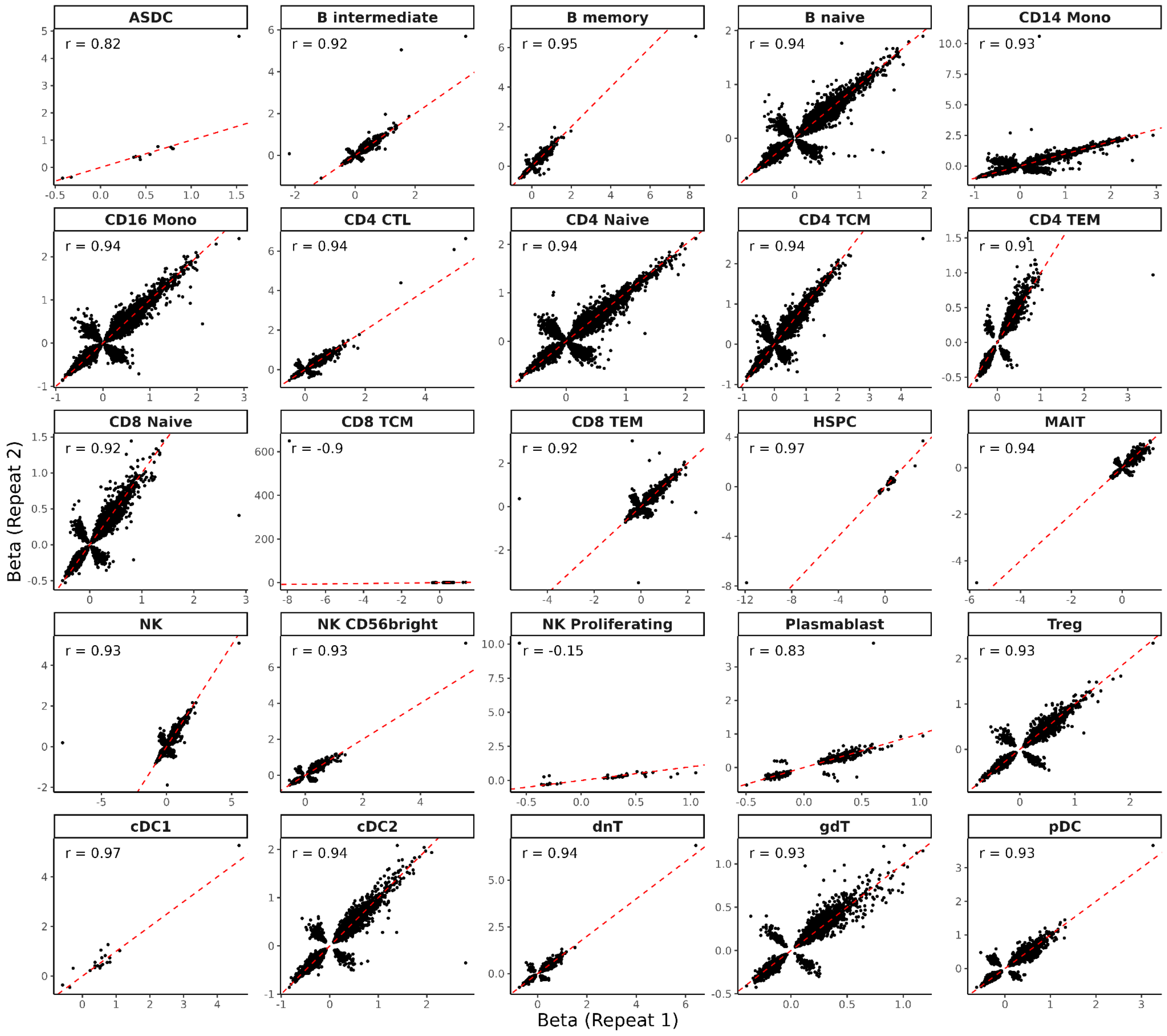


**Supplementary Figure 49. Reproducibility of caQTL mapping across repeat runs.** Comparison of -$\log_{10}$(q-value) and beta estimates between Repeat 1 and Repeat 2 across all 26 cell types. Each run used the number of ATAC PCs selected by the elbow method and a 1 Mb *cis*-window. Pearson's correlation between repeats was calculated for each cell type.
